# Phenome-wide associations of polygenic scores for anorexia nervosa and binge eating in the UK Biobank

**DOI:** 10.64898/2026.11.25.26368946

**Authors:** Helena L. Davies, Abigail R. ter Kuile, Stefana Aicoboaie, Jessica S. Johnson, Sang Hyuck Lee, Jonathan R. I. Coleman, Gerome Breen, Eating Disorders Working Group of the Psychiatric Genomics Consortium, Nadia Micali

**Affiliations:** Center of Eating and feeding Disorder Research, Mental Health Centre Ballerup, Copenhagen University Hospital – Herlev and Gentofte, Denmark; Institute of Biological Psychiatry, Mental Health Services, Copenhagen University Hospital, University of Copenhagen, Denmark; Social, Genetic, Developmental Psychiatry Centre, King’s College London, London, UK; Department of Clinical, Educational, and Health Psychology, University College London, London, United Kingdom; Department of Psychiatry, University of North Carolina at Chapel Hill School of Medicine, US; National Institute for Health and Care Research Biomedical Research Centre, King’s College London and South London and Maudsley National Health Service Trust, UK; Great Ormond Street Institute of Child Health, University College London, London, UK

## Abstract

Eating disorders are heritable and comorbid with many psychiatric and somatic conditions, potentially due to shared genetic risk. We investigated associations of genetic liability to anorexia nervosa and binge eating across diverse health-related phenotypes in the UK Biobank. Using PHESANT, we tested associations between SBayes-RC-derived polygenic scores for (1) anorexia nervosa (PGS_AN_) and (2) binge eating (PGS_BE_) with 8,657 phenotypes in UK Biobank (N=456,775). Analyses focused on European genetically-inferred ancestry, with secondary analyses examining the role of measured BMI and BMI polygenic scores (PGS_BMI_), and extending to African, Admixed American, South Asian, and East Asian genetically-inferred ancestry groups. Sensitivity analyses adjusted for sampling weights. PGS_AN_ and PGS_BE_ showed broadly similar associations with psychiatric and psychological vulnerability. However, they diverged across somatic, metabolic, and anthropometric traits, e.g., PGS_BE_ was associated with earlier pubertal timing across female (age at menarche; OR=0.93, 95% CI 0.93, 0.94, *p*<0.001) and male (e.g., relative age of voice breaking; OR=0.96, 95% CI 0.95, 0.97, *p*<0.001) markers of puberty, although associations with male markers were attenuated after BMI adjustment. No such associations were observed for PGS_AN_. Adjusting for BMI and PGS_BMI_ substantially attenuated anthropometric and metabolic associations, particularly for PGS_BE_. Analyses in non-European ancestry groups identified fewer significant associations, likely reflecting reduced power and poor cross-ancestry transferability of PGS. Adjusting for sampling weights altered effect sizes most for psychiatric and somatic traits, suggesting these associations are particularly susceptible to participation bias. Across the phenome, PGS_AN_ and PGS_BE_ exhibited common psychiatric liability alongside divergent somatic, anthropometric, and metabolic association patterns which reflects clinically observed differences in medical sequelae between the phenotypes.

## Introduction

Anorexia nervosa (AN) is an eating disorder characterised by restrictive eating, excessive exercise, and intense fear of weight gain, whilst binge eating (BE), i.e., eating an unusually large amount of food in a short period with a sense of loss of control, is a transdiagnostic symptom found in bulimia nervosa (BN), binge-eating disorder (BED), and a subtype of AN^1^. Twin-based heritability estimates range from 28% to 88% for AN, 54% to 83% for BN, and 41% to 57% for BED^2^, and liability-scale SNP-based heritability estimates stand at 5% for BE and 13% for AN^3^. GWAS have identified risk loci for AN and BE^3^, and for a model-derived binge-eating disorder phenotype^4^. Eating disorders are highly comorbid with psychiatric and somatic diseases^5^. Such comorbidity may arise through pleiotropy, whereby genetic variants affect multiple phenotypes independently (horizontal pleiotropy) or through one phenotype influencing another (vertical pleiotropy). Cross-trait associations can also arise through non-pleiotropic processes, such as indirect genetic effects^6^, assortative mating^7^, and selective participation^8^.

## Hypothesis-free approach

One way to explore cross-trait associations is via phenome-wide association studies (pheWAS), which tests associations between genetic liability and a wide range of phenotypes. This approach can uncover novel associations and generate hypotheses about broad health consequences of genetic liability to disordered eating. Previous pheWASs of AN genes^9^, AN polygenic score (PGS)^10,11^, and a PGS of a factor related to BN^11–13^ in datasets with a wealth of phenotypic information such as Mount Sinai BioMe^9^ and Vanderbilt University^10^ have uncovered relationships with autoimmune, gastrointestinal, metabolic, and psychiatric phenotypes. However, these studies are largely limited to individuals of European ancestry, restricting global generalisability. We partly address this by extending our analyses to other genetically-inferred ancestry groups, although the PGS remain derived from European-ancestry GWASs.

## The role of BMI and PGS_BMI_

Body mass index (BMI) can be indexed phenotypically (measured BMI) or genetically (e.g., via polygenic scores). Its role differs between AN and BE. In AN, low BMI is a core diagnostic feature, complicating separation of genetic effects on BMI from those on the disorder. Adjusting for BMI PGS attenuates associations between PGS_AN_ and metabolic traits, while psychiatric associations remain largely unchanged^10^. Stratified analyses by measured BMI further support heterogeneity in associations^9^. In contrast, BE is related to but distinct from body weight. Not all individuals with BE have overweight or obesity, and associations with metabolic outcomes such as hypertension and type 2 diabetes are independent of baseline BMI and change in BMI^14^. BE also shows specific psychiatric comorbidity not fully explained by obesity^15,16^ . However, binge eating is genetically correlated with obesity-related traits^3^. Genome-wide association study (GWAS)-by-subtraction analyses in which the genetic variance of BMI was removed from both BE and AN demonstrated substantially attenuated genetic correlations with anthropometric traits in both phenotypes and additionally with metabolic traits in AN^3^. Taken altogether, these findings underscore the importance of considering both measured BMI and the genetics of BMI when evaluating the relationships of PGS_AN_ and PGS_BE_ with diverse phenotypes. These approaches answer different questions: adjusting for measured BMI estimates associations independent of current adiposity, whereas adjusting for PGS_BMI_ estimates associations independent of genetic liability to higher BMI.

## Aims

The largest GWAS meta-analysis of binge-eating behaviour and AN to date^3^ provides a new opportunity to examine the phenotypic associations of genetic liability to these phenotypes. We conducted two pheWASs examining associations between health-related traits and PGSs of (1) AN (PGS_AN_) and (2) BE (PGS_BE_) in UK Biobank. Our study extends previous work in three ways. First, by examining PGS_AN_ and PGS_BE_ in the same sample and using the same methods, we can more directly compare where genetic liability to AN and BE shows shared and divergent associations, which separate studies are less able to. Second, whereas previous UK Biobank pheWASs^11,12^ used scores derived from earlier, smaller GWAS, we use the latest AN and BE GWASs, together with SBayesRC, which improves prediction. Third, we examine the role of measured and genetic BMI, the influence of selective participation, and associations in non-European genetic ancestry groups.

## Methods

### Participants

#### UK Biobank (UKB)

UKB is a population-based prospective study, established in 2006 to explore genetic and non-genetic factors associated with disease in middle and older age^17^. Recruitment of ∼500,000 UK-based adults aged 40 to 69 years across 22 assessment centres in the UK was completed in 2010. Primary analyses were restricted to all individuals of European genetically-inferred ancestry with available genotype data, defined as the largest homogeneous cluster from 4-means clustering of the first two genomic principal components calculated via *flashPCA*^18^. These individuals primarily report white ethnicities. To better isolate genetic liability rather than the downstream consequences of the eating disorders themselves, we additionally excluded 2,927 participants with anorexia nervosa or binge eating, identified via the mental health questionnaire^19^, hospital episode statistics, death register, and primary care linked data (total *n*= 456,775; 54% female; mean age=57 years).

## Statistical analysis

### Genotyping and quality control

Genotype data underwent standard quality control using PLINK v1.9. Variants with a minor allele frequency <0.01, missingness >0.02, or deviation from Hardy–Weinberg equilibrium (p<1×10⁻⁸) were excluded. Samples with genotype missingness >0.02, sex discrepancies between reported and genetic sex (reported females Fx < 0.5, reported males Fx > 0.9) were removed, as were samples recommended for exclusion by the central UK Biobank team due to missing data or being outliers on variant heterozygosity. Relatedness was assessed using KING, and one individual from each 1st, 2nd, or 3rd degree related pair (KING relatedness coefficient > 0.044) was randomly excluded using a greedy algorithm to retain an unrelated sample.

### Polygenic scores

We used SBayesRC^20^ and the imputed UK Biobank European LD reference panel to generate standardised PGS using GWAS summary statistics from Termorshuizen et al. (2026)^3^ for AN (PGS_AN_) and BE (PGS_BE_) with UK samples excluded.

### PheWAS

We used the PHEnome Scan ANalysis Tool (PHESANT), which is described elsewhere^21^ and in our Supplementary Materials. In brief, we included a total of 8,657 health-related traits (**Table S1**). We categorised our findings into the following six categories: somatic (including metabolic), anthropometric, psychiatric, psychological, lifestyle, and biomarkers. We controlled for age, sex, genotyping chip, assessment centre, and the first ten PCs (generated via *flashpca*^18^ within the genetically-inferred European ancestry sample). To account for multiple testing, we utilised the R tool *phenoSpD*^22^ which calculates phenotypic correlations and uses spectral decomposition of matrices to identify the number of independent traits and thus the Bonferroni-corrected *p-*value threshold of 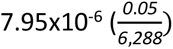

## Secondary analyses

### Role of BMI and PGS_BMI_

To assess the extent to which the associations observed in our pheWAS were independent of BMI, we repeated each pheWAS twice, in separate models: first adjusting for measured BMI, and second adjusting for BMI PGS (PGS_BMI_). These analyses address different questions. Measured BMI captures current adiposity, reflecting genetic and environmental influences as well as potential consequences of the phenotypes themselves (e.g., weight change associated with eating disorders). Measured BMI may therefore act as a confounder or as a mediator of PGS-phenotype associations, and adjustment estimates associations independent of current BMI rather than removing confounding. PGS_BMI_ captures only genetic liability to higher BMI, and was used to assess whether associations reflect genetic variants shared between eating disorders and BMI. PGS_BMI_ was obtained from the UK Biobank PRS Release (Standard PRS set, field [X], Version [2])^23^, which was derived by Genomics plc using external GWAS training data only, with no UK Biobank participants, therefore avoiding overlap with our target sample. Scores were standardised to mean 0 and SD 1 .

### Beyond European genetically-inferred ancestry

We created PGS_AN_ and PGS_BE_ in participants of African (*n*=3,125), South Asian (*n=*4,404), East Asian (*n=*864), and Admixed American (*n=*773) genetically-inferred ancestry, identified via principal components analysis across the whole sample. Discovery GWAS of eating disorders in genetically-inferred ancestries other than European are lacking. We therefore used the European genetically-inferred ancestry summary statistics from Termorshuizen et al. (2026)^3^ to calculate the scores with SBayesRC. SBayesRC uses the LD reference to model the correlation between SNPs in the discovery GWAS when estimating joint SNP effects, so the LD reference should match the ancestry of the discovery sample rather than the target sample^20^. We therefore used the imputed UK Biobank European LD reference panel for all ancestry groups. Within each genetically-inferred ancestry, we identified ten principal components using *flashpca*^18^ for inclusion as covariates in ancestry-specific pheWAS models, alongside sex, age, genotyping chip, and assessment centre. We note that applying European summary statistics and reference panel to samples of other genetically-inferred ancestries is likely to reduce predictive accuracy^24^, although SbayesRC’s incorporation of functional annotations has been shown to improve cross-ancestry prediction from European ancestry GWAS by 16-34% on average relative to earlier methods^20^.

## Sensitivity analyses

### Role of selective participation

UKB is not representative of the UK population; participants are generally healthier, wealthier, and more highly educated^8^. Therefore, to assess the impact of selective participation, we applied sampling weights within each pheWAS that increased the representativeness of UKB to that of the UK population^8^ via *svyglm* from the R package *survey*^25^.

## Ethical approval

The authors assert that all procedures contributing to this work comply with the ethical standards of the relevant national and institutional committees on human experimentation and with the Helsinki Declaration of 1975, as revised in 2013. UK Biobank is registered as a Research Tissue Bank (North West – Haydock Research Ethics Committee, 21/NW/0157). All data and sample applications may use this ethical clearance to conduct their research. Separate Research Ethics Committee (“REC”) or other ethical clearance is not required. This research was conducted under application 82087 (PI: Jonathan Coleman).

## Results

### Main analyses

We found significant results across six categories: anthropometric, somatic, psychological, psychiatric, lifestyle, and biomarkers (**Figure 1; Table S2-S4**), which we describe in further detail below.

**Figure 1.**
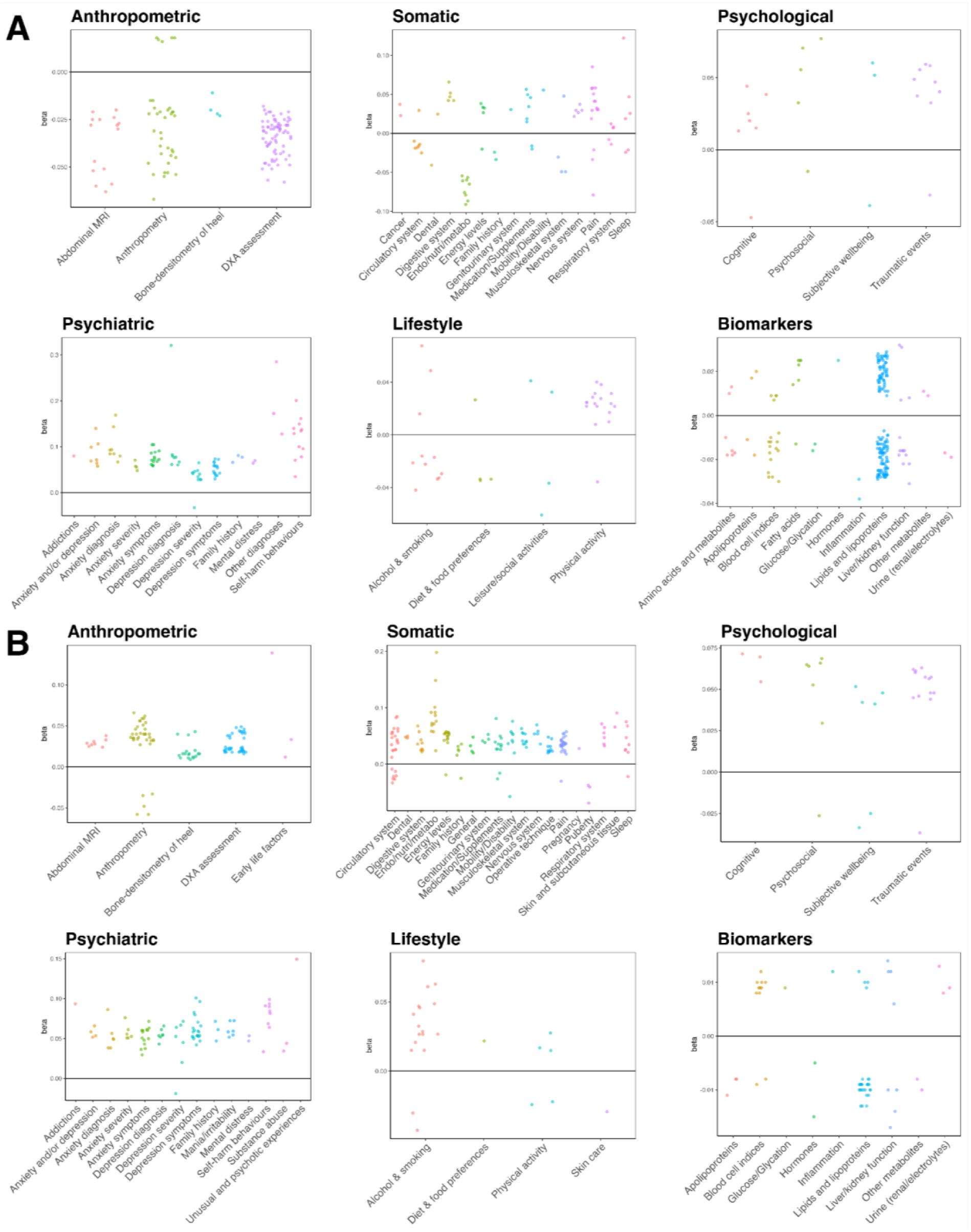
Significant phenome-wide association results between A) anorexia nervosa and B) binge eating polygenic scores with health-related traits in UK Biobank, split into six categories: anthropometric, somatic, psychological, psychiatric, lifestyle, and biomarkers. Analyses are adjusted for age, sex, genotyping chip, assessment centre, and the first ten principal components. *Note.* endo/nutri/metabo=endocrine, nutritional, metabolic diseases.

### Somatic

PGS_BE_ demonstrated a number of significant positive associations with traits and diseases linked to the circulatory system, energy levels (e.g., feeling tired, waking unrefreshed), genitourinary system, mobility/disability, skin and subcutaneous tissue. For example, each standard deviation (SD) increase in PGS_BE_ was linked with higher odds of having an ICD10 diagnosis of hypertension (OR=1.03, 95% CI 1.02, 1.03, *p<*0.001; **Figure S1**). PGS_BE_ was also associated with earlier pubertal timing, e.g., each SD increase in PGS_BE_ was associated with lower odds of being in higher categories for the following: age at menarche (OR=0.93, 95% CI 0.93, 0.94), relative age at voice breaking (OR=0.96, 95% CI 0.95, 0.97), and relative age at first facial hair (OR=0.96, 95% CI 0.95, 0.97, *p*s<0.001). Whilst PGS_AN_ displayed weaker or no associations with traits in these categories, both PGS_AN_ and PGS_BE_ were positively associated with digestive system traits. For example, PGS_AN_ was associated with higher odds of receiving an irritable bowel syndrome diagnosis (OR=1.05, 95% CI 1.03, 1.07, *p*=2.1x10^-9^) and PGS_BE_ was associated with higher odds of recent watery stools (OR=1.03, 95% CI 1.02, 1.04, *p*=1.03x10^-7^). For nervous system traits, both PGSs were linked with higher impact of, for example, recent dizziness (PGS_AN_ OR = 1.03, 95% CI 1.02, 1.04; PGS_BE_ OR = 1.04, 95% CI 1.03, 1.05; *p*s<0.001; **Figure S1**). We identified diverging patterns of associations for some categories, including endocrine, nutritional, metabolic, and musculoskeletal system-related traits. For these, PGS_AN_ was largely negatively associated and PGS_BE_ largely positively, e.g., an ICD10 diagnosis of gonarthrosis (PGS_AN_ OR = 0.97, 95% CI 0.96, 0.98; PGS_BE_ OR = 1.03, 95% CI 1.02, 1.04; *p*s<0.001).

### Anthropometric

PGS_AN_ and PGS_BE_ demonstrated diverging associations across all anthropometric categories: abdominal magnetic resonance imaging, anthropometry, bone-densitometry of heel, dual X-ray absorptiometry assessment, and early life factors (e.g., birth weight; **Figure S2**). Bone density and mineral content, fat mass and percentage, fat and muscle volume, and fat-free mass traits were positively associated with PGS_BE_, but negatively with PGS_AN_. For example, a one SD increase in PGS_AN_ and PGS_BE_ were linked with a 0.05 SD decrease and 0.06 SD increase in hip circumference, respectively (*p*s<0.001). Further, PGS_AN_ was associated with 0.03 SD lower basal metabolic rate whilst PGS_BE_ was associated with a 0.04 SD higher rate (*p*s< 0.001).

### Psychiatric

PGS_AN_ and PGS_BE_ showed largely similar associations across all psychiatric categories including addictions, anxiety, depression, mental distress, and self-harm behaviours (**Figure S3**). For example, PGS_AN_ and PGS_BE_ were both positively associated with miserableness (PGS_AN_ OR = 1.04, 95% CI 1.04, 1.05; PGS_BE_ OR = 1.05, 95% CI 1.05, 1.06; *p*s<0.001) and with lifetime contemplation of self-harm (PGS_AN_ OR = 1.10, 95% CI 1.09, 1.12; PGS_BE_ OR = 1.07, 95% CI 1.06, 1.09; *p*s<0.001). Points of divergence between PGS_BE_ and PGS_AN_ included that PGS_BE_ was significantly positively associated with traits linked with mania/irritability (e.g., racing thoughts; OR=1.06, 95% CI 1.04, 1.08, *p*=4x10^-10^), substance abuse (e.g., psychoactive substance abuse; OR=1.04, 95% CI 1.03, 1.04, *p*<0.001), and unusual and psychotic experiences (i.e., ever believed in an un-real conspiracy against self; OR=1.16, 95% CI 1.09, 1.23, *p*=9.6x10^-7^) whilst PGS_AN_ was significantly positively linked with bipolar disorder (OR=1.14, 95% CI 1.08, 1.19, *p*=5.7x10^-7^), obsessive-compulsive disorder (OCD; OR=1.33, 95% CI 1.19, 1.49, *p*=7.2x10^-7^), and schizophrenia (OR=1.19, 95% CI 1.11, 1.27, *p*=1.5x10^-6^).

### Psychological

Both PGS_AN_ and PGS_BE_ were positively associated with self-reported traumatic events, e.g., feeling hated by a family member as a child (PGS_AN_ OR = 1.06, 95% CI 1.05, 1.08; PGS_BE_ OR = 1.06, 95% CI 1.04, 1.07; *p*s<0.001) and negatively with believing one’s own life is meaningful (PGS_AN_ OR = 0.96, 95% CI 0.95, 0.97; PGS_BE_ OR = 0.97, 95% CI 0.96, 0.98; *p*s<0.001; **Figure S4**). PGS_AN_ was positively associated with cognitive traits such as time spent on matrix reasoning puzzles which measures non-verbal fluid reasoning (β = 0.03, 95% CI 0.014, 0.036, *p*<0.001), and with the number of incorrect matches on a visual memory task pair (β = 0.02, 95% CI 0.01, 0.021, *p*<0.001).

### Lifestyle

PGS_AN_ showed positive associations with physical activity traits, e.g., one SD increase in PGS_AN_ was linked with a 0.017 SD increase in summed days of walking, moderate, and vigorous activity per week (95% CI 0.014, 0.021, *p*<0.001; **Figure S5**), equivalent to approximately 0.08 additional days per week. In line with this, PGS_AN_ was associated with lower odds of being in a higher category of time spent watching TV (OR=0.94, 95% CI 0.94, 0.95, *p*<0.001). PGS_BE_ demonstrated a number of positive associations with alcohol and smoking traits including cannabis use (OR = 1.05, 95% CI 1.04, 1.06, *p*<0.001) and the frequency of the inability to cease drinking (OR = 1.06, 95% CI 1.04, 1.09, *p*<0.001; **Figure S5**).

### Biomarkers

PGS_AN_ showed a number of significant associations with biomarkers where PGS_BE_ did not, particularly with fatty acids and amino acids and metabolites (**Figure S6**). For instance, PGS_AN_ was negatively associated with total fatty acids (β=-0.01, 95% CI -0.02, -0.01, *p*<0.001) whereas PGS_BE_ showed no significant association. We found different associations with hormones. PGS_AN_ was positively associated with sex hormone-binding globulin (β=0.02, 95% CI 0.02, 0.03, *p*<0.001), whilst PGS_BE_ was negatively associated with testosterone (β=-0.005, 95% CI -0.006, -0.003, *p*<0.001) and Insulin-like Growth Factor 1 (β=-0.01, 95% CI -0.02, -0.01, *p*<0.001). Similarly, PGS_AN_ demonstrated negative associations with inflammation biomarkers such as C-reactive protein (β=-0.04, 95% CI -0.04, -0.03, *p*<0.001) and urine biomarkers reflecting renal/electrolyte function such as sodium (β=-0.02, 95% CI -0.02, -0.01, *p*=3x10^-10^), whilst PGS_BE_ showed positive associations (β=0.01, 95% CI 0.01, 0.02; β=0.01, 95% CI 0.006, 0.011, *p*s <0.001). PGS_BE_ was broadly negatively (and non-significantly) associated with lipids and lipoproteins, whilst PGS_AN_ showed multiple significant associations in both directions. Both PGS_AN_ and PGS_BE_ exhibited a pattern of significant associations, both positive and negative, with blood cell indices and liver/kidney function biomarkers.

### Secondary analyses

#### Role of BMI and PGS_BMI_

**BMI.** Controlling for BMI had little effect on the associations of PGS_BE_ or PGS_AN_ with lifestyle, psychiatric, or psychological traits (**Figure 2; Tables S5 & S6; Figures S7-S12**). However, most associations with anthropometric traits, and all endocrine, nutritional, metabolic, and musculoskeletal system traits were substantially attenuated (**Figures S14-S16**). After BMI adjustment, the association with PGS_BE_ and age at menarche was attenuated but remained significant (OR = 0.95, 95% CI 0.94, 0.96, *p*<0.001), whereas associations with relative age of voice breaking (OR = 0.98, 95% CI 0.97, 1.00, *p*=0.009) and first facial hair (OR = 0.98, 95% CI 0.97, 0.99, *p*=2.4 × 10⁻⁴) were attenuated by approximately half and no longer met the Bonferroni-corrected threshold (Tables S5-S8). PGS_AN_ associations with biomarkers were also attenuated after BMI adjustment (**Figures S17**). By comparison, PGS_BE_ demonstrated a *greater* number of significant associations with biomarkers after BMI adjustment. This pattern may reflect a suppression effect, in which BMI -being positively correlated with the PGS_BE_ and also associated with biomarkers -obscures the direct PGS-biomarker association until included in the model (**Figures S18)**, but could also reflect mediation, over-adjustment, or collider bias, which these data cannot distinguish. Among the top 10% of absolute differences in effect size after controlling for BMI, an overwhelming majority were observed in the anthropometric trait categories for both PGS_AN_ and PGS_BE_ (**Figure 2)**. However, the top 10% of relative effect size differences included anthropometric (23%), somatic (19%), and psychological (10%) traits for PGS_AN_ and biomarkers (29%) and lifestyle (5%) for PGS_BE_. Overall, PGS_AN_ appeared less impacted by BMI adjustment than PGS_BE._ Adjustment for PGS_BMI_ produced a similar but weaker pattern to adjustment for measured BMI, with smaller attenuation of anthropometric and metabolic associations (**Figures S19-S31; Tables S7 & S8; Supplementary Results**).

**Figure 2.**
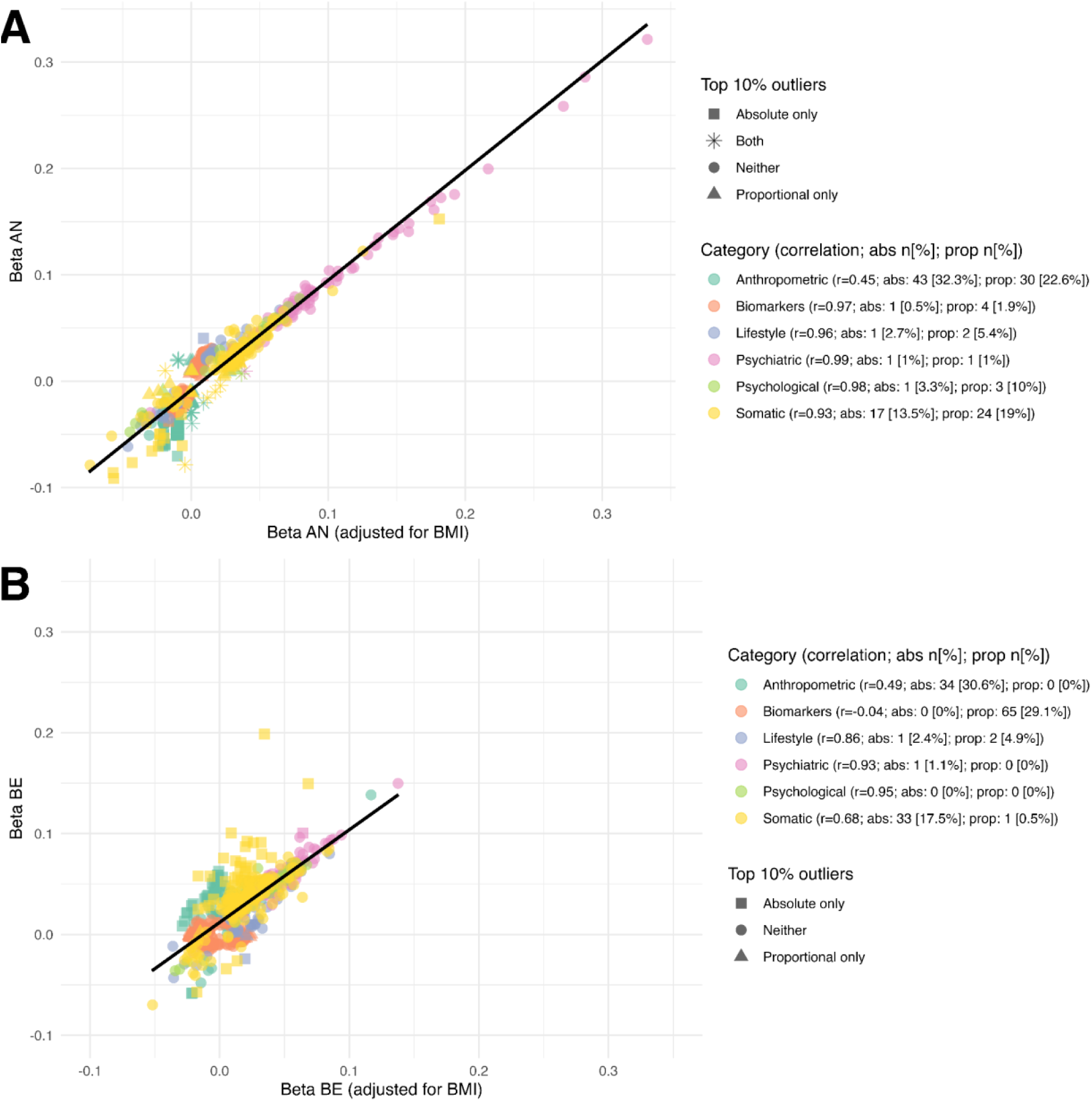
Correlation between original analyses results and analyses adjusted for body mass index in a phenome-wide association study in which the exposure is A) anorexia nervosa polygenic score and B) binge eating polygenic score, and the outcomes are UK Biobank health-related traits. Number and percentage of traits within each category that are in the absolute (abs) and proportional (prop) top 10% of change following adjustment are also displayed. *Note*. Odds ratios are converted to logarithmic odds for consistency with beta. The low correlation between BMI-adjusted and unadjusted betas for biomarkers in B but zero values falling in the top 10% of absolute change is due to very small effect sizes in this category (many close to zero).

#### Beyond European genetically-inferred ancestry

We discovered no Bonferroni significant associations in those of Admixed American or African genetically-inferred ancestry (**Tables S9 & S10**). In those of East Asian genetically-inferred ancestry, a one SD increase in PGS_AN_ was associated with a 0.13 SD lower platelet count (β = -0.13, 95% CI -0.19, -0.06, *p*<0.001; **Table S11**). Among participants of South Asian genetically-inferred ancestry, several anthropometric traits showed Bonferroni-significant associations with PGS_BE_, including waist circumference (β = 0.07, 95% CI 0.04, 0.1, *p*<0.001) and weight (β = 0.07, 95% CI 0.04, 0.1, *p*<0.001; **Table S12)**. Further, a one SD increase in PGS_BE_ was associated with a 0.05 SD higher basal metabolic rate (β = 0.05, 95% CI 0.03, 0.07, *p*<0.001). Sampling weights are currently only available for those of European genetic ancestry.

### Sensitivity analyses

#### Role of selective participation

Adjustment for sampling weights most strongly affected psychiatric (**Figures S32 & S33**) and somatic (**Figures S34 & S35)** traits for PGS_AN_ and PGS_BE_. Of these, ∼40% of psychiatric and somatic traits were in the top 10% of absolute effect size differences following sampling weight adjustment in the PGS_AN_ pheWAS, whilst 19% of somatic and 15% of psychiatric traits were in the top 10% for the PGS_BE_ pheWAS (**Figures 3**). Anthropometric traits and biomarkers had the lowest percentage of traits (0-4%) in the top 10% affected for both the PGS_BE_ pheWAS and 16% in the PGS_AN_ pheWAS (**Figures S36-S39**). Lifestyle traits showed some attenuation in both the PGS_AN_ (**Figures S40**) and PGS_BE_ pheWAS (**Figures S41**). We observed a larger effect from the adjustment in the psychological traits in the PGS_BE_ pheWAS (*r*=0.97; 13% in top 10%; **Figure S42**) than the PGS_AN_ pheWAS (*r*=1; 0% in top 10%; **Figure S43**).

**Figure 3.**
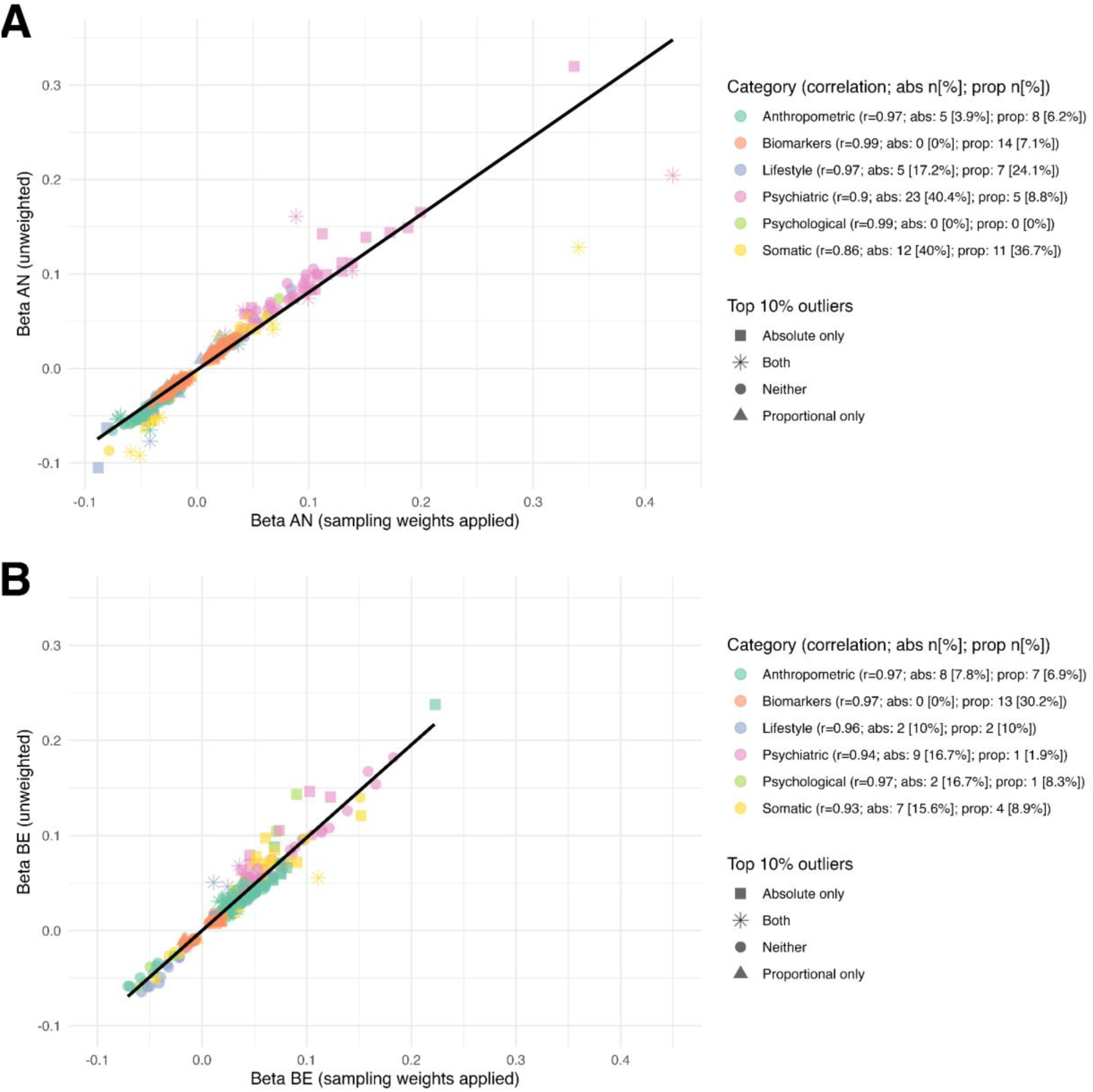
Correlation between original analyses results and analyses adjusted for sampling weights in a phenome-wide association study in which the exposure is A) anorexia nervosa polygenic score and B) binge eating polygenic score, and the outcomes are UK Biobank health-related traits. Number and percentage of traits within each category that are in the absolute (abs) and proportional (prop) 10% of change following adjustment are also displayed. *Note*. Odds ratios are converted to logarithmic odds for consistency with beta.

## Discussion

### Divergent somatic and metabolic associations

By systematically evaluating over 8,500 traits, we found that genetic liability to AN and BE showed divergent, often opposing, somatic and metabolic associations, particularly across musculoskeletal, endocrine, nutritional, and metabolic diseases, as well as most anthropometric traits. PGS_BE_ was linked to higher body mass and metabolic load, with similar anthropometric associations observed in individuals of South Asian genetically-inferred ancestry. In contrast, PGS_AN_ was associated with lower body mass, reduced bone density, and markers of energy deficiency. One explanation is a shared genetic axis of energy balance and appetite regulation, with genetic liability to AN and BE lying in opposite directions, rather than separate biological pathways; this would be consistent with negative genetic correlations between AN and BMI and metabolic traits^3^, and with the attenuation of these associations after BMI adjustment. However, AN and BE are positively genetically correlated (rg = 0.46), and most of their genetic variance is not shared with BMI^3^, so they are not simply opposite ends of one dimension. The two scores derive from GWAS of different phenotypes with different statistical power, thus these differential association patterns cannot by themselves establish distinct mechanisms. Anthropometric associations are typically less attenuated in within-family designs than behavioural and cognitive traits^26^, suggesting these associations are less likely to reflect indirect genetic effects. Nonetheless, these divergent associations may help explain why metabolic comorbidities differ across eating disorders^27^.

Our findings suggest earlier pubertal timing is linked specifically to PGS_BE_ and not PGS_AN_. PGS_BE_ associations converged with previously-described links with earlier age at menarche in females in UK Biobank^28^ and extended to associations with male pubertal markers, including age of first facial hair and voice breaking. Our findings are consistent with phenotypic patterns showing younger pubertal onset in bulimia nervosa (BN) and eating disorder not otherwise specified (EDNOS)^29^, while evidence for AN has been more mixed^30,31^. Genetic studies further support a specific pubertal timing pattern related to BED; age-at-menarche PGS is significantly negatively associated with BED, but not with AN or, interestingly, with BN (however this group was the smallest and thus most limited in power, and did not exclude people with a history of AN)^28^. Genetic correlation analyses mirror these results, with age at menarche negatively correlated with BE^3^, but no significant correlations with AN^32^. One possible mechanism is estradiol, which rises during puberty and may have an effect on genetic risk for BE. Girls with comparatively lower estradiol levels during this developmental period have shown enhanced genetic influence and greater binge-eating liability^33^. After adjustment for BMI, the association with age at menarche was attenuated but remained significant, whereas associations with male pubertal markers were attenuated by approximately half and no longer met the Bonferroni-corrected threshold. This may indicate that the male associations are partly explained by BMI, although adult BMI may itself lie partly downstream of pubertal timing, so this attenuation should be interpreted cautiously. Together, these findings suggest that genetic liability to BE may be associated with earlier pubertal timing, particularly in females, which warrants further investigation.

### Shared and distinct psychiatric liability

In psychiatric and psychological domains, both PGSs were positively associated with depression, anxiety, distress, self-harm, and self-reported childhood trauma, pointing to substantial shared psychiatric vulnerability. However, assortative mating across psychiatric conditions can inflate apparent genetic overlap between them^7^ so the extent of shared liability may be overestimated. Importantly, these findings do not imply that individuals at high genetic risk of AN or BE are responsible for or genetically determined to experience trauma. Self-reports of traumatic experiences reflect a complex interplay of factors, including negative affectivity^34^, risk-taking^34^, subjective interpretation processes^34^, memory recall^34^, and current mood^35,36^. Thus, observed associations may reflect genetically influenced tendencies shaping the perception and reporting of trauma, or passive gene-environment correlation^37^ (e.g., parents may carry genetic liability to traits that are associated with creating higher-risk environments, and children inherit both genetic susceptibility and increased exposure to trauma), rather than a direct causal link. GWAS may also capture variants associated with the effects of risk factors such as trauma^37^.

While both PGSs showed shared vulnerability with common psychiatric traits, they also indexed partly distinct associations. PGS_BE_ was positively associated with mania/irritability, substance use, psychotic-like experiences, self-care problems, and risk-taking -consistent with a long-standing recognition of the role of impulsivity in BE^38^. In contrast, PGS_AN_ showed positive associations with bipolar disorder, OCD, schizophrenia, and poorer performance on cognitive measures. This latter finding may indicate poorer visual memory, attention, or executive function, but also sits somewhat at odds with the genetic overlap between AN and higher educational attainment^32^. Instead, it may reflect maladaptive cognitive consequences of over-control found in AN^39^. Longer time spent on matrix-reasoning puzzles does not necessarily indicate poorer performance, and may reflect perfectionistic cognitive styles characteristic of AN. Given the cross-sectional design, these associations should be interpreted cautiously. Cognitive traits are also among those most attenuated in within-family analyses^26^, so indirect genetic effects and population stratification may also contribute to these associations.

## Distinct lifestyle traits

Lifestyle associations also diverged between PGS_AN_ and PGS_BE_. PGS_AN_ was positively associated with physical activity and lower sedentary behaviour, in line with clinical evidence of hyperactivity and overexercise in AN^40^ and an established positive genetic correlation between physical activity and AN^32^. PGS_AN_ showed some negative significant associations with, for example, frequency of consuming >6 alcohol units. Conversely, PGS_BE_ was positively associated with tobacco, cannabis, and alcohol use, consistent with reported overlap between binge eating and addictive behaviours^41^ and possibly reflecting shared genetic liability to reward sensitivity or impulsivity, although substance use associations are particularly susceptible to selective participation and are attenuated in within-family designs^26^.

## Divergent biomarkers

Both AN and BE PGSs showed associations with blood cell indices and liver/kidney biomarkers, suggesting some shared associations of eating disorder genetic liability with systemic physiology. The two PGSs were associated with different hormones: PGS_AN_ was positively associated with sex hormone-binding globulin, whereas PGS_BE_ was negatively associated with testosterone and IGF-1. PGS_AN_ was also negatively associated with fatty acids, whereas PGS_BE_ showed no significant associations. PGS_AN_ was negatively associated with renal/electrolyte measures and C-reactive protein (CRP), whereas PGS_BE_ showed positive associations. This aligns with evidence linking higher CRP to obesity and BED^42,43^, and lower CRP to fasting and dieting^44^, as well as a negative genetic correlation between CRP and AN (rg=-0.29)^45^. Researchers have implicated inflammatory biomarkers in homeostatic and hedonic eating regulation, as well as psychosocial risk factors for BE such as distress and inhibitory control deficits^46^. However, CRP is positively correlated with fat mass and body weight, meaning differences between AN and BE may partly reflect adiposity^46^.

Importantly, the negative association between PGS_AN_ and CRP persisted after adjusting for BMI and PGS_BMI_, suggesting it is not fully explained by body weight. This raises the possibility that immune and inflammatory processes are linked to AN genetic liability beyond low adiposity^46^. In contrast, controlling for BMI and PGS_BMI_ revealed a negative association between PGS_BE_ and CRP, suggesting that the positive association may be largely accounted for by BMI and genetic liability to higher BMI. Conditioning on measured BMI may introduce bias if BMI is a mediator or is influenced by other factors related to both PGS_BE_ and CRP. However, PGS_BMI_ is not subject to these biases. Overall, our findings suggest that biochemical markers may be informative in genetic analyses of eating disorders, although further work is needed to establish whether these associations reflect causal processes.

## Role of BMI and PGS_BMI_

As expected, most associations with anthropometric traits, as well as endocrine, nutritional, metabolic, and musculoskeletal outcomes, were substantially attenuated to non-significance after controlling for BMI, consistent with previous research^10^. In contrast, associations with lifestyle, psychiatric, and psychological traits were largely unaffected, suggesting these associations are not explained by BMI. Notably, associations with PGS_AN_ appeared less influenced by BMI adjustment than those with PGS_BE_, aligning with the close phenotypic overlap between binge-type eating disorders and obesity-related metabolic dysfunction^47^. PGS_AN_ associations may involve other processes, e.g., altered hormonal regulation, energy balance, or inflammatory mechanisms, however these cannot be inferred from these data. Analyses adjusting for PGS_BMI_ produced similar but weaker patterns compared to measured BMI, likely because measured BMI captures additional variance from environmental, non-additive genetic, and developmental factors. Again, PGS_AN_ was less impacted than PGS_BE_, consistent with a stronger role of PGS_BMI_ in PGS_BE_ associations. These findings underscore the importance of considering both measured and genetic BMI in analyses of eating disorder genetic liability, as disentangling BMI-dependent and independent effects may clarify mechanisms underlying somatic comorbidities.

## Strengths and limitations

Where known, we excluded all UK Biobank participants with AN or BE, strengthening inference that associations reflect genetic liability rather than downstream consequences of the traits. Nonetheless, conditioning on eating disorder status may introduce collider bias, as individuals with high genetic liability who do not develop AN or BE may possess protective factors reflected in the traits examined. Further, only a subset of UK Biobank completed the mental health questionnaire^19^ and therefore participants with eating disorder symptoms likely remain in our sample and may bias results. More generally, population-based PGS associations may reflect indirect genetic effects^6^, assortative mating^7^, residual population stratification, and selective participation^8^ as well as pleiotropy. These processes are likely to affect behavioural, cognitive, and psychiatric traits more than anthropometric traits, and within-family designs will be needed to distinguish them. UKB’s healthy volunteer bias limits generalisability to the wider UK population and beyond; applying sampling weights partially addressed this by aligning the analytic sample with the source population. Associations with psychiatric and somatic traits require cautious interpretation, as these domains are particularly vulnerable to participation bias. There is substantial genetic overlap between UK Biobank participation and traits such as ADHD and asthma^8^, likely reflecting participation-related behaviours. Further, while we describe differences between PGS_AN_ and PGS_BE_, these scores differ in statistical power (AN=24,223 cases, 1,243,971 controls; BE=39,279 cases, 1,227,436 controls) and capture distinct constructs (disorder versus symptom). These phenotypes are also not entirely distinct: AN includes a binge-eating/purging subtype, and the two are positively genetically correlated (rg = 0.46)^3^. Some shared associations may therefore reflect overlapping phenotypes as well as shared genetic liability. Moreover, PGS_BE_ was derived from a broad definition of binge eating that did not require a minimum frequency or duration of episodes^3^. Therefore, PGS_BE_ represents liability to a heterogeneous behavioural construct and may not generalise to BED or BN. Further, the discovery GWASs were predominantly female (94% of AN cases and 96% of BE-BROAD cases). If genetic influences on eating disorders differ by sex, the scores may capture liability less accurately in males. Termorshuizen et al.^3^ found that PGSs derived from female-only GWASs were positively associated with eating disorder status in males (BE: OR 1.06-1.20, AN: 1.07-1.41), although not consistently significantly across cohorts. Our models adjusted for sex but did not test whether associations differed by sex, so associations with male-specific traits, such as pubertal markers, should be interpreted cautiously. We are also limited by reduced generalisability of our main results to genetically-inferred ancestries outside of European. The paucity of statistically significant findings in other genetic ancestries reflects the smaller sample sizes and trans-ancestry portability issues in calculating PGS. Although SBayesRC improves cross-ancestry prediction relative to earlier methods, prediction accuracy of European-derived scores in other ancestries remains substantially lower. Well-powered GWAS in ancestries other than European, combined with multi-ancestry PGS methods, are needed.

## Conclusion

Across the phenome, PGS_AN_ and PGS_BE_ showed both overlapping and distinct association patterns, reflecting shared psychiatric liability as well as divergent somatic, anthropometric, and metabolic association patterns. This divergence parallels clinically observed differences in medical sequelae between anorexia nervosa and binge-type eating disorders. We observed these associations after excluding participants with a recorded eating disorder, although subclinical disordered eating may still contribute. Our results generate new avenues for exploration; further research with well-powered GWASs, within-family designs, and causal-inference approaches is needed to disentangle these associations.

## Author Contribution

All authors contributed to the conception of the work and the interpretation of the data; HLD completed all data analysis and drafted the work; all authors reviewed the work for important intellectual content; all authors approved the final version for publication; all authors agree to be accountable for all aspects of the work in ensuring that questions related to the accuracy or integrity of any part of the work are appropriately investigated and resolved.

## Data Availability

Data supporting the results of this study are available from UK Biobank. Data are not publicly available but can be accessed by researchers with an approved UK Biobank application.

## Acknowledgements

This work was undertaken in the UK Biobank under project 82087. The views expressed in this publication are those of the authors and not necessarily those of the NHS, the NIHR or the UK Department of Health.

