## Supplementary Figures for "Phenome-wide associations of polygenic scores for anorexia nervosa and binge eating in the UK Biobank"

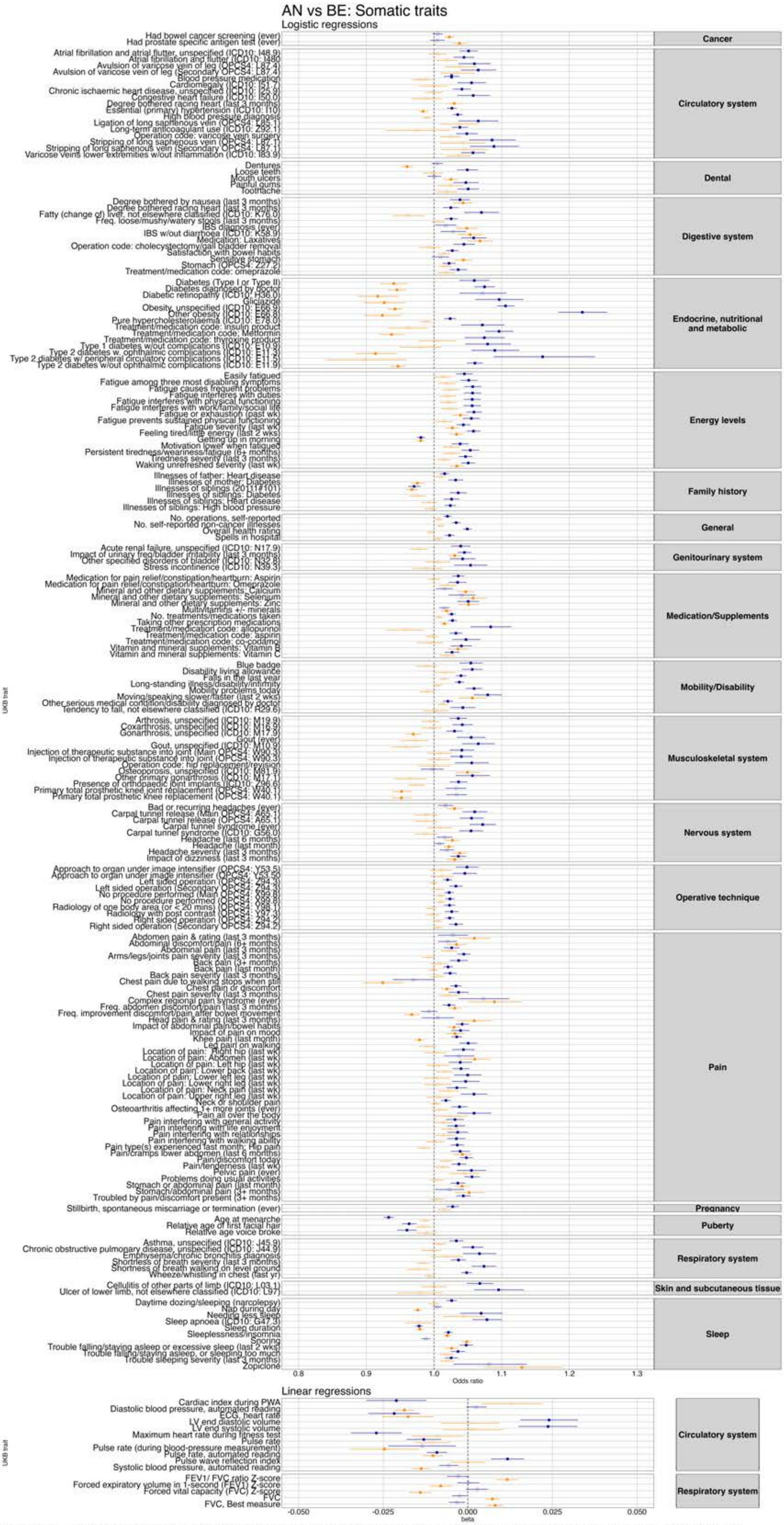


**Figure S1.** Phenome-wide association results for binge eating PGS (blue) and anorexia nervosa PGS (orange) and UK Biobank somatic traits. *Note. Filled squares=significant; Empty squares=non-significant. We have displayed all results that were significant in at least one of the pheWASs.*

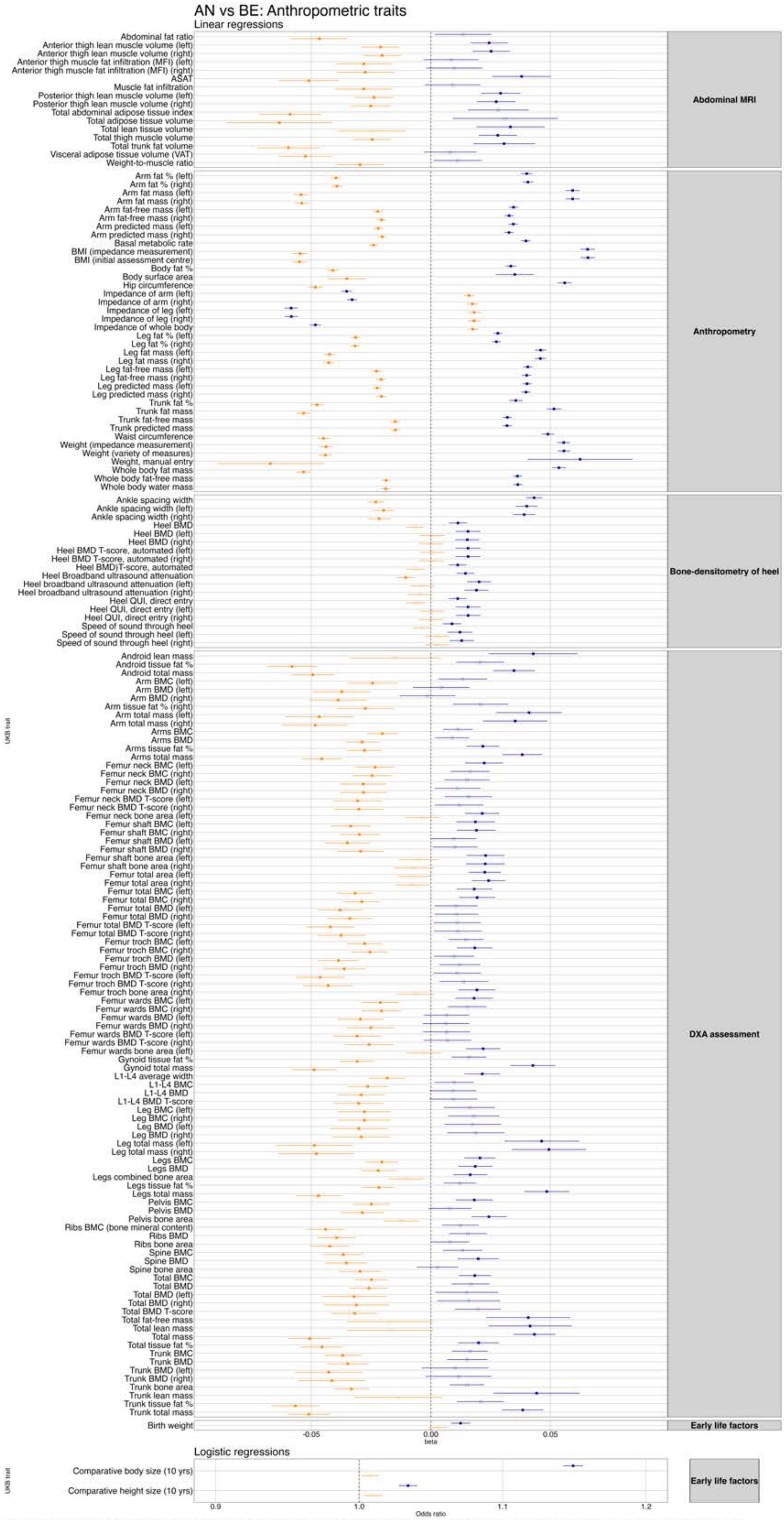


**Figure S2.** Phenome-wide association results for binge eating PGS (blue) and anorexia nervosa PGS (orange) and UK Biobank anthropometric traits. *Note. Filled squares=significant; Empty squares=non-significant. We have displayed all results that were significant in at least one of the pheWASs.*

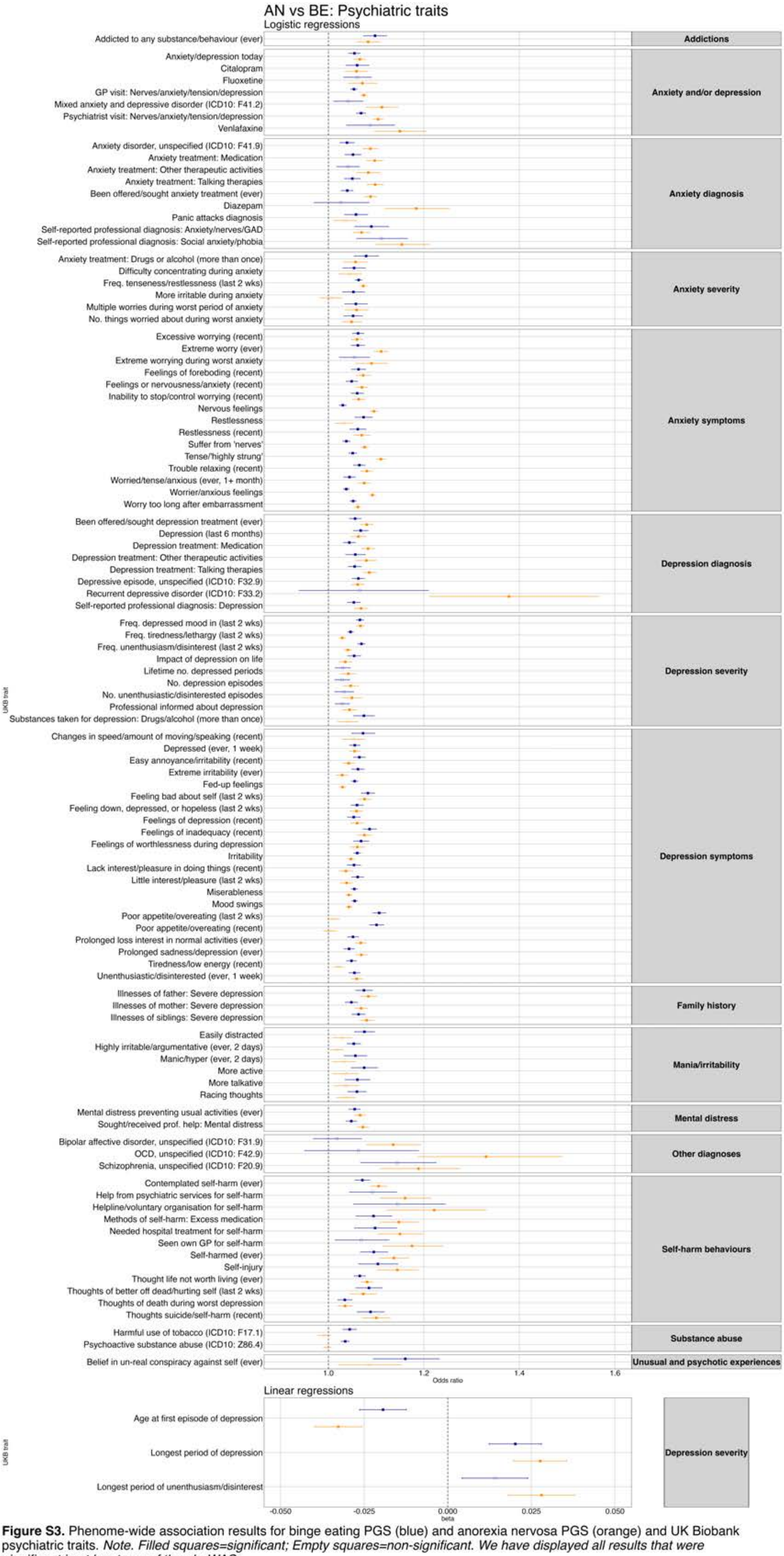


**Figure S3.** Phenome-wide association results for binge eating PGS (blue) and anorexia nervosa PGS (orange) and UK Biobank psychiatric traits. *Note. Filled squares=significant; Empty squares=non-significant. We have displayed all results that were significant in at least one of the pheWASs.*

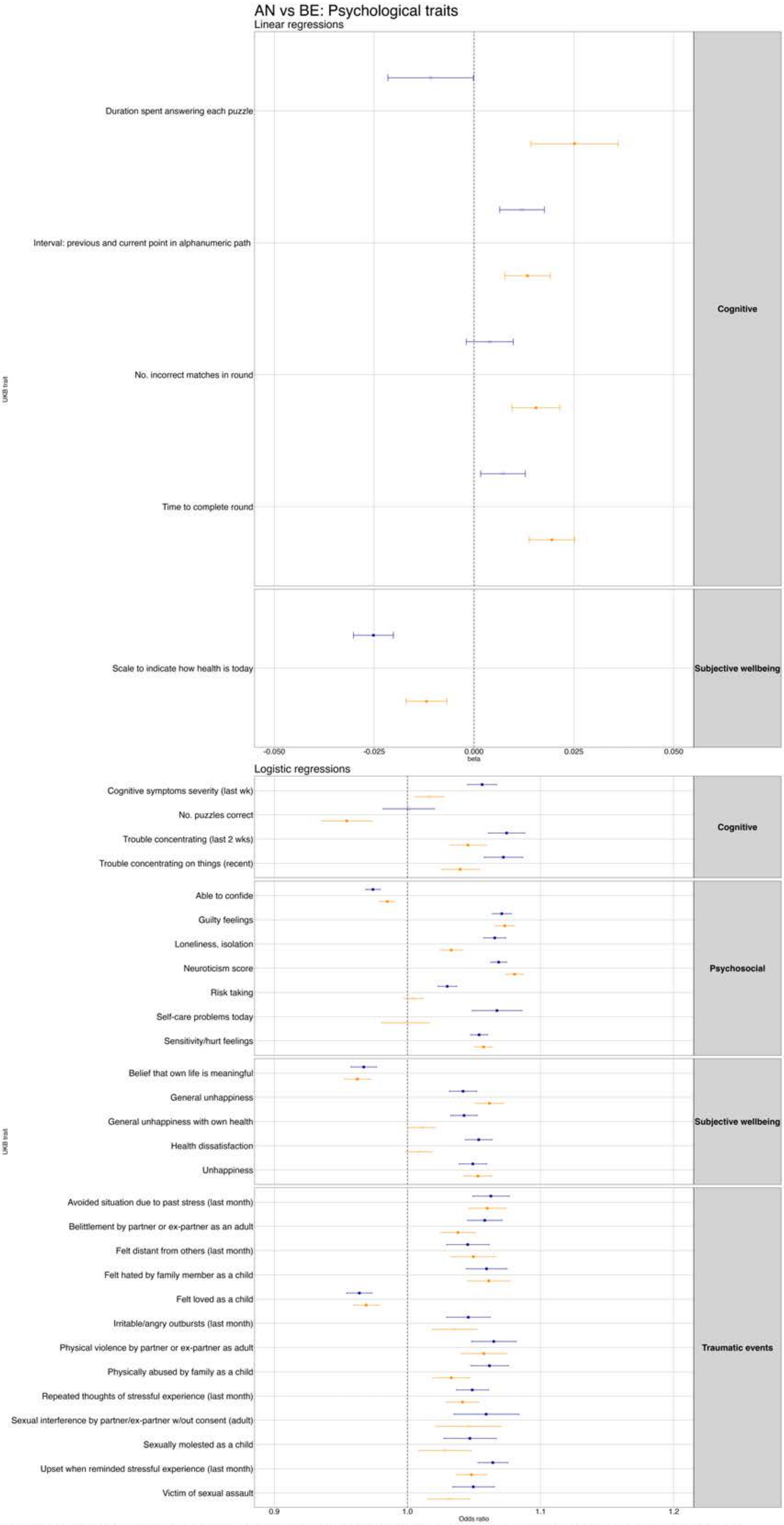


**Figure S4.** Phenome-wide association results for binge eating PGS (blue) and anorexia nervosa PGS (orange) and UK Biobank psychological traits. *Note. Filled squares=significant; Empty squares=non-significant. We have displayed all results that were significant in at least one of the pheWASs.*

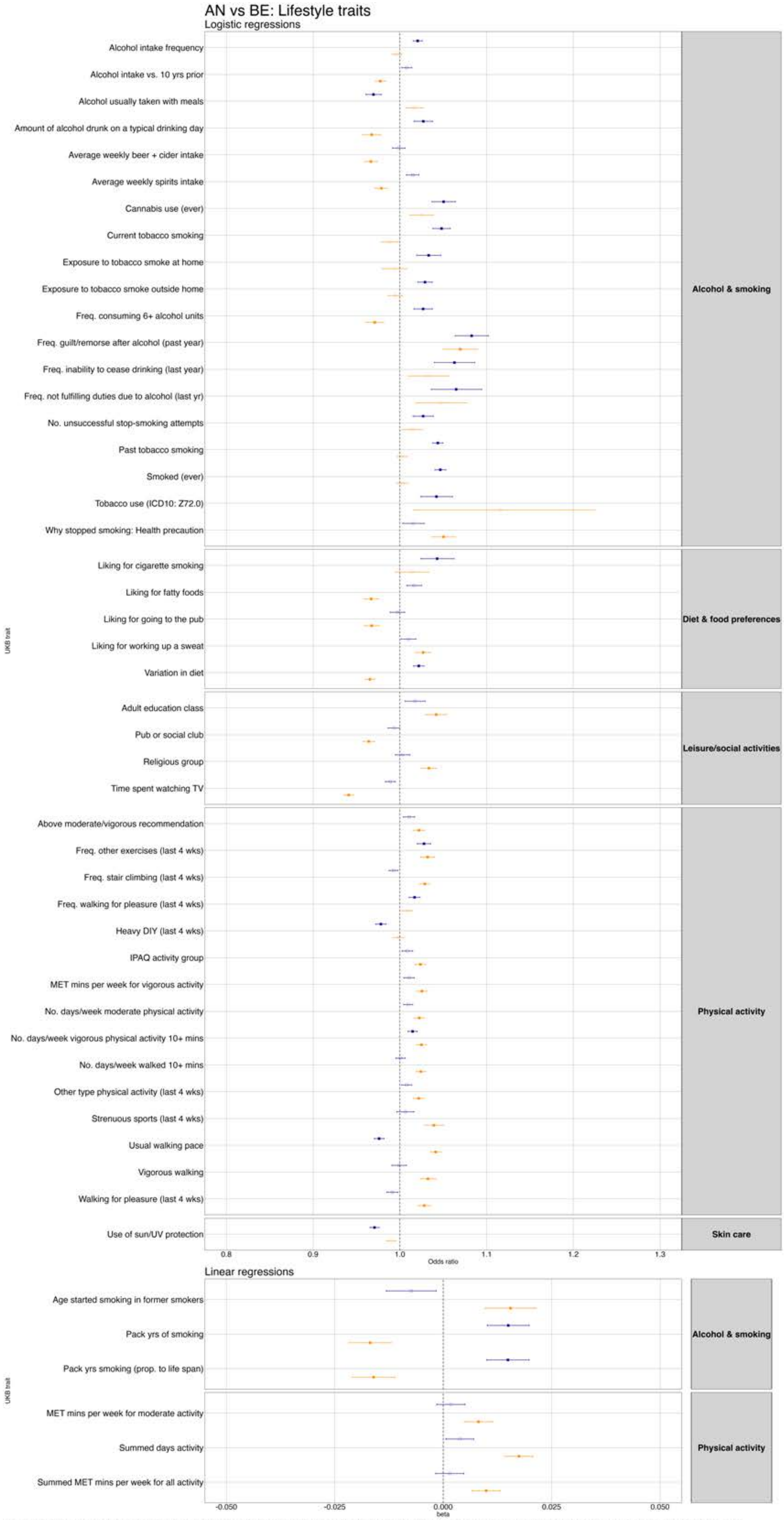


**Figure S5.** Phenome-wide association results for binge eating PGS (blue) and anorexia nervosa PGS (orange) and UK Biobank lifestyle traits. *Note. Filled squares=significant; Empty squares=non-significant. We have displayed all results that were significant in at least one of the pheWASs.*

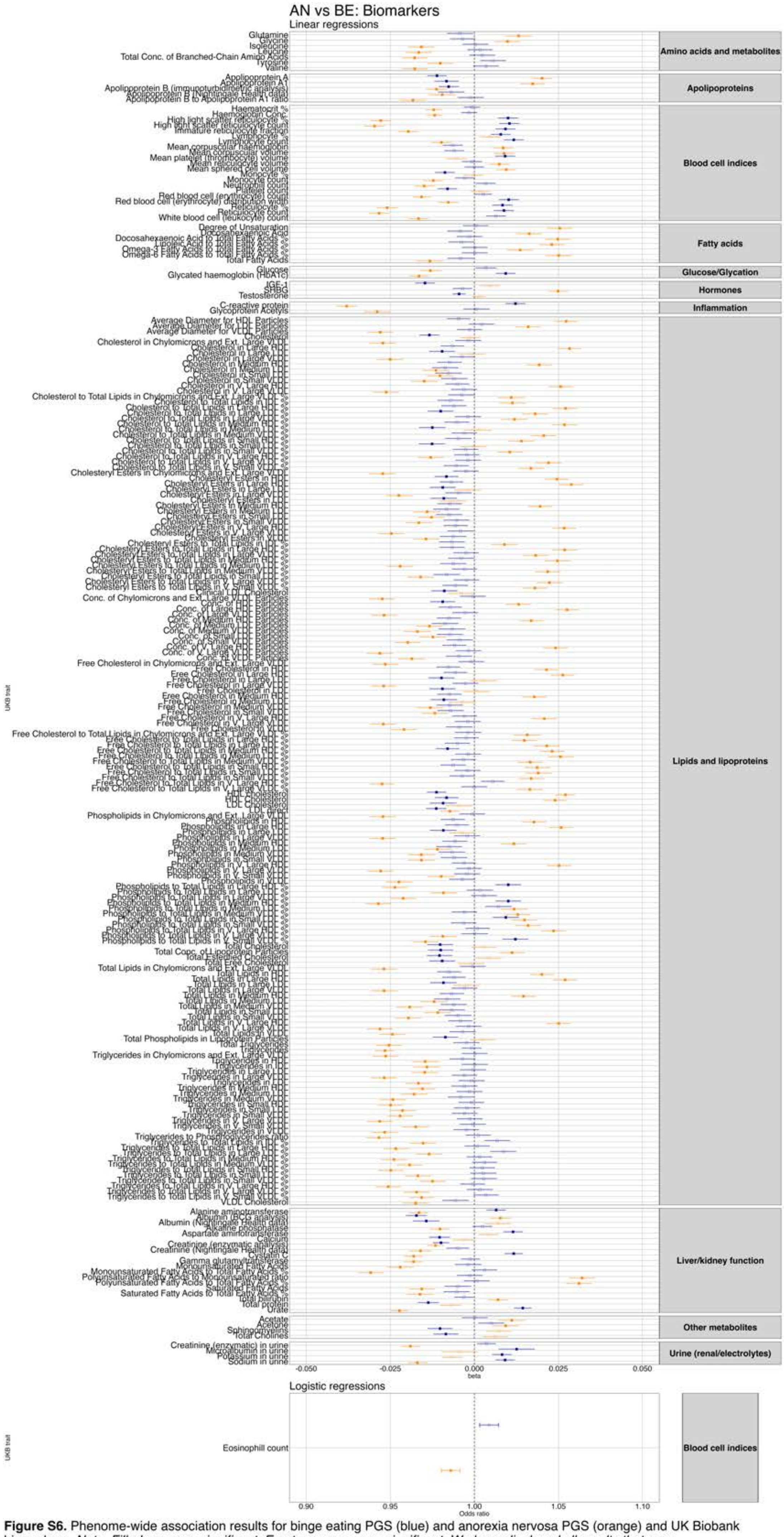


**Figure S6.** Phenome-wide association results for binge eating PGS (blue) and anorexia nervosa PGS (orange) and UK Biobank biomarkers. *Note. Filled squares=significant; Empty squares=non-significant. We have displayed all results that were significant in at least one of the pheWASs.*

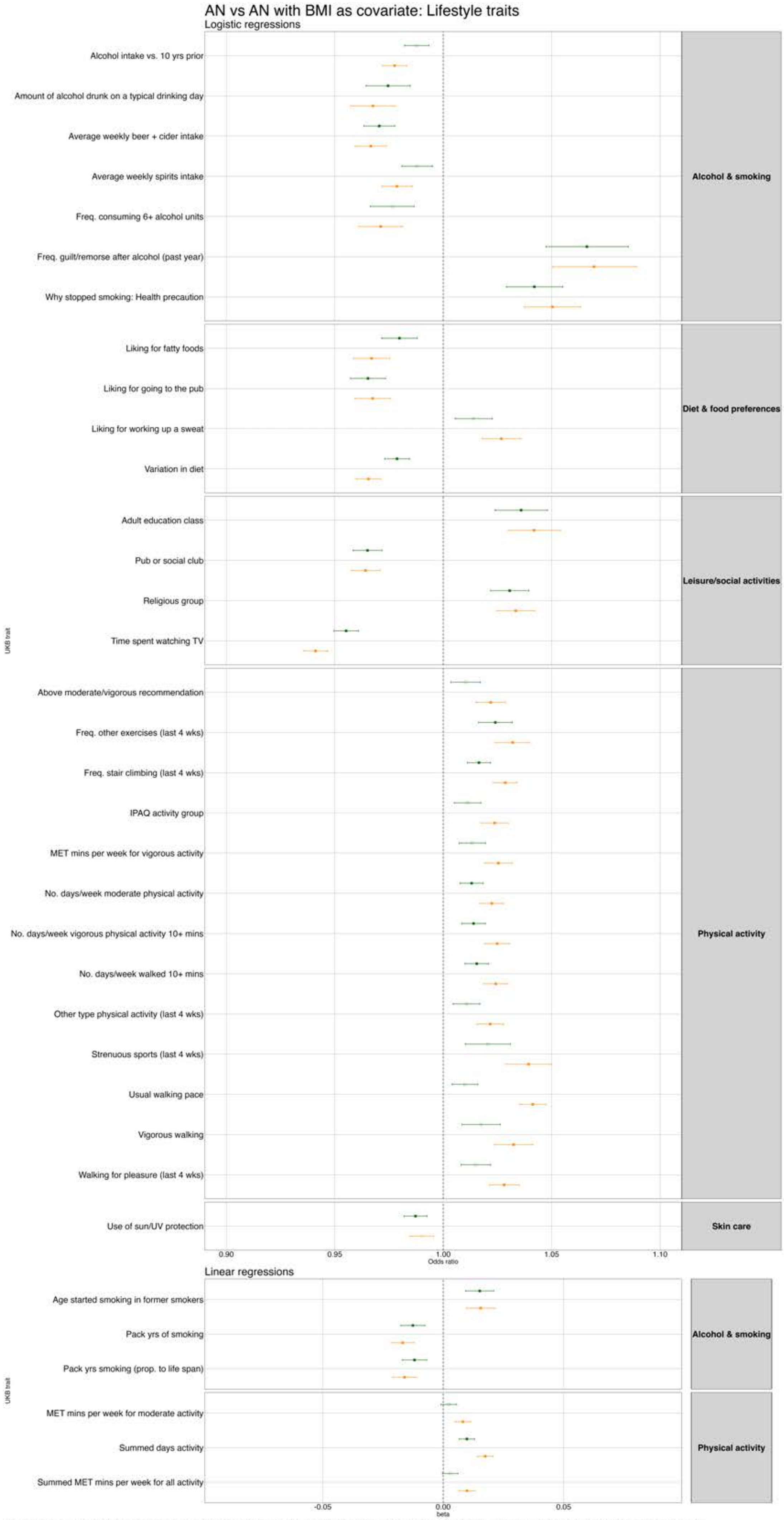


**Figure S7.** Phenome-wide association results between the anorexia nervosa PGS and UK Biobank lifestyle traits from the main analysis (orange) and after controlling for BMI (green). *Note. Filled squares=significant; Empty squares=non-significant. We have displayed all results that were significant in at least one of the pheWASs.*

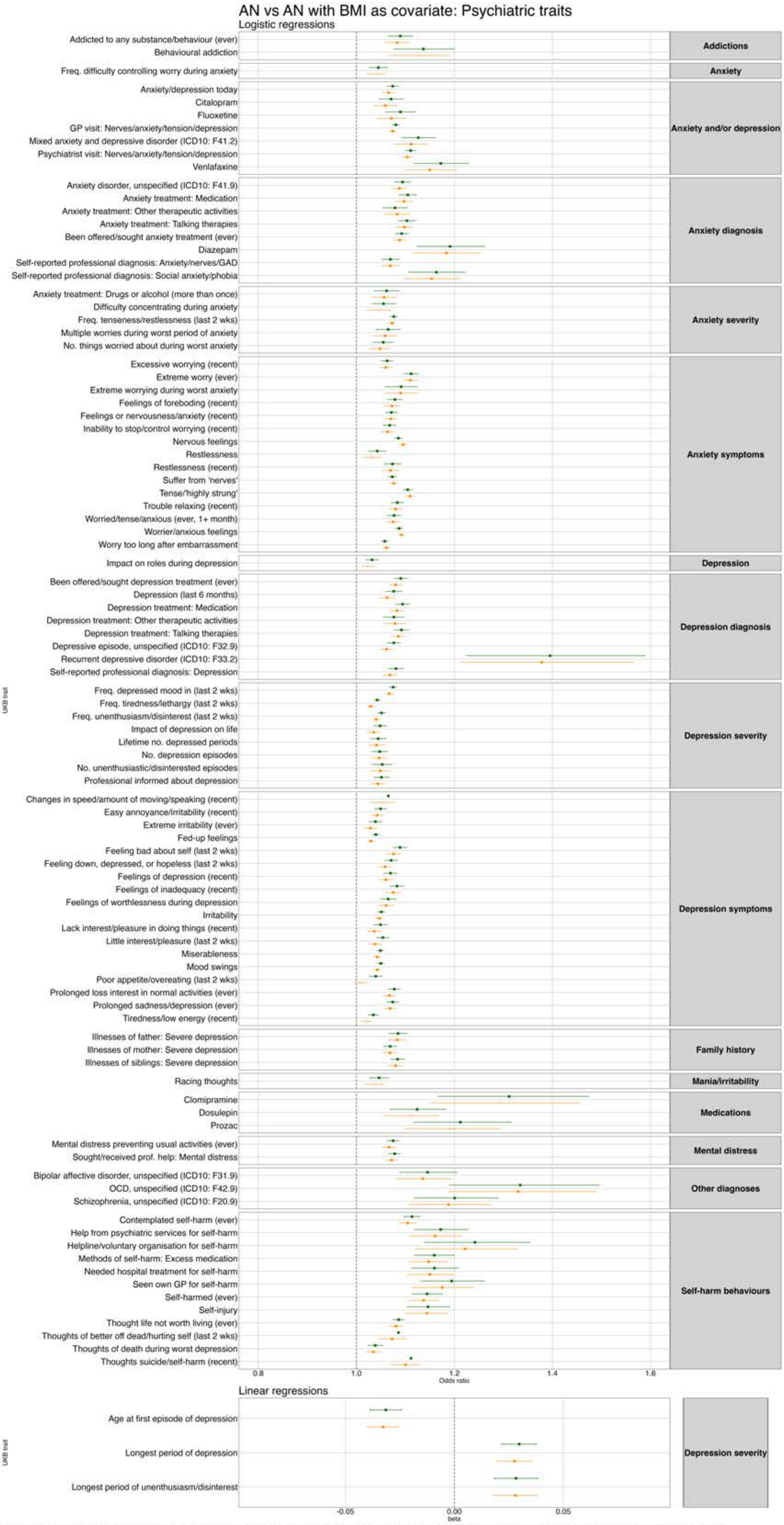


**Figure S9.** Phenome-wide association results between the anorexia nervosa PGS and UK Biobank psychiatric traits from the main analysis (orange) and after controlling for BMI (green). *Note. Filled squares=significant; Empty squares=non-significant. We have displayed all results that were significant in at least one of the pheWASs.*

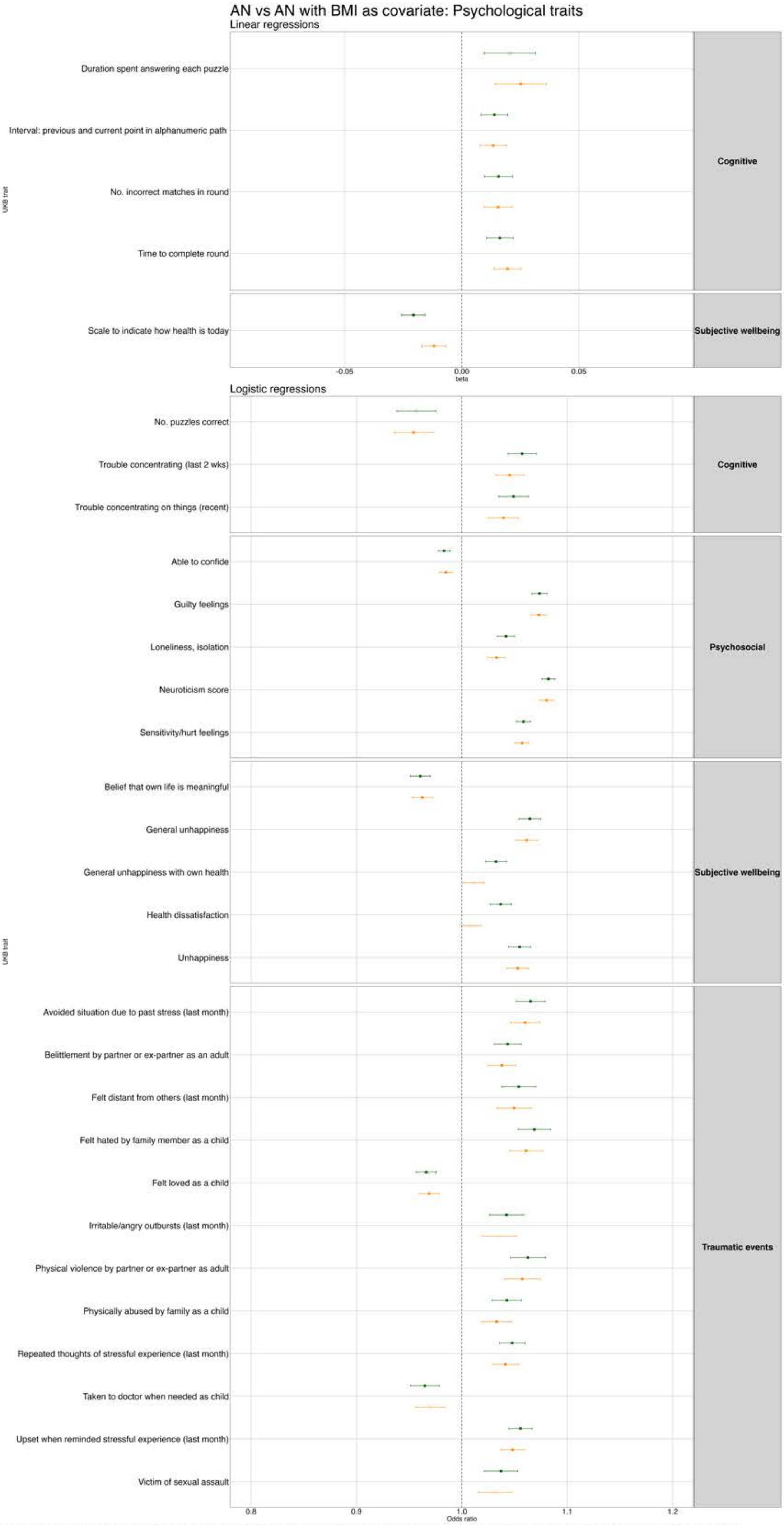


**Figure S9.** Phenome-wide association results between the anorexia nervosa PGS and UK Biobank psychological traits from the main analysis (orange) and after controlling for BMI (green). *Note. Filled squares=significant; Empty squares=non-significant. We have displayed all results that were significant in at least one of the pheWASs.*

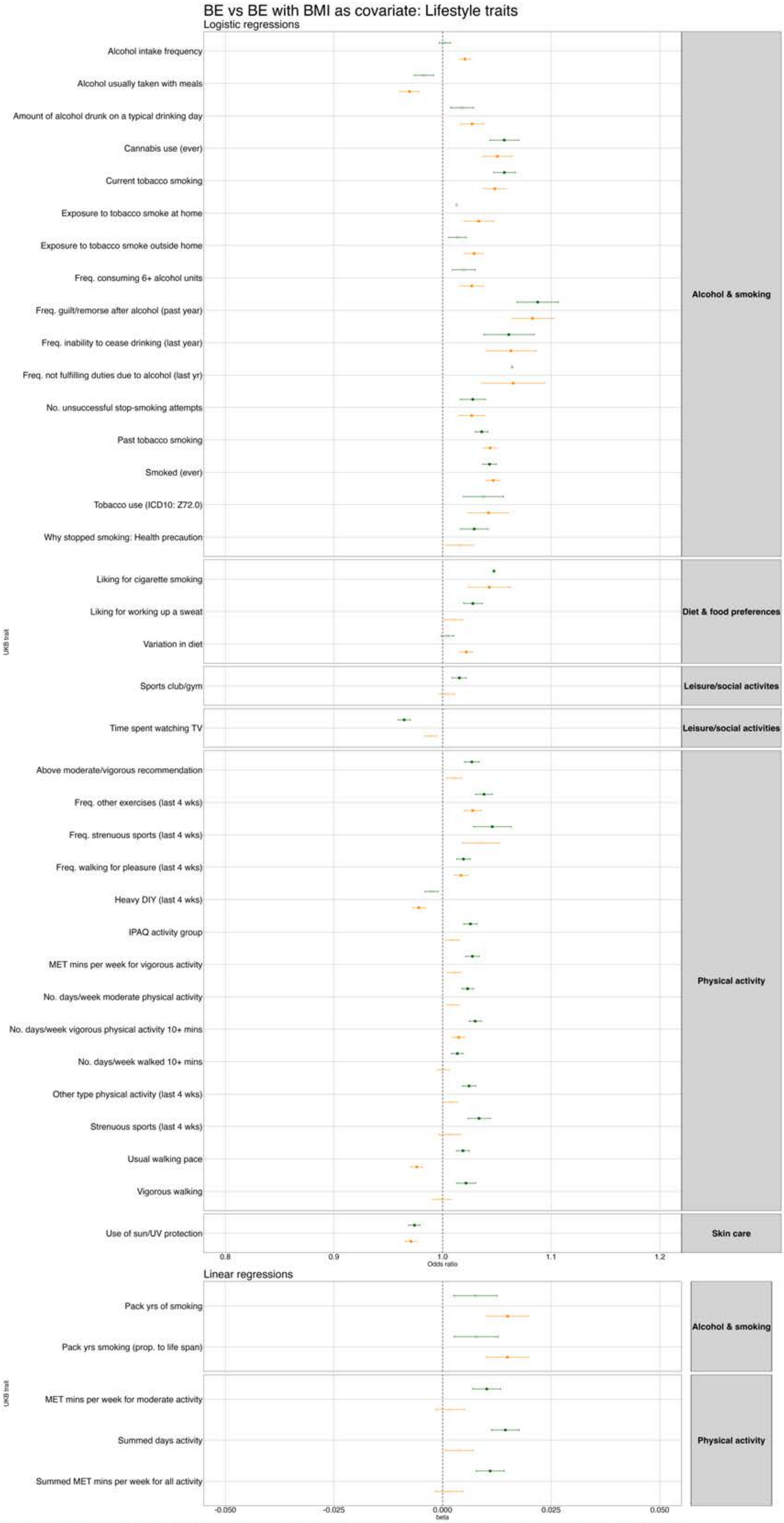


**Figure S10.** Phenome-wide association results between the binge eating PGS and UK Biobank lifestyle traits from the main analysis (orange) and after controlling for BMI (green). *Note. Filled squares=significant; Empty squares=non-significant. We have displayed all results that were significant in at least one of the pheWASs.*

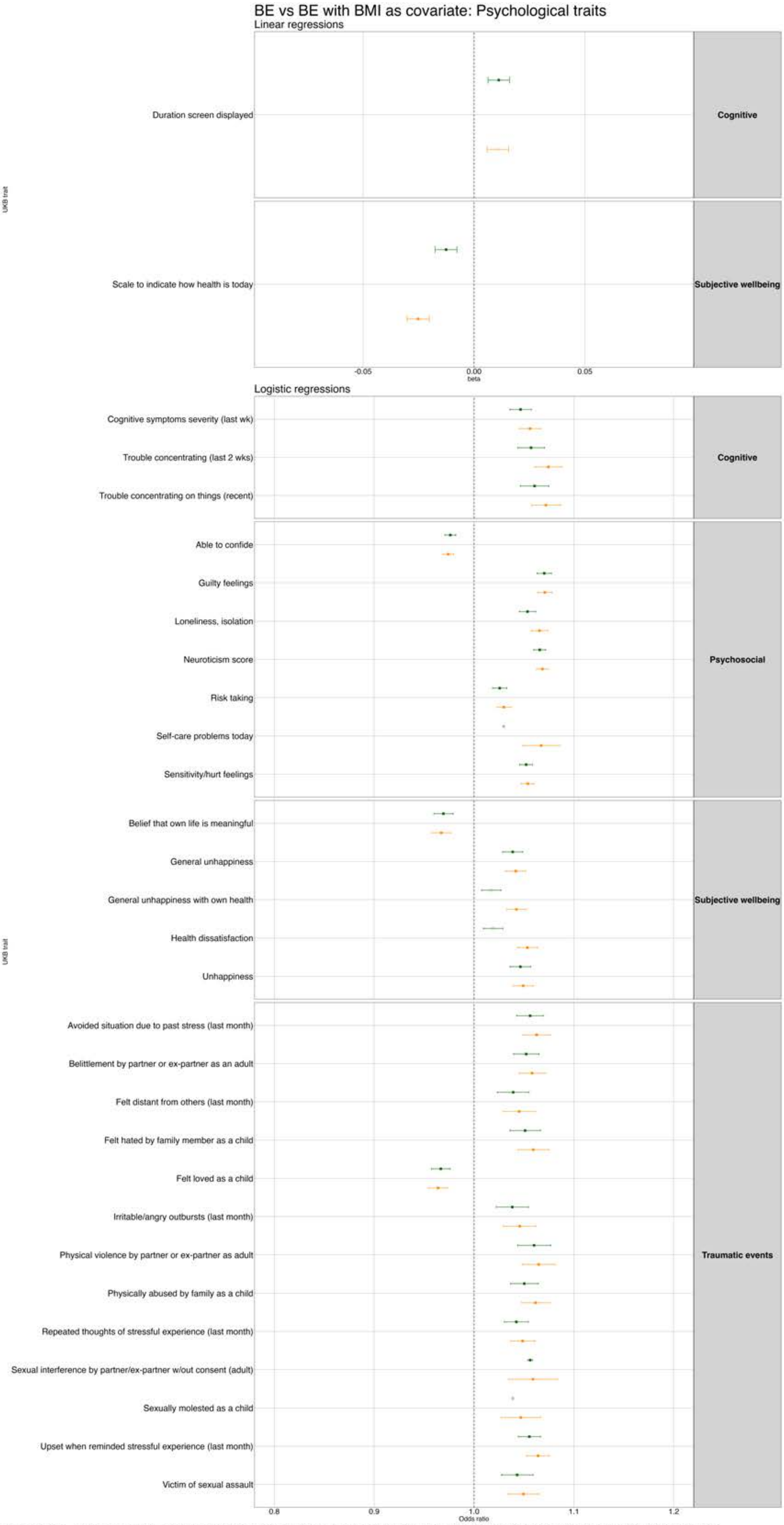


**Figure S11.** Phenome-wide association results between the binge eating PGS and UK Biobank psychological traits from the main analysis (orange) and after controlling for BMI (green). *Note. Filled squares=significant; Empty squares=non-significant. We have displayed all results that were significant in at least one of the pheWASs.*

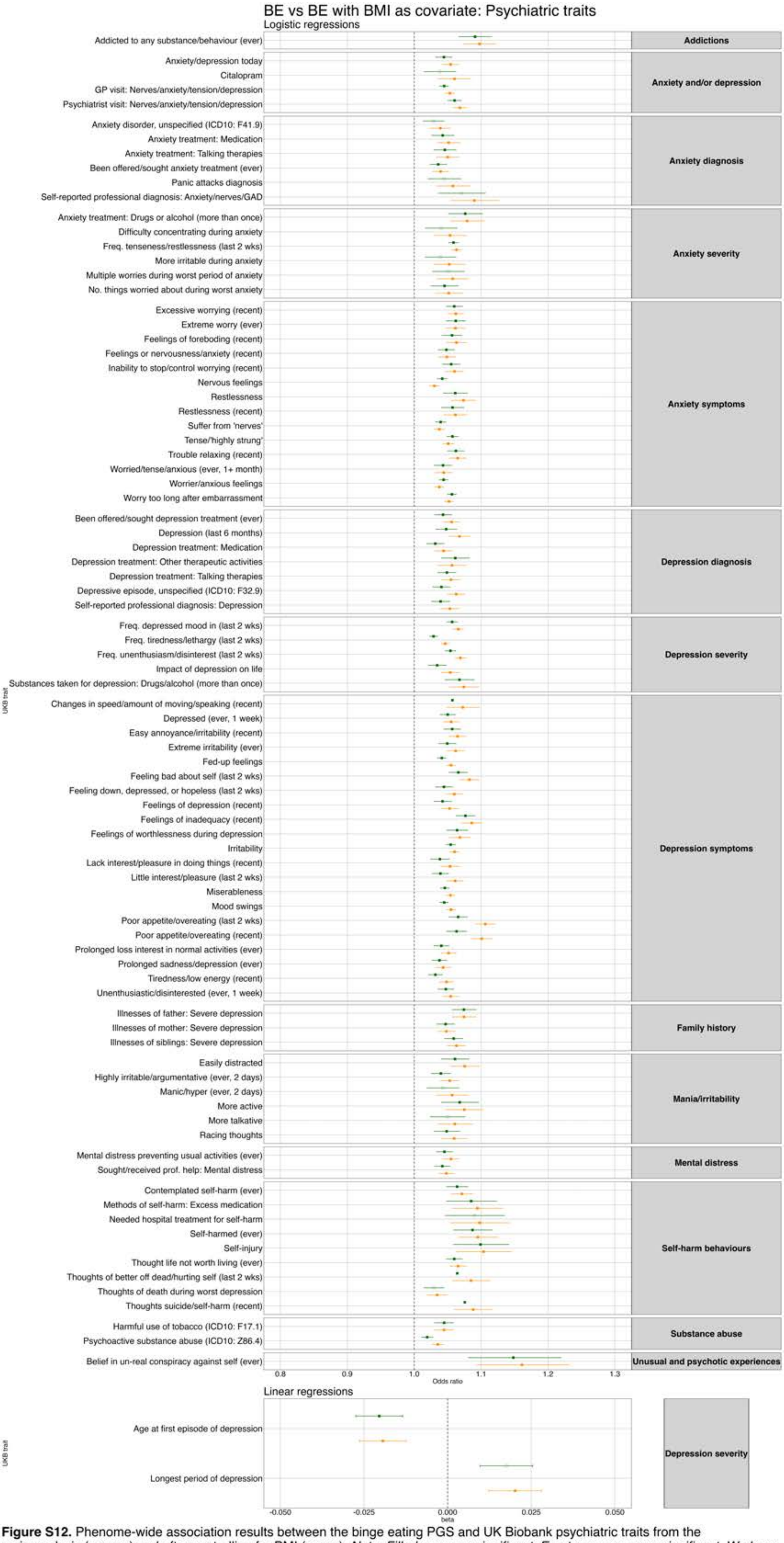


**Figure S12.** Phenome-wide association results between the binge eating PGS and UK Biobank psychiatric traits from the main analysis (orange) and after controlling for BMI (green). *Note. Filled squares=significant; Empty squares=non-significant. We have displayed all results that were significant in at least one of the pheWASs.*

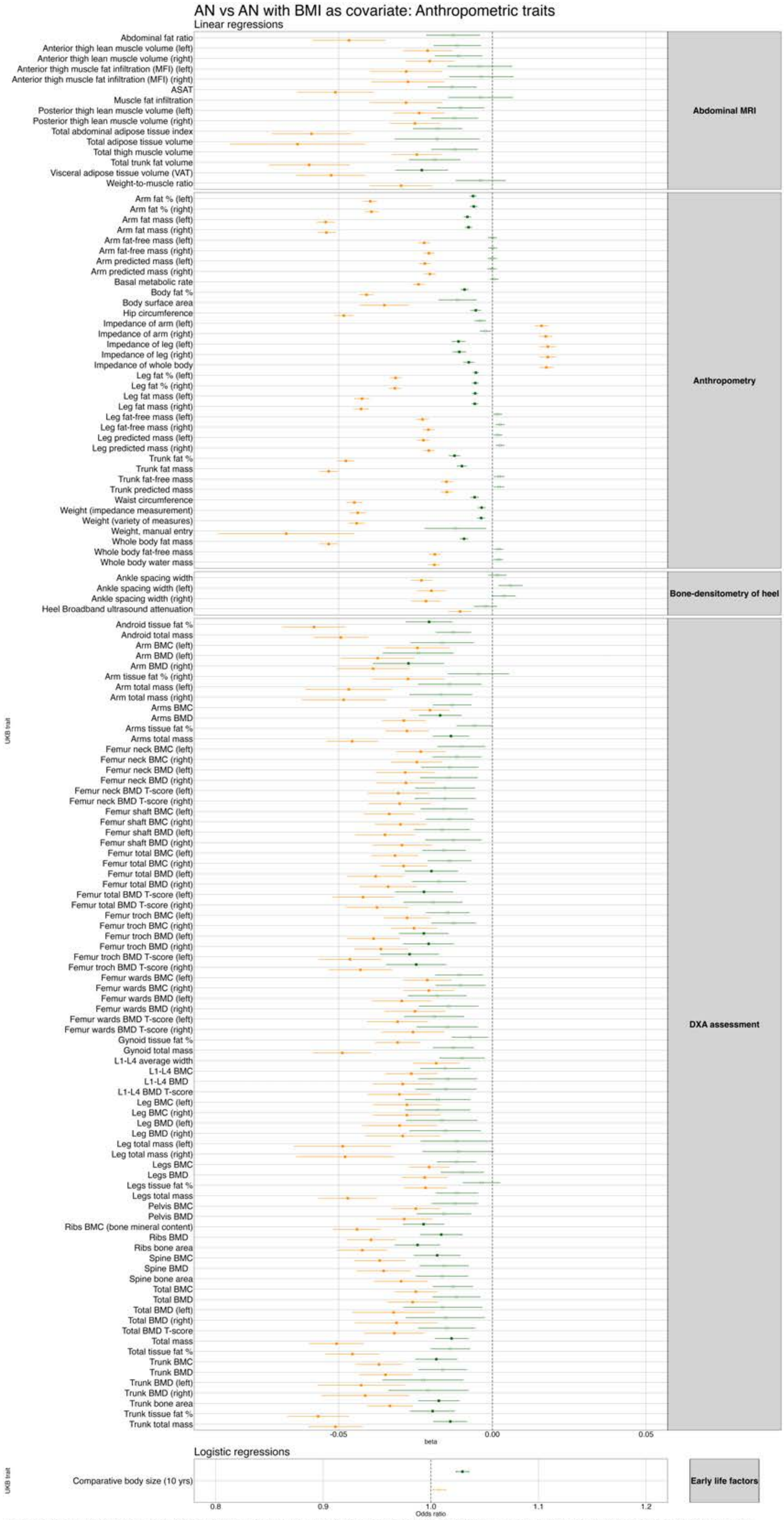


**Figure S13.** Phenome-wide association results between the anorexia nervosa PGS and UK Biobank anthropometric traits from the main analysis (orange) and after controlling for BMI (green). *Note. Filled squares=significant; Empty squares=non-significant. We have displayed all results that were significant in at least one of the pheWASs.*

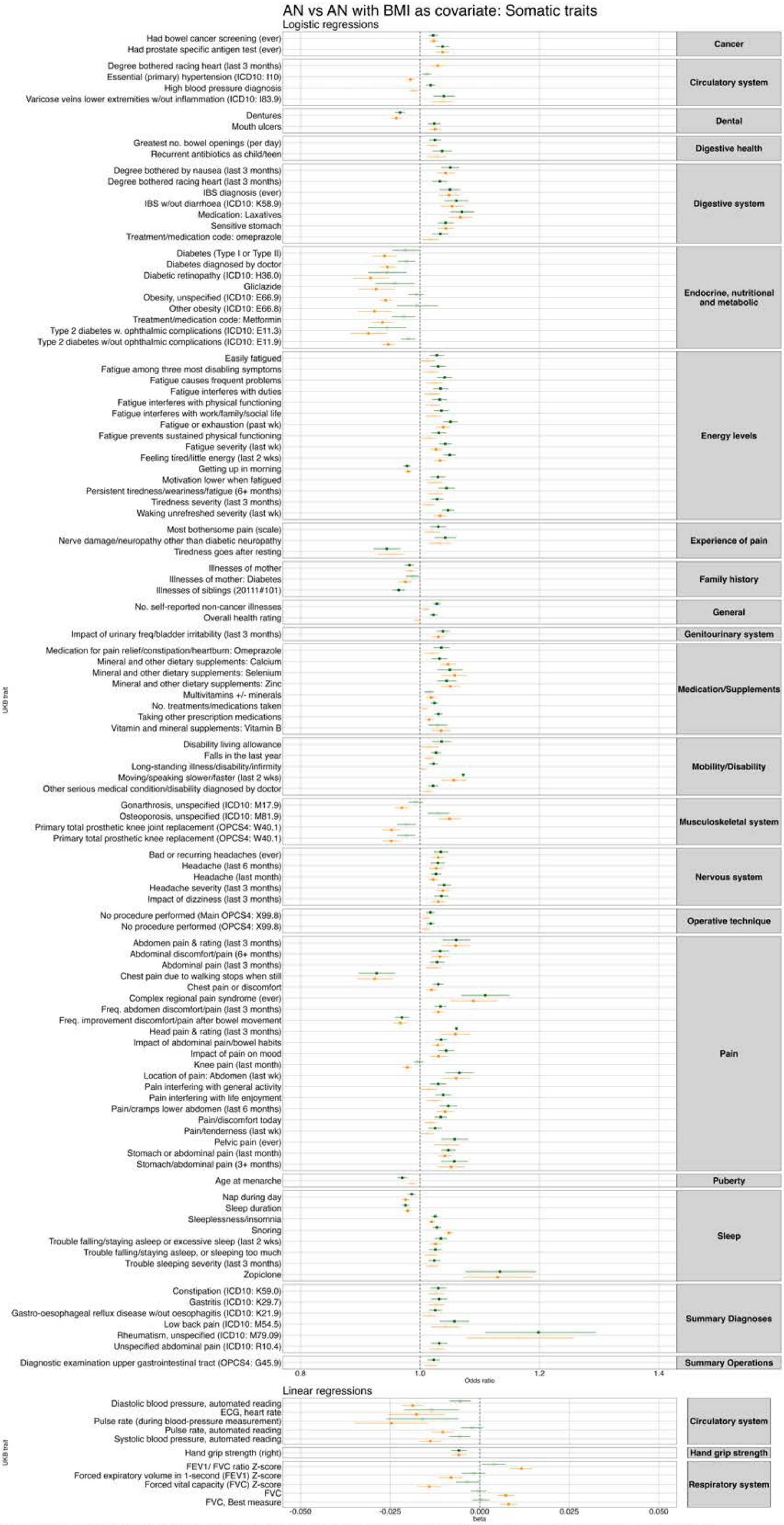


**Figure S14.** Phenome-wide association results between the anorexia nervosa PGS and UK Biobank somatic traits from the main analysis (orange) and after controlling for BMI (green). *Note. Filled squares=significant; Empty squares=non-significant. We have displayed all results that were significant in at least one of the pheWASs.*

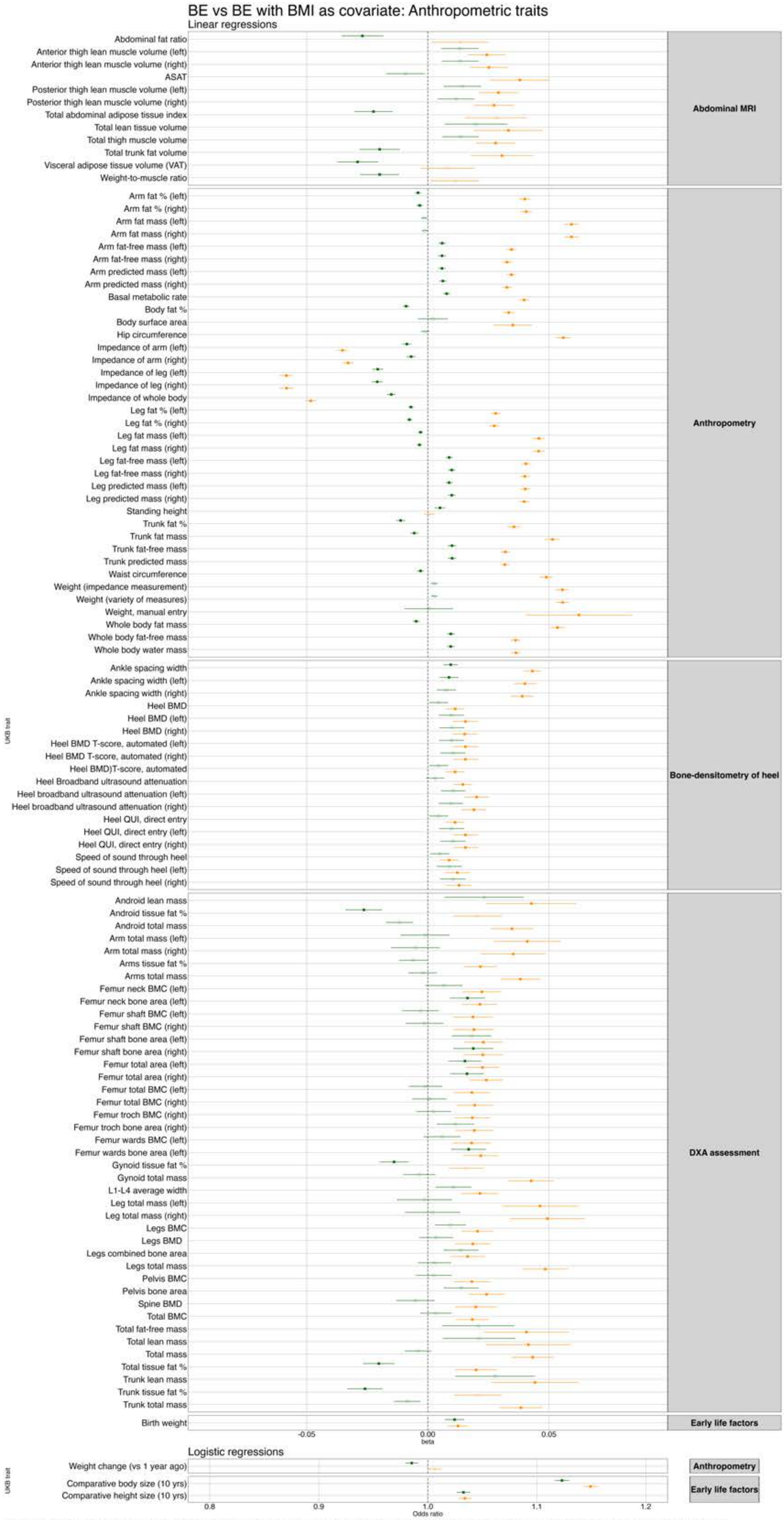


**Figure S15.** Phenome-wide association results between the binge eating PGS and UK Biobank anthropometric traits from the main analysis (orange) and after controlling for BMI (green). *Note. Filled squares=significant; Empty squares=non-significant. We have displayed all results that were significant in at least one of the pheWASs.*

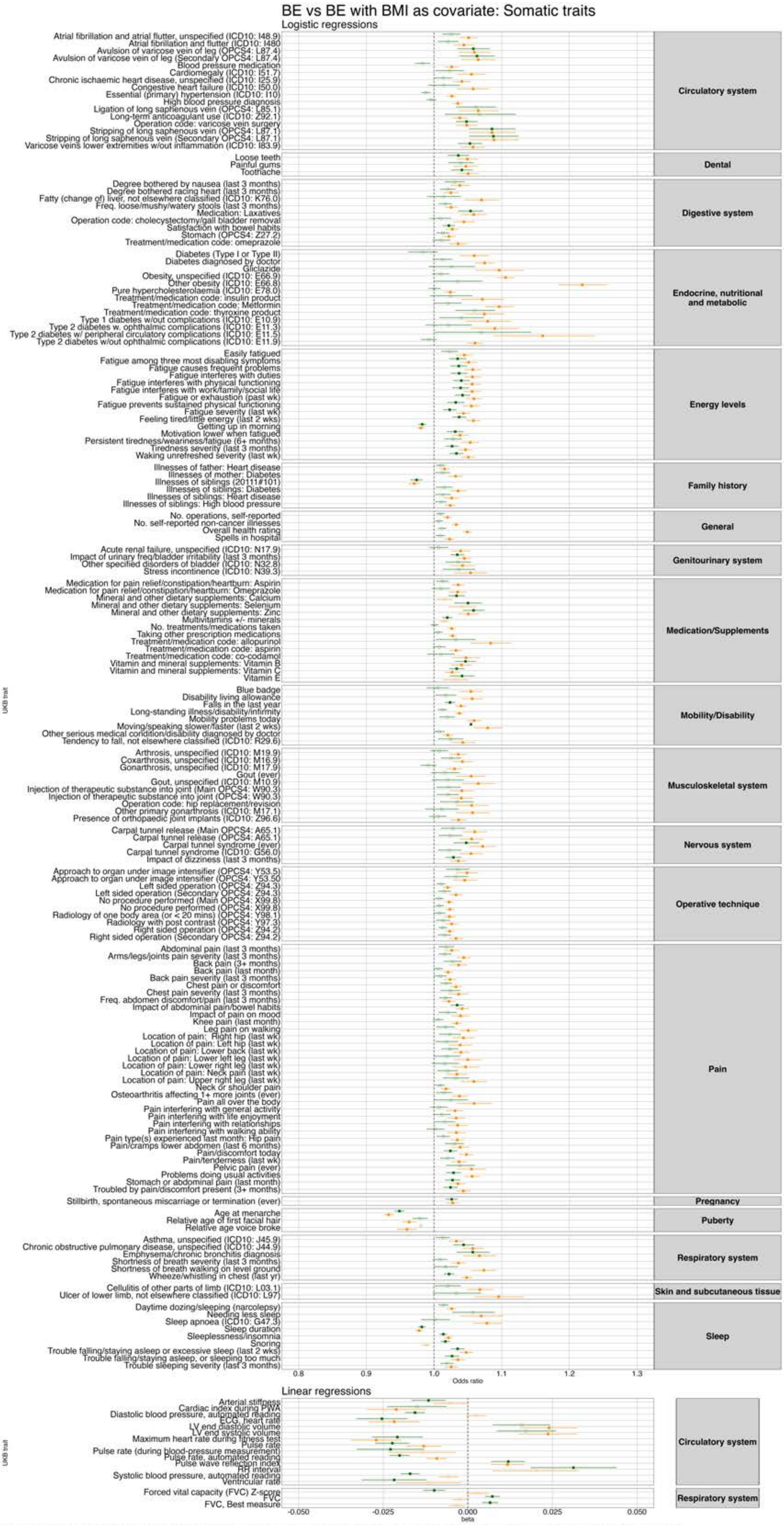


**Figure S16.** Phenome-wide association results between the binge eating PGS and UK Biobank somatic traits from the main analysis (orange) and after controlling for BMI (green). *Note. Filled squares=significant; Empty squares=non-significant. We have displayed all results that were significant in at least one of the pheWASs.*

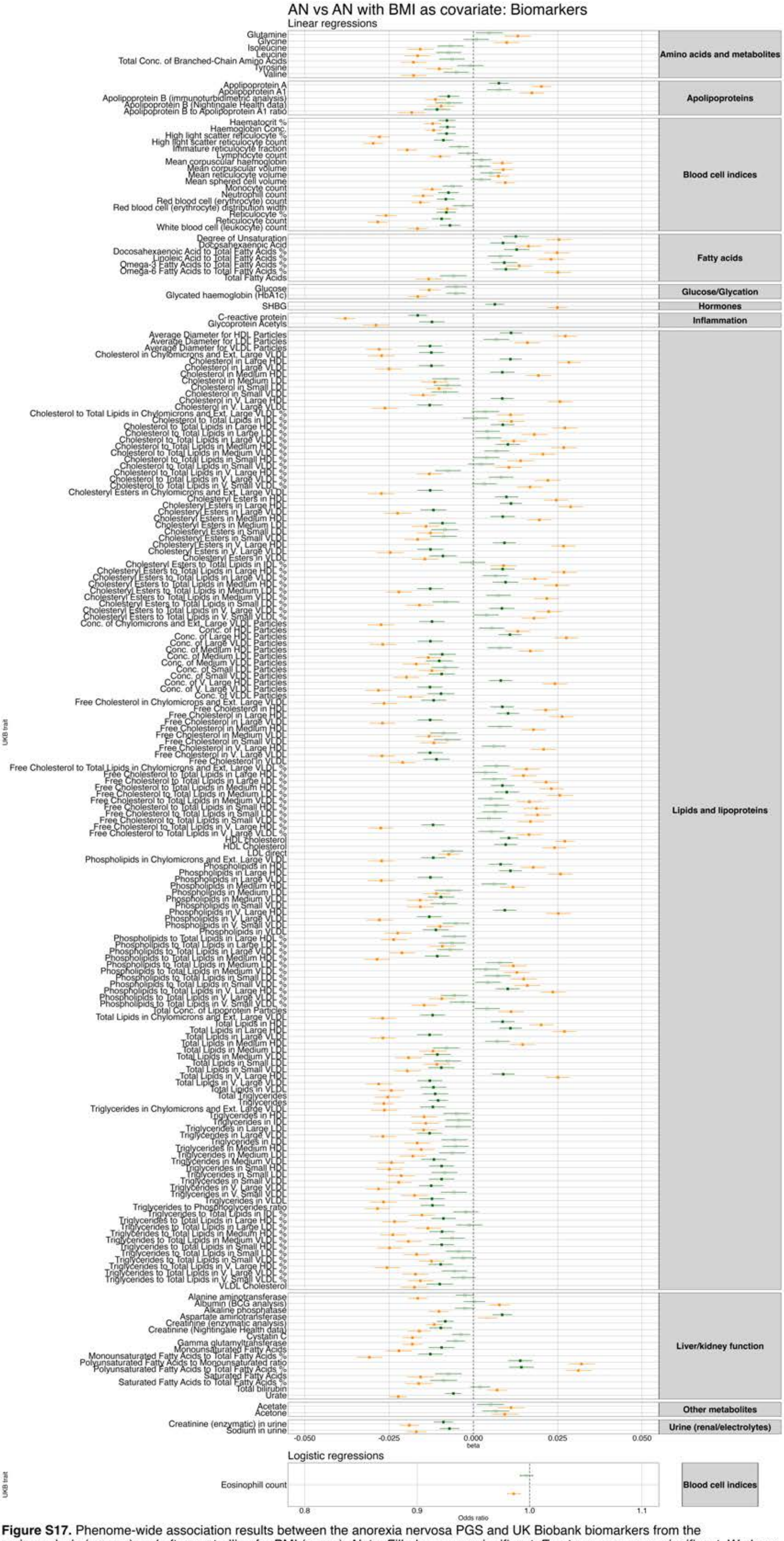


**Figure S17.** Phenome-wide association results between the anorexia nervosa PGS and UK Biobank biomarkers from the main analysis (orange) and after controlling for BMI (green). *Note. Filled squares=significant; Empty squares=non-significant. We have displayed all results that were significant in at least one of the pheWASs.*

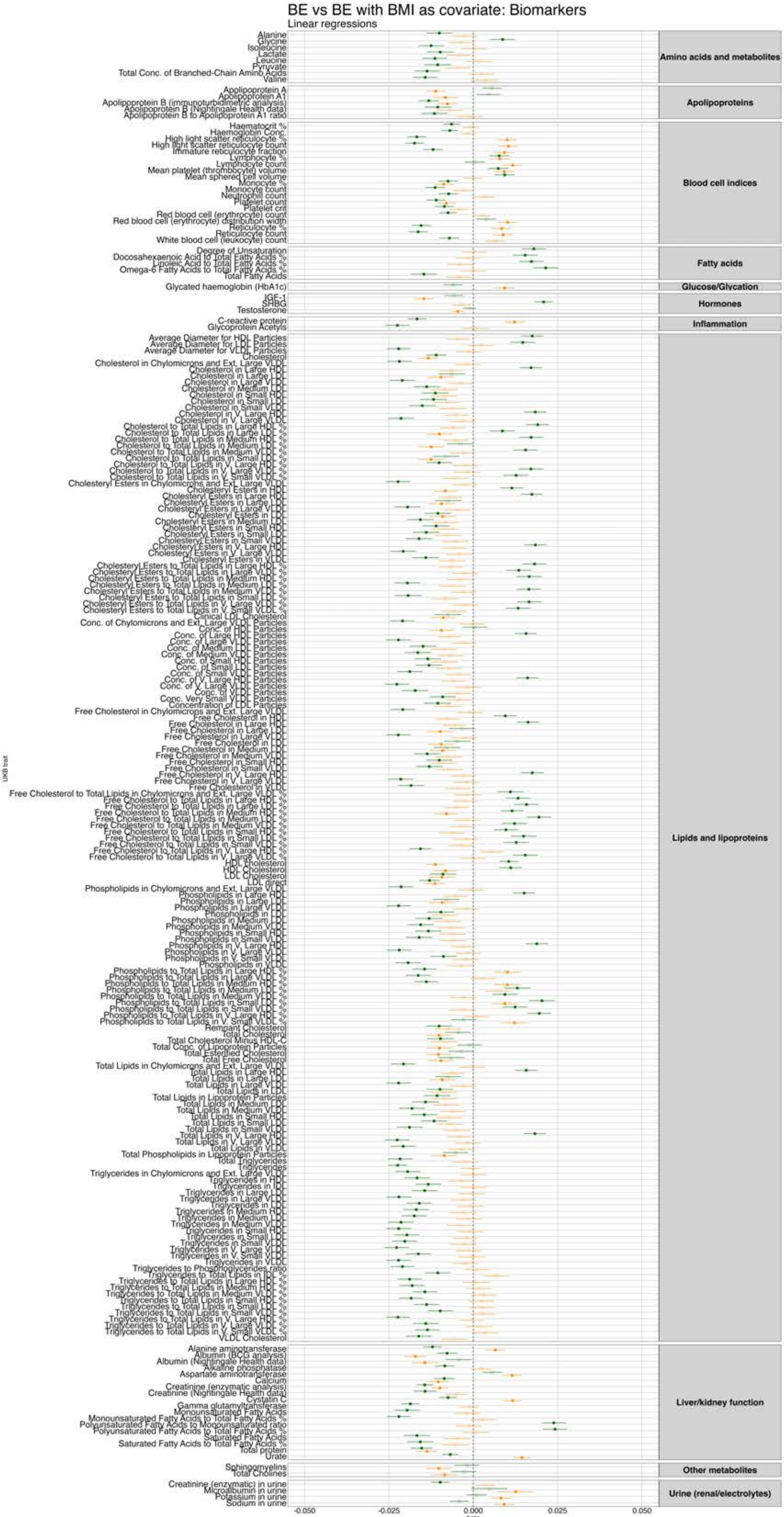


**Figure S18.** Phenome-wide association results between the binge eating PGS and UK Biobank biomarkers from the main analysis (orange) and after controlling for BMI (green). *Note. Filled squares=significant; Empty squares=non-significant. We have displayed all results that were significant in at least one of the pheWASs.*

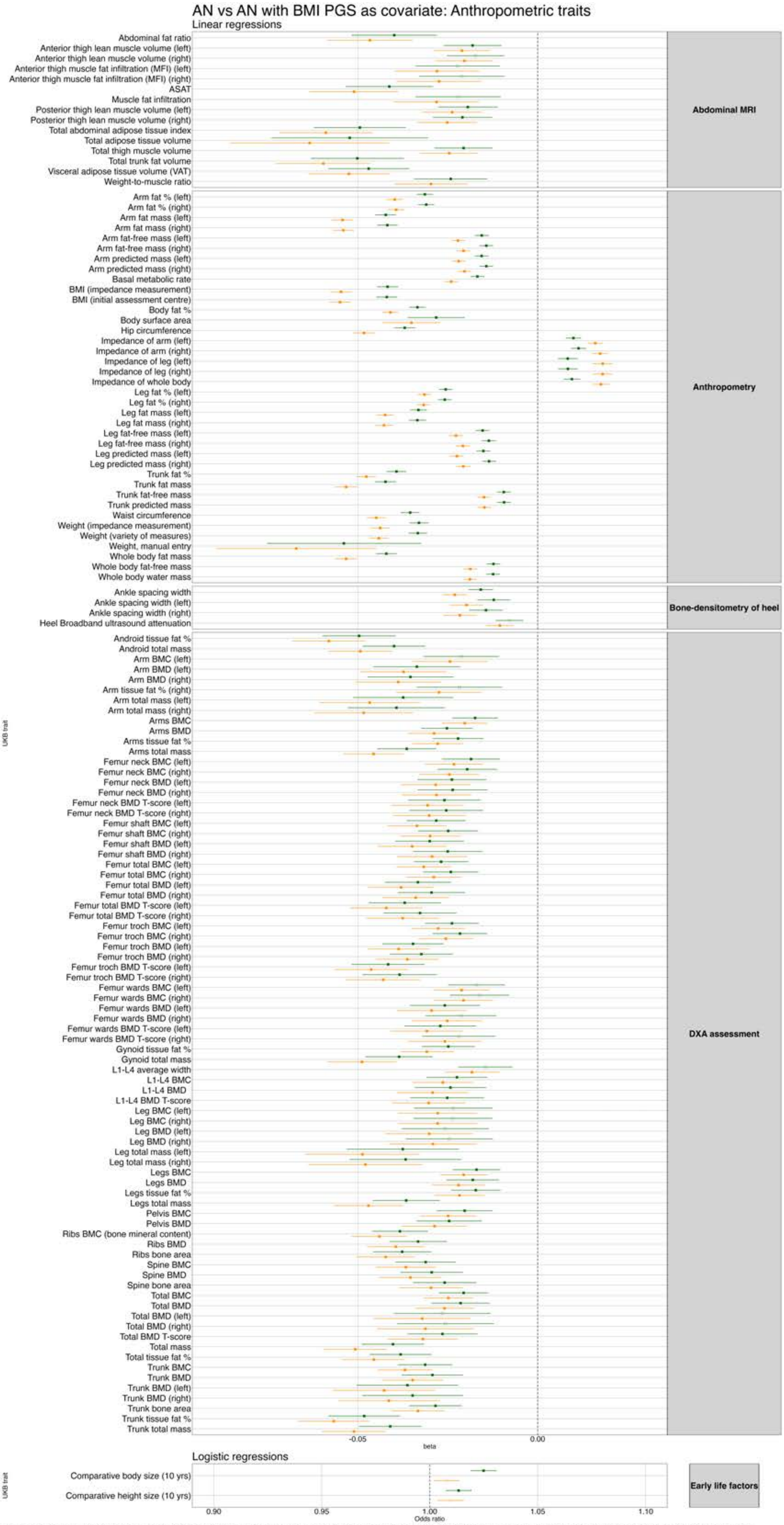


**Figure S19.** Phenome-wide association results between the anorexia nervosa PGS and UK Biobank anthropometric traits from the main analysis (orange) and after controlling for BMI PGS (green). *Note. Filled squares=significant; Empty squares=non-significant. We have displayed all results that were significant in at least one of the pheWASs.*

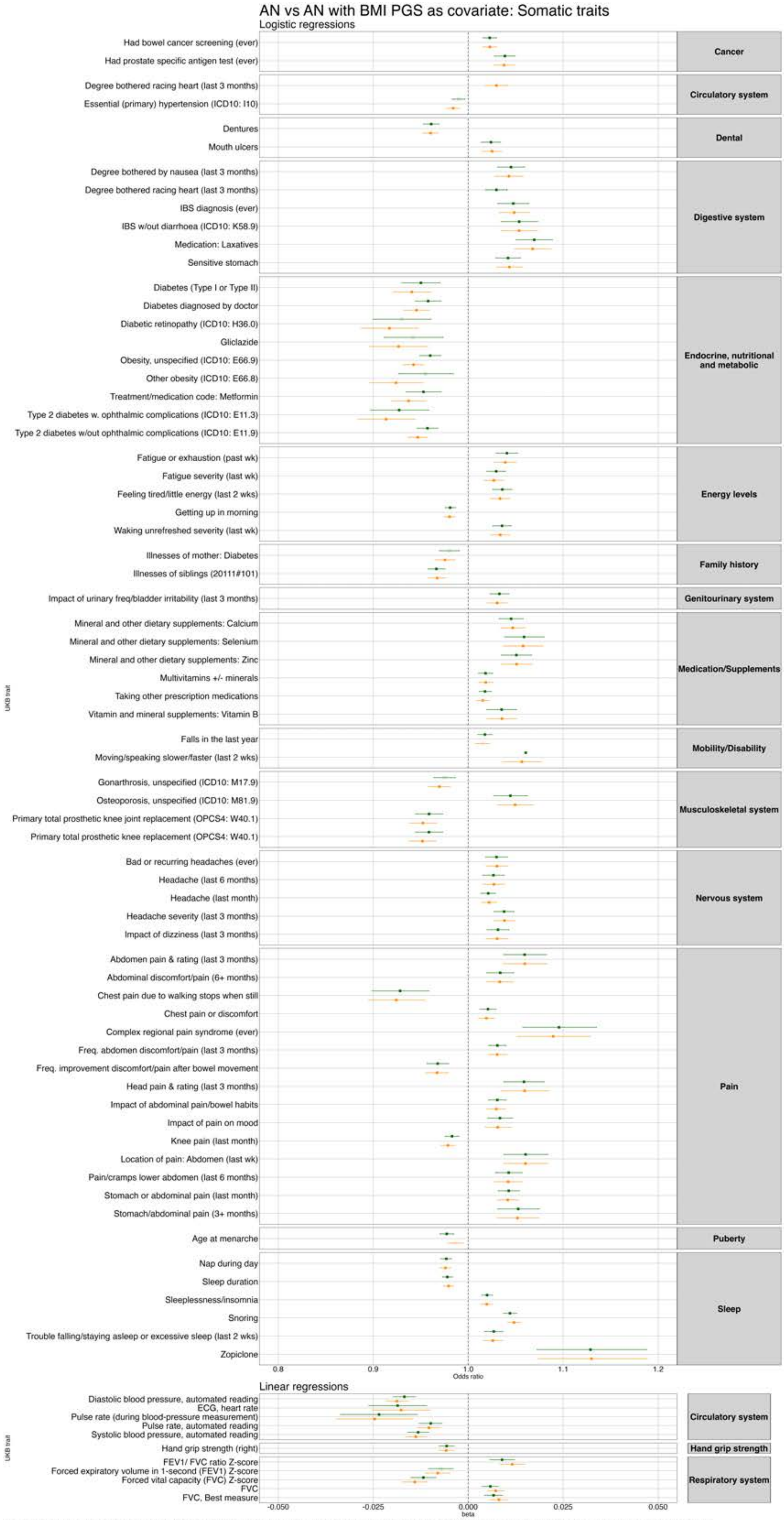


**Figure S20.** Phenome-wide association results between the anorexia nervosa PGS and UK Biobank somatic traits from the main analysis (orange) and after controlling for BMI PGS (green). *Note. Filled squares=significant; Empty squares=non-significant. We have displayed all results that were significant in at least one of the pheWASs.*

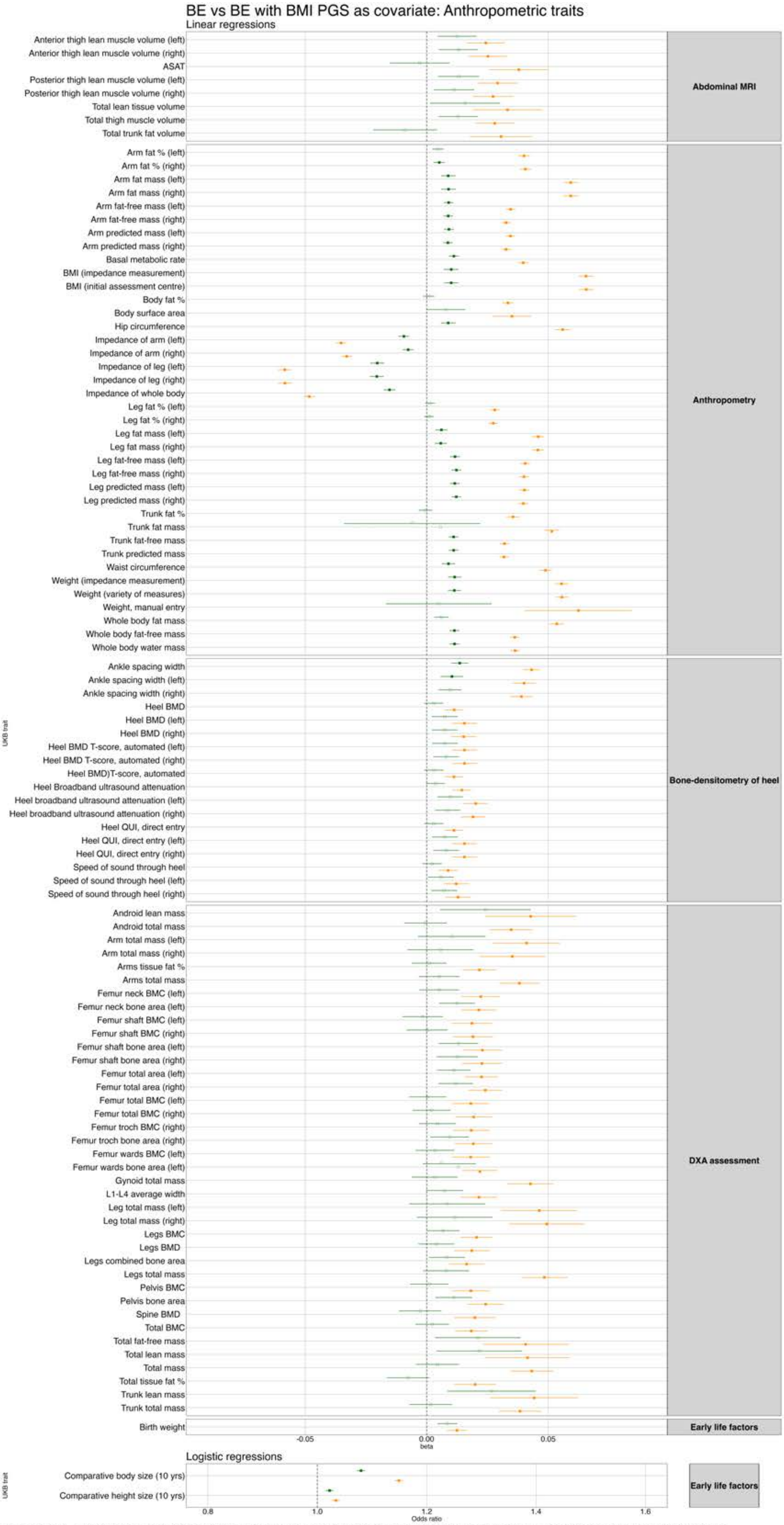


**Figure S21.** Phenome-wide association results between the binge eating PGS and UK Biobank anthropometric traits from the main analysis (orange) and after controlling for BMI PGS (green). *Note. Filled squares=significant; Empty squares=non-significant. We have displayed all results that were significant in at least one of the pheWASs.*

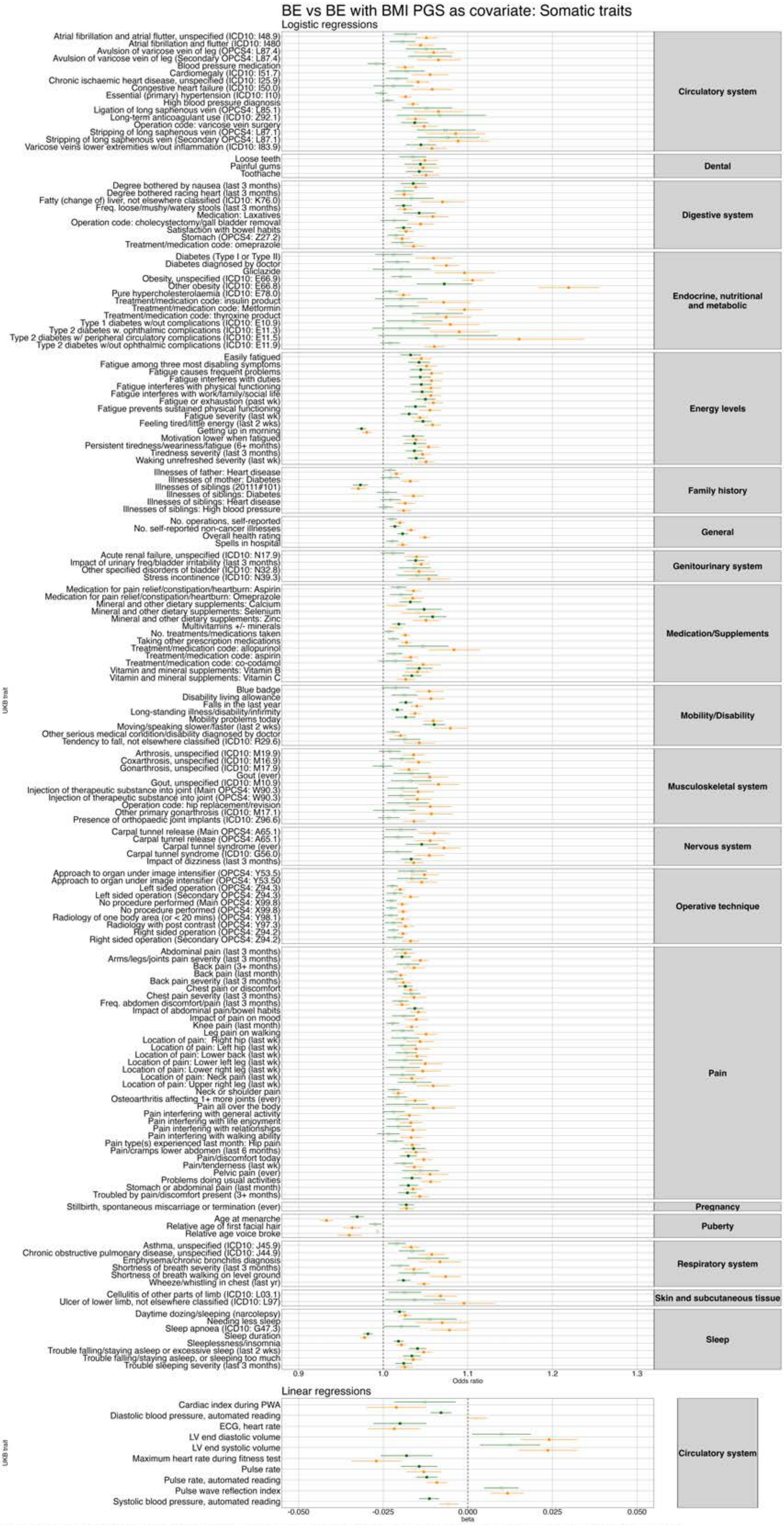


**Figure S22.** Phenome-wide association results between the binge eating PGS and UK Biobank somatic traits from the main analysis (orange) and after controlling for BMI PGS (green). *Note. Filled squares=significant; Empty squares=non-significant. We have displayed all results that were significant in at least one of the pheWASs.*

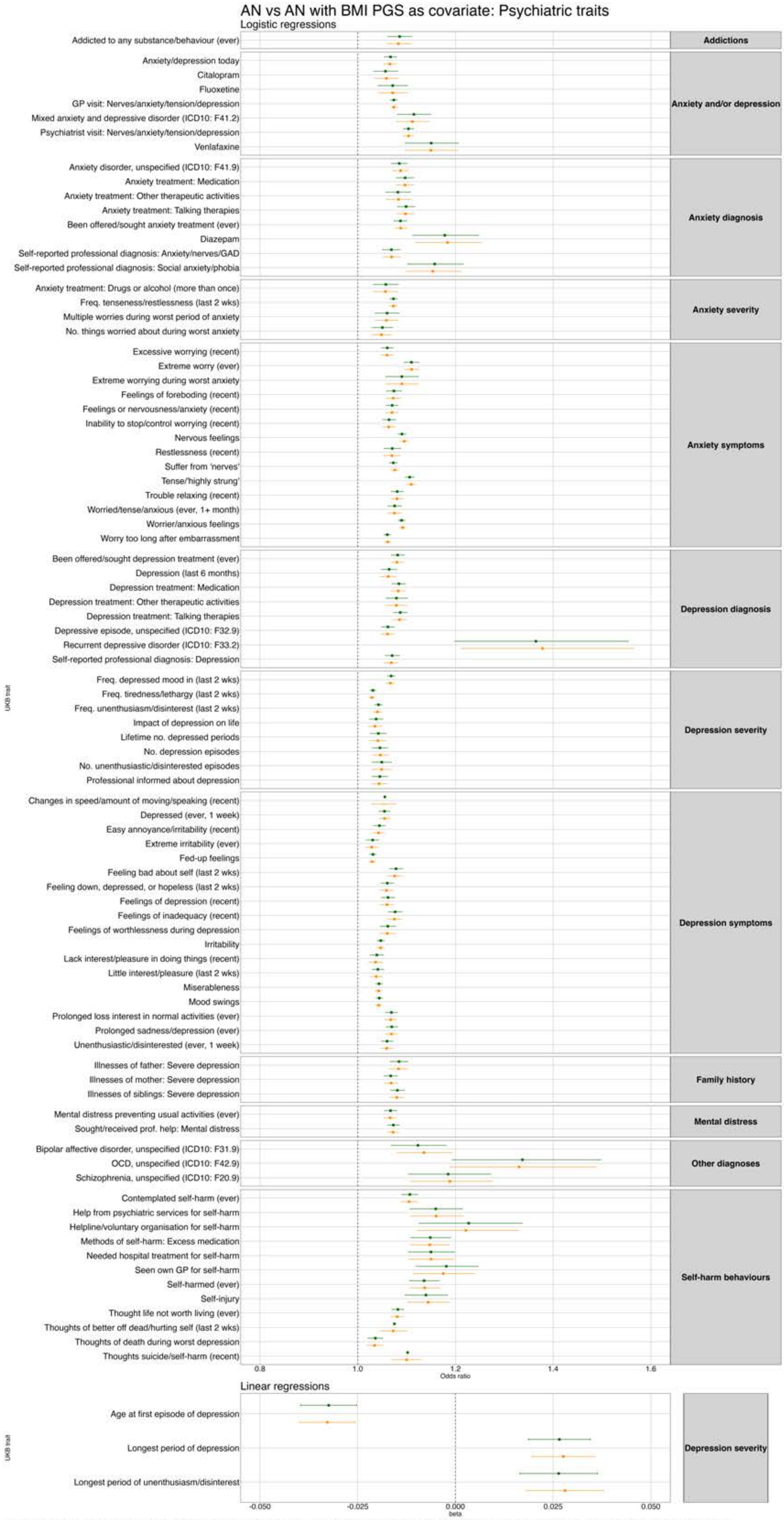

**Figure S23.** Phenome-wide association results between the anorexia nervosa PGS and UK Biobank psychiatric traits from the main analysis (orange) and after controlling for BMI PGS (green). *Note. Filled squares=significant; Empty squares=non-significant. We have displayed all results that were significant in at least one of the pheWASs.*

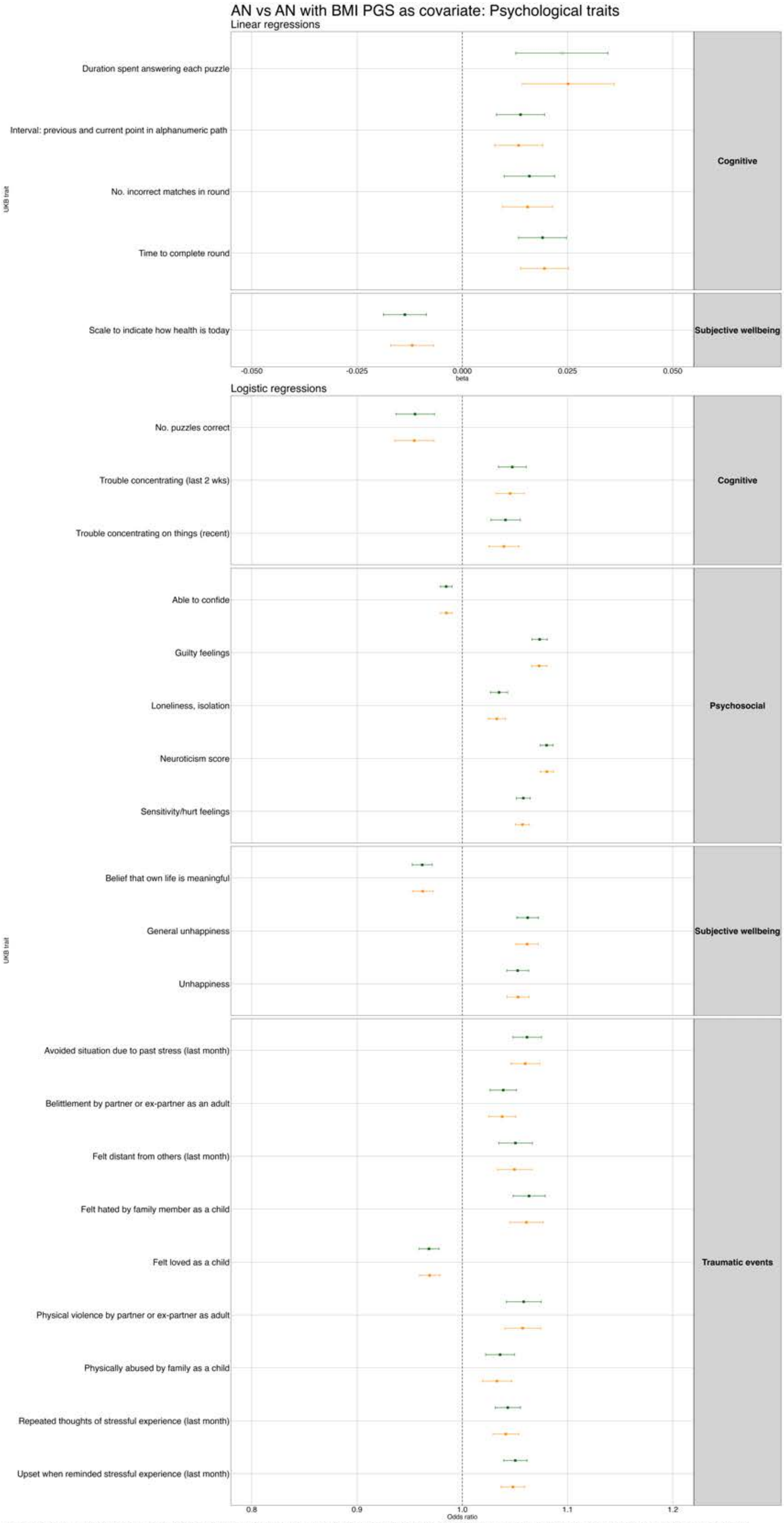


**Figure S24.** Phenome-wide association results between the anorexia nervosa PGS and UK Biobank psychological traits from the main analysis (orange) and after controlling for BMI PGS (green). *Note. Filled squares=significant; Empty squares=non-significant. We have displayed all results that were significant in at least one of the pheWASs.*

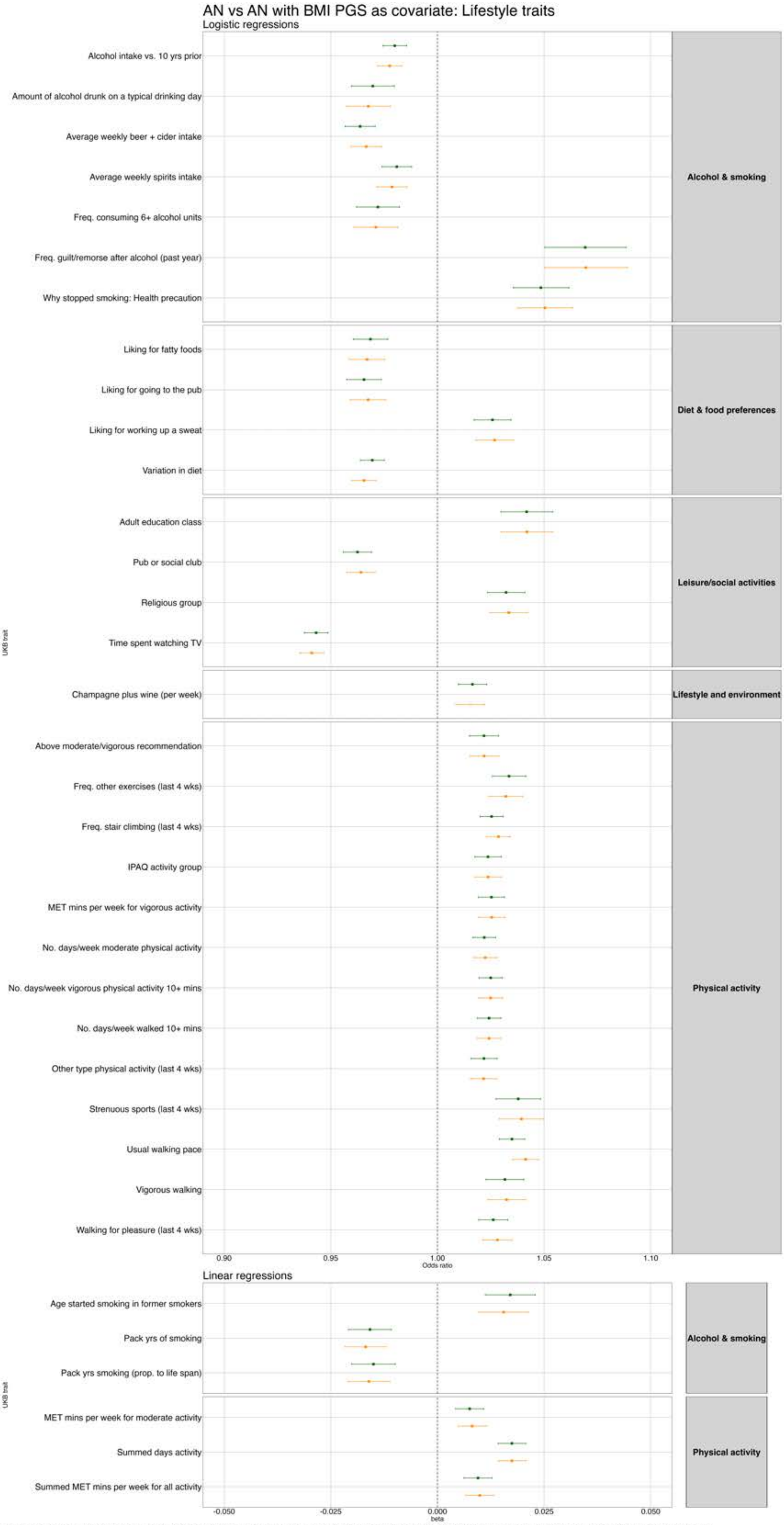


**Figure S25.** Phenome-wide association results between the anorexia nervosa PGS and UK Biobank lifestyle traits from the main analysis (orange) and after controlling for BMI PGS (green). *Note. Filled squares=significant; Empty squares=non-significant. We have displayed all results that were significant in at least one of the pheWASs.*

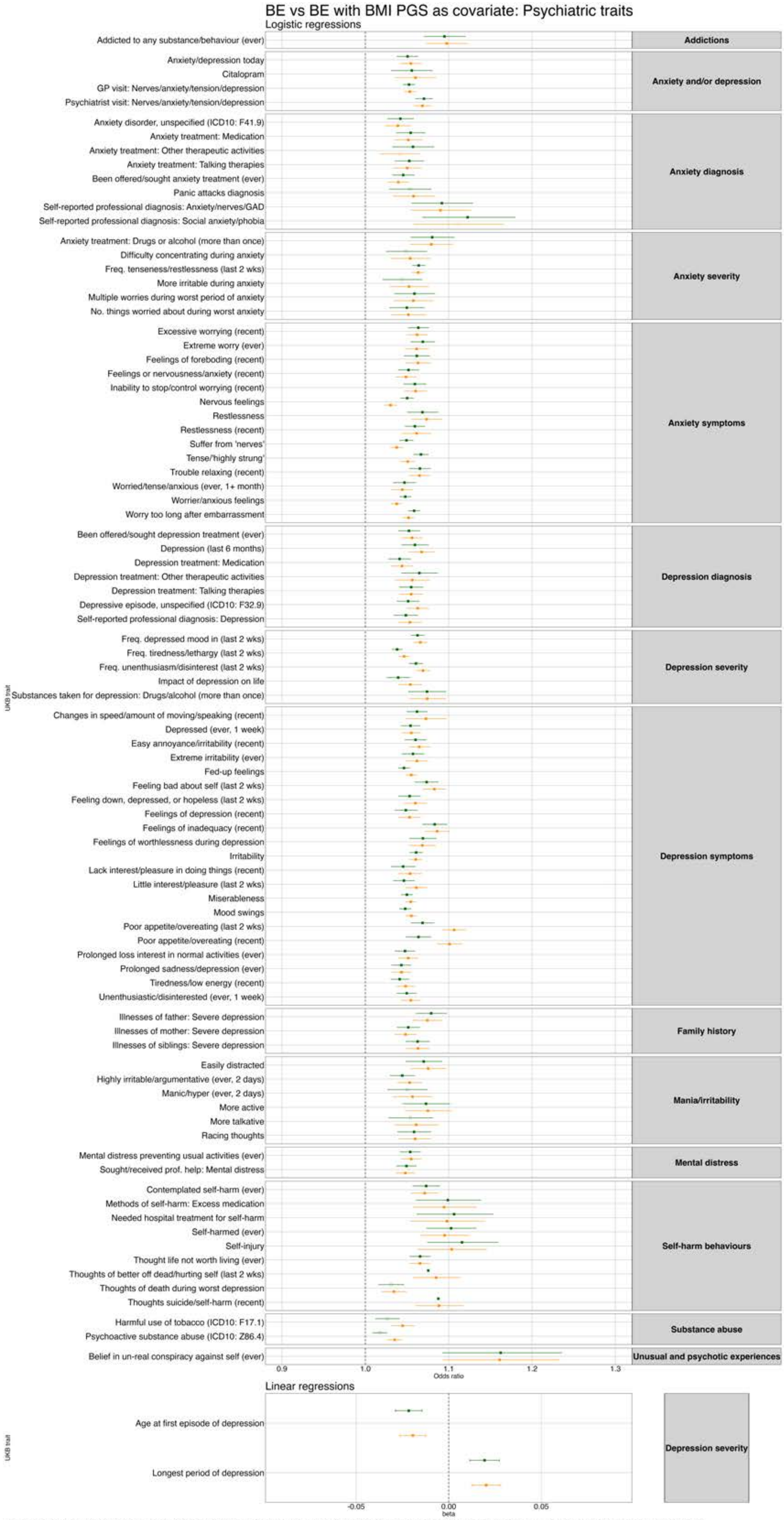


**Figure S26.** Phenome-wide association results between the binge eating PGS and UK Biobank psychiatric traits from the main analysis (orange) and after controlling for BMI PGS (green). *Note. Filled squares=significant; Empty squares=non-significant. We have displayed all results that were significant in at least one of the pheWASs.*

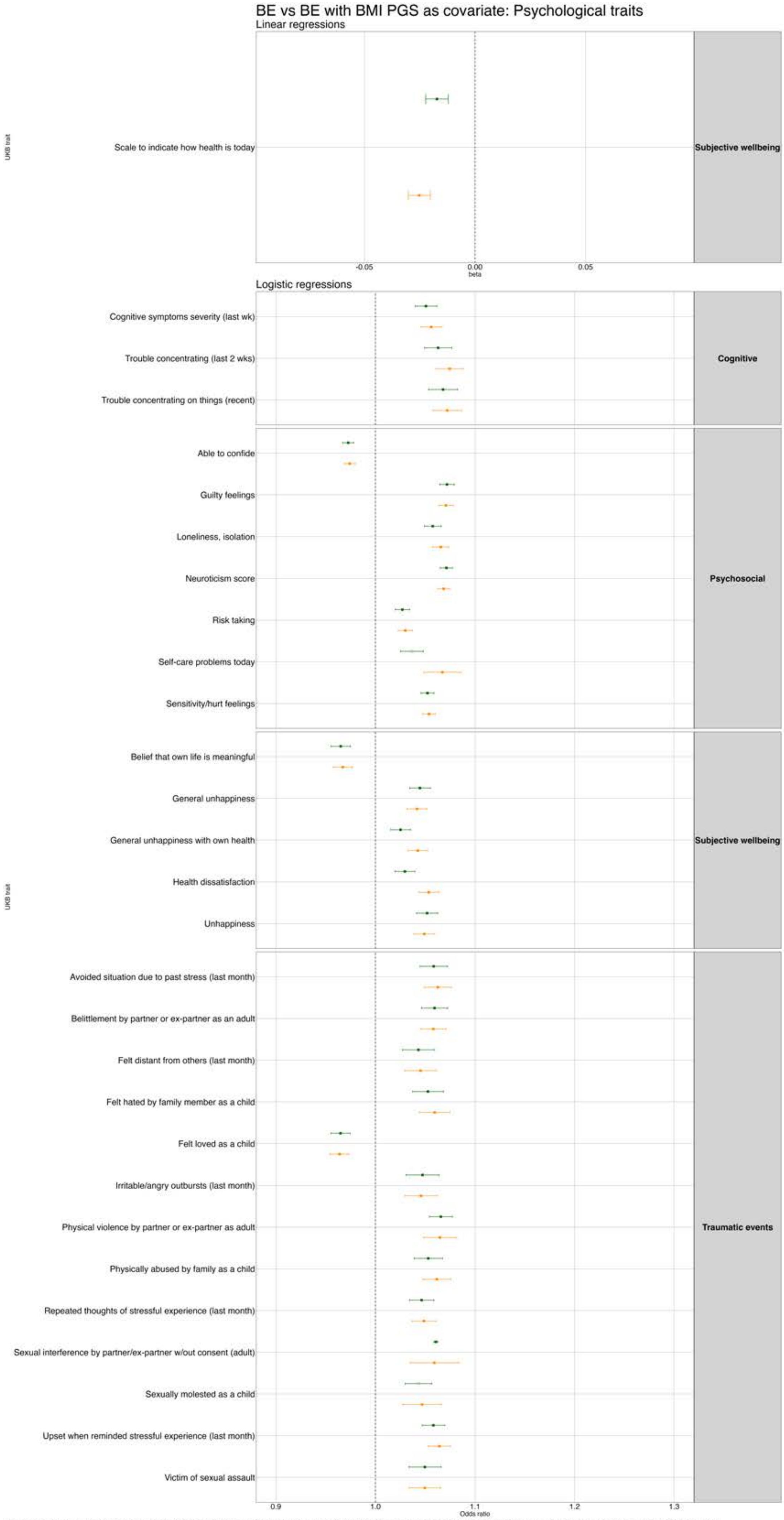


**Figure S27.** Phenome-wide association results between the binge eating PGS and UK Biobank psychological traits from the main analysis (orange) and after controlling for BMI PGS (green). *Note. Filled squares=significant; Empty squares=non-significant. We have displayed all results that were significant in at least one of the pheWASs.*

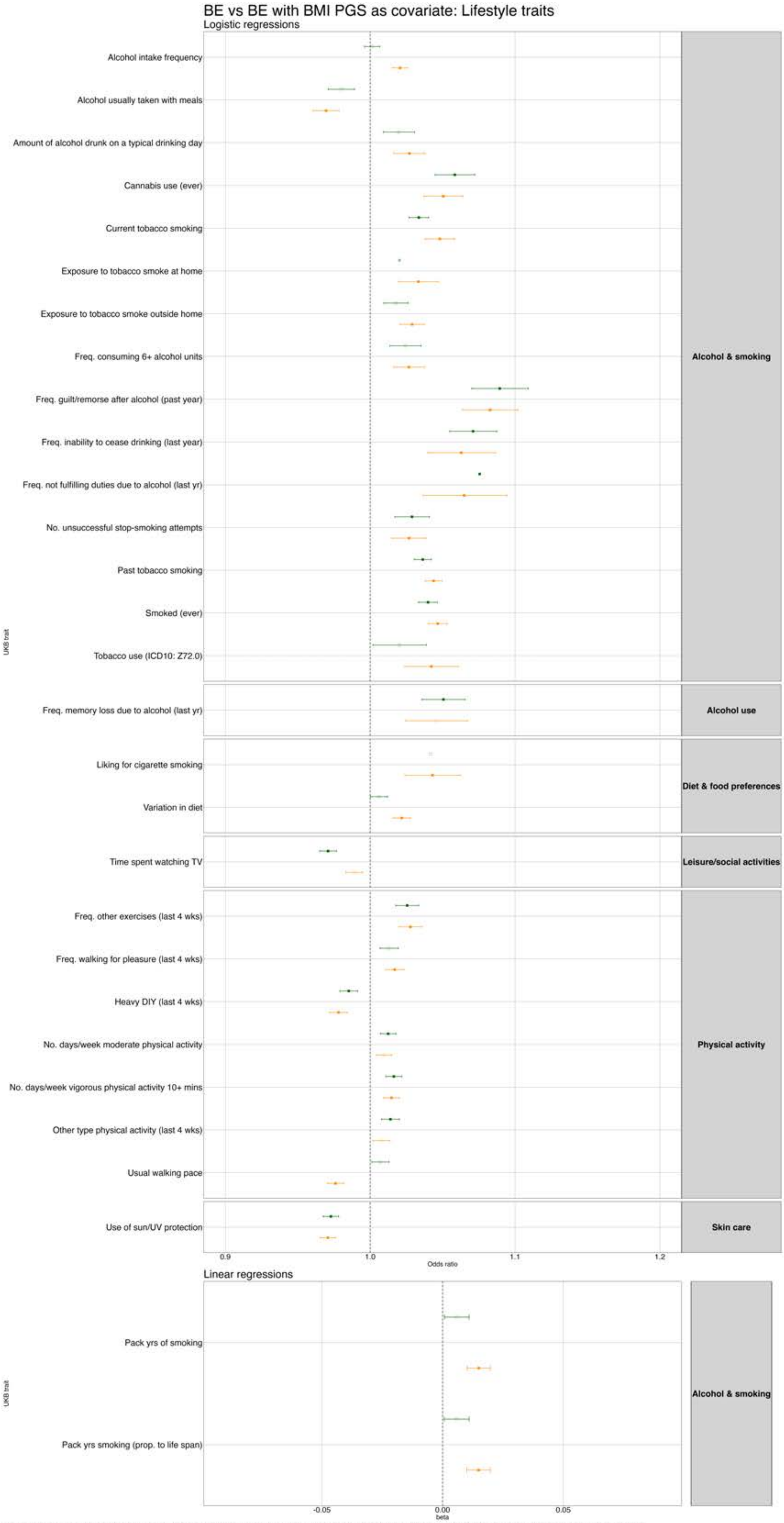


**Figure S28.** Phenome-wide association results between the lifestyle PGS and UK Biobank biomarkers from the main analysis (orange) and after controlling for BMI PGS (green). *Note. Filled squares=significant; Empty squares=non-significant. We have displayed all results that were significant in at least one of the pheWASs.*

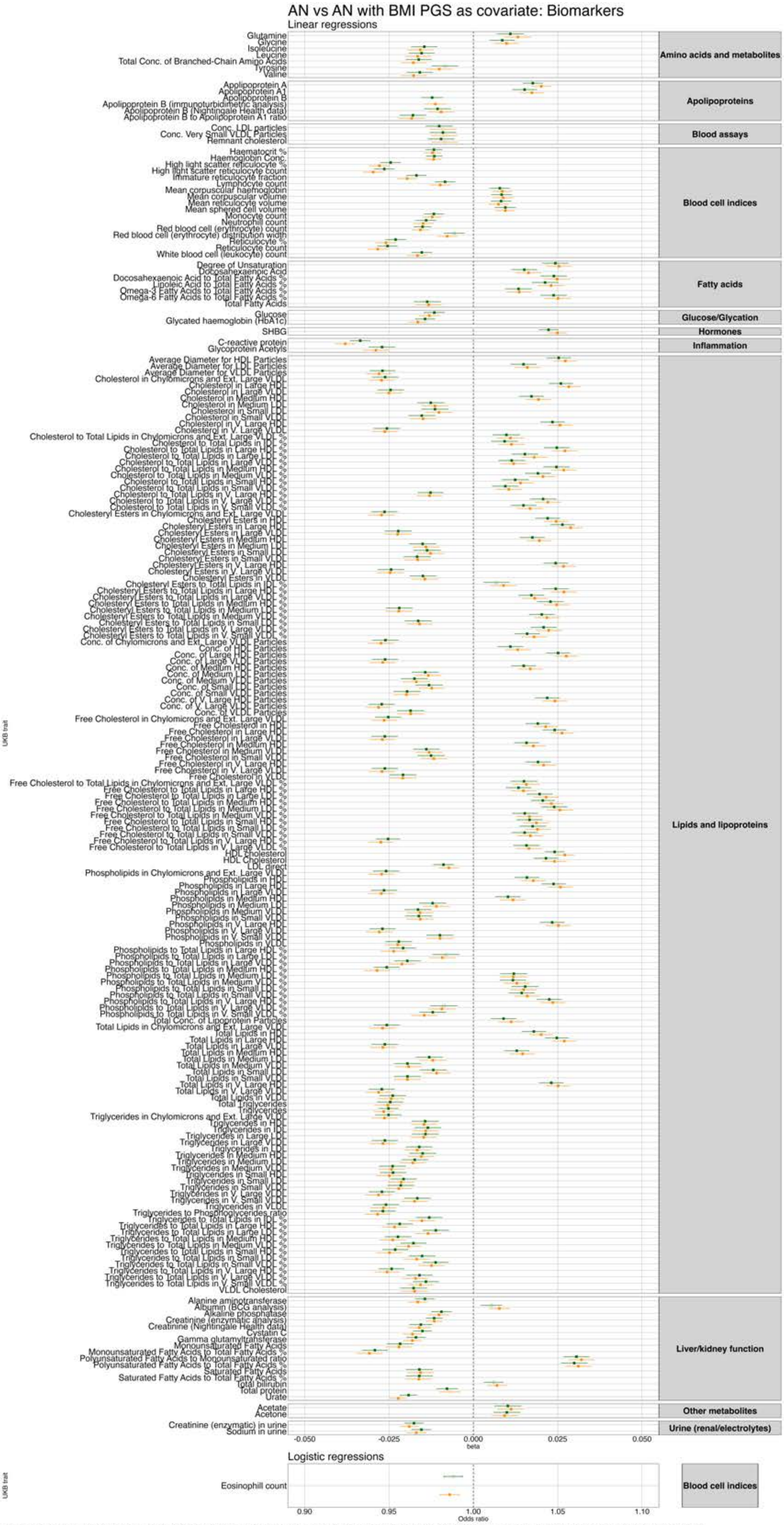


**Figure S29.** Phenome-wide association results between the anorexia nervosa PGS and UK Biobank biomarkers from the main analysis (orange) and after controlling for BMI PGS (green). *Note. Filled squares=significant; Empty squares=non-significant. We have displayed all results that were significant in at least one of the pheWASs.*

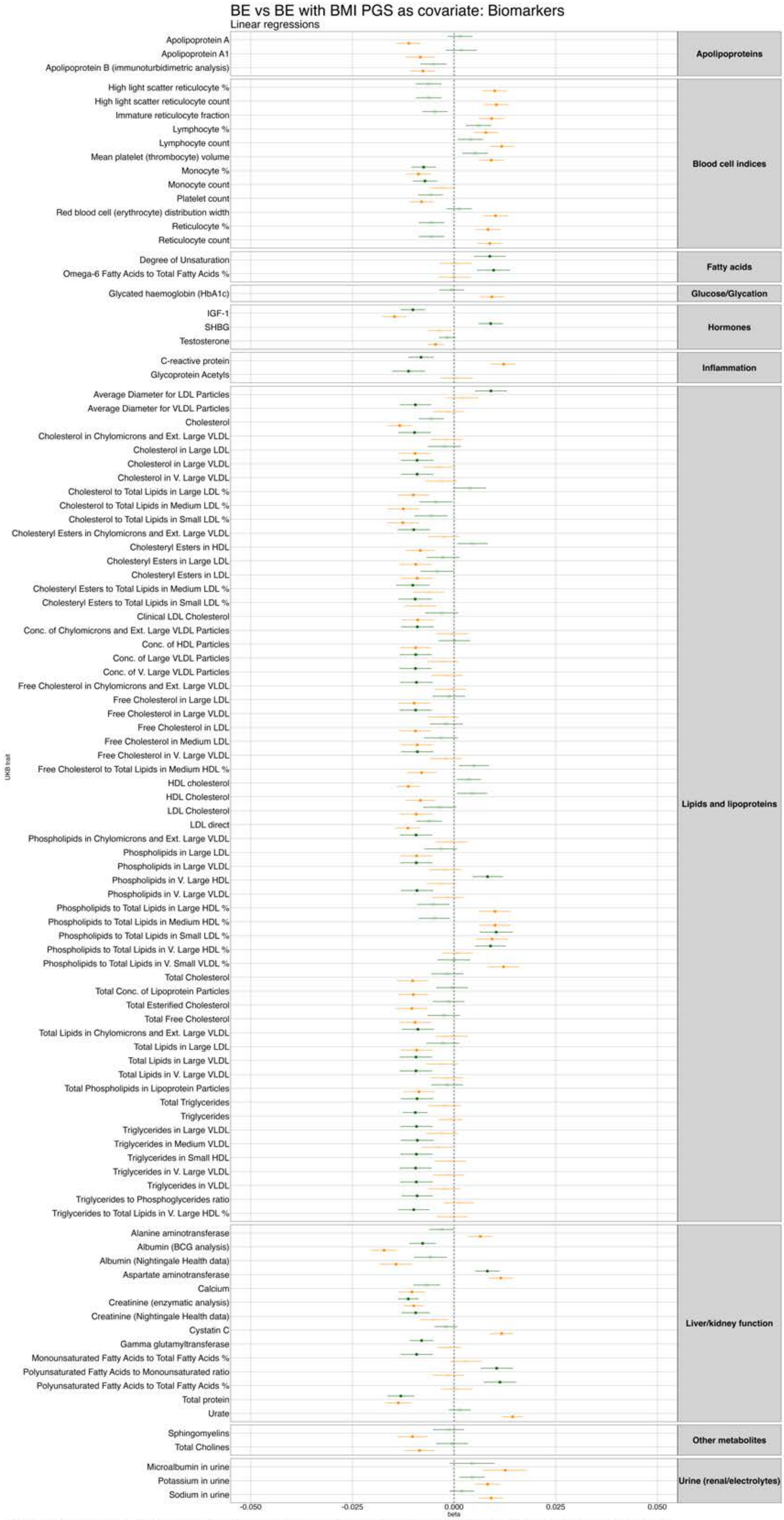


**Figure S30.** Phenome-wide association results between the binge eating PGS and UK Biobank biomarkers from the main analysis (orange) and after controlling for BMI PGS (green). *Note. Filled squares=significant; Empty squares=non-significant. We have displayed all results that were significant in at least one of the pheWASs.*

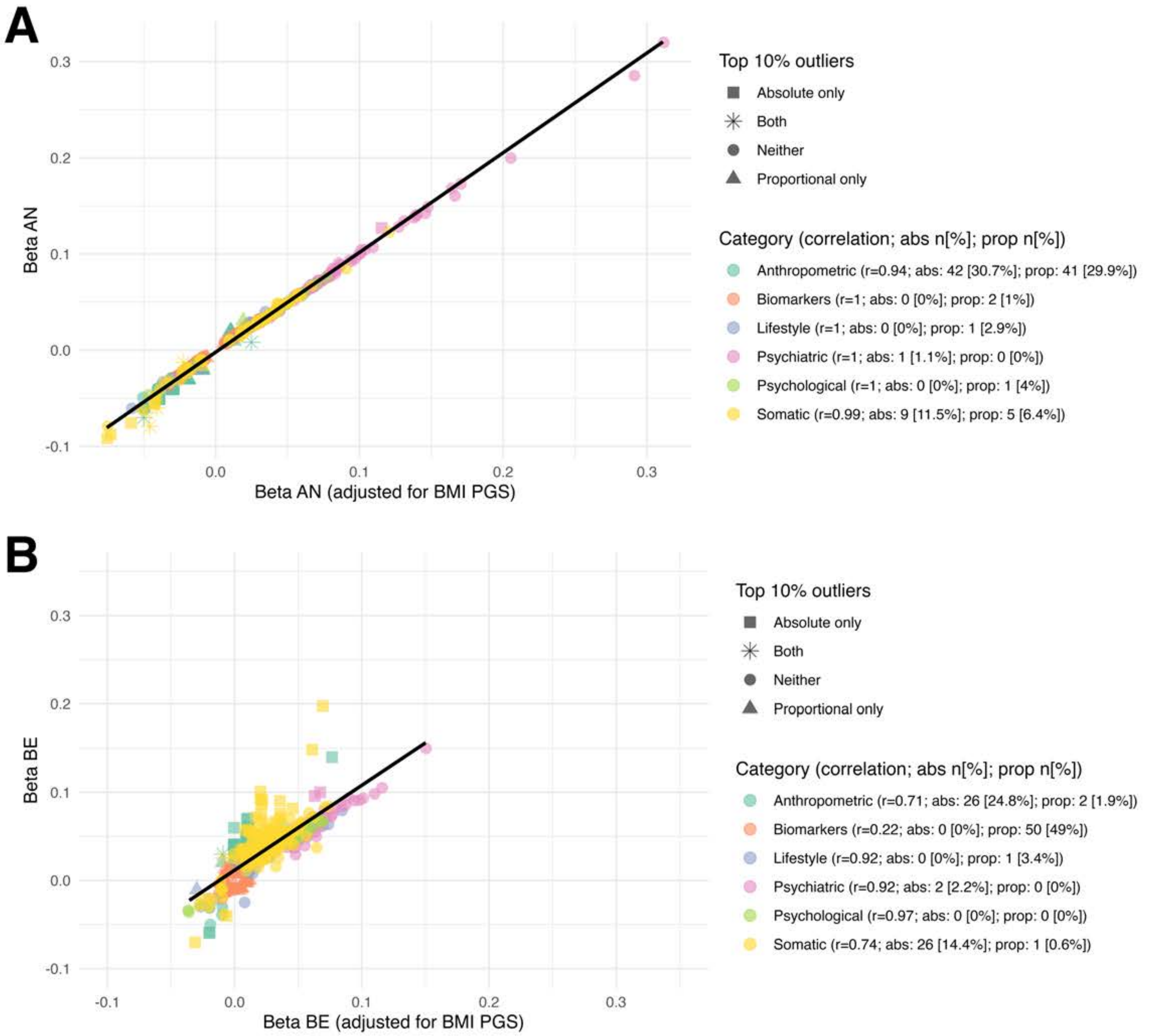


**Figure S31.** Correlation between original analyses results and analyses adjusted for body mass index PGS in a phenome-wide association study in which the exposure is A) anorexia nervosa polygenic score and B) binge eating polygenic score, and the outcomes are UK Biobank health-related traits.
Number and percentage of traits within each category that are in the absolute and proportional top 10% of change following adjustment are also displayed.

*Note. Odds ratios are converted to logarithmic odds for consistency with beta. The low correlation between BMI-adjusted and unadjusted betas for biomarkers in B but zero values falling in the top 10% of absolute change is due to very small effect sizes in this category (many close to zero).*

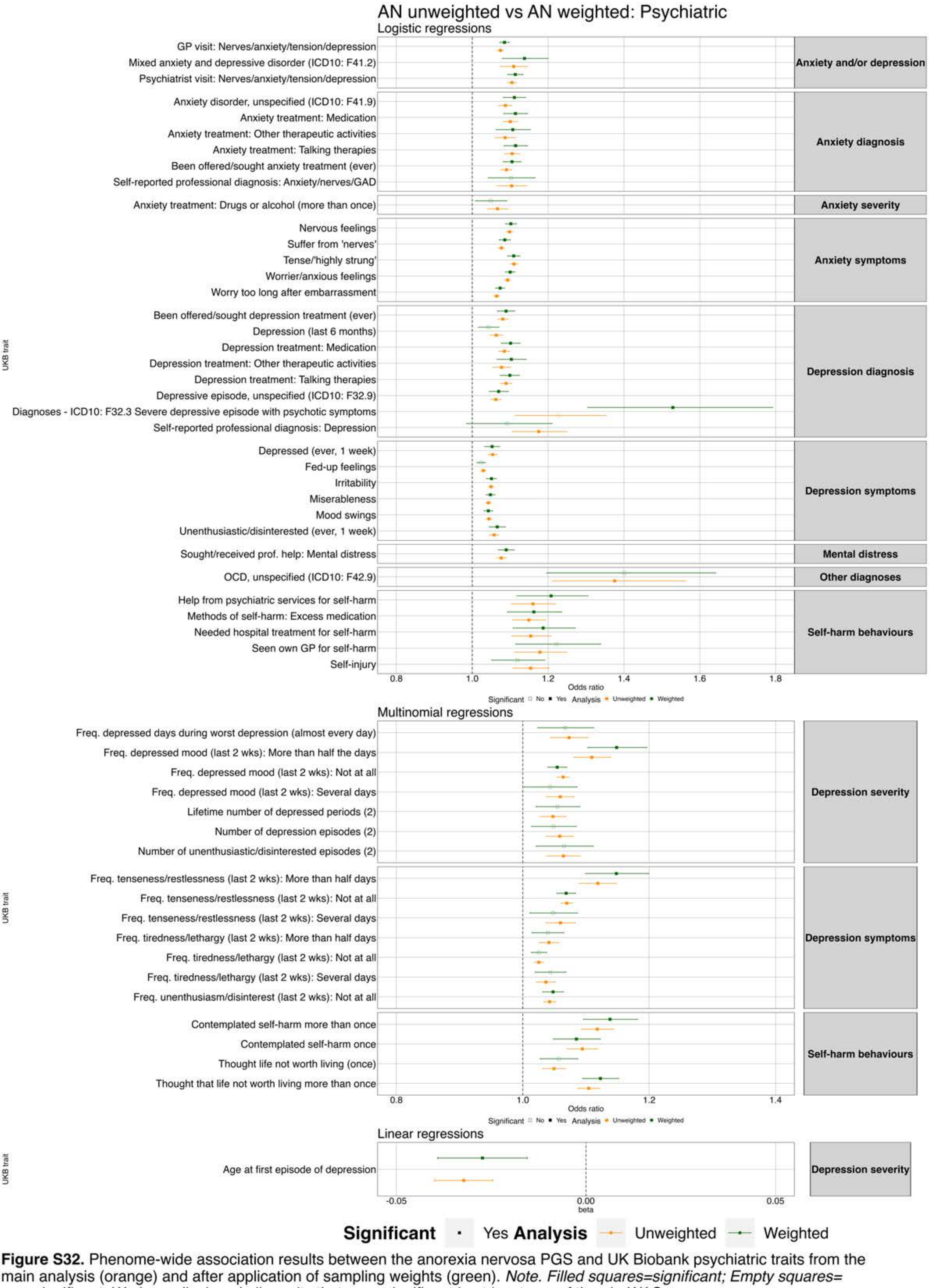


**Figure S32.** Phenome-wide association results between the anorexia nervosa PGS and UK Biobank psychiatric traits from the main analysis (orange) and after application of sampling weights (green). *Note. Filled squares=significant; Empty squares= non-significant. We have displayed all results that were significant in at least one of the pheWASs.*

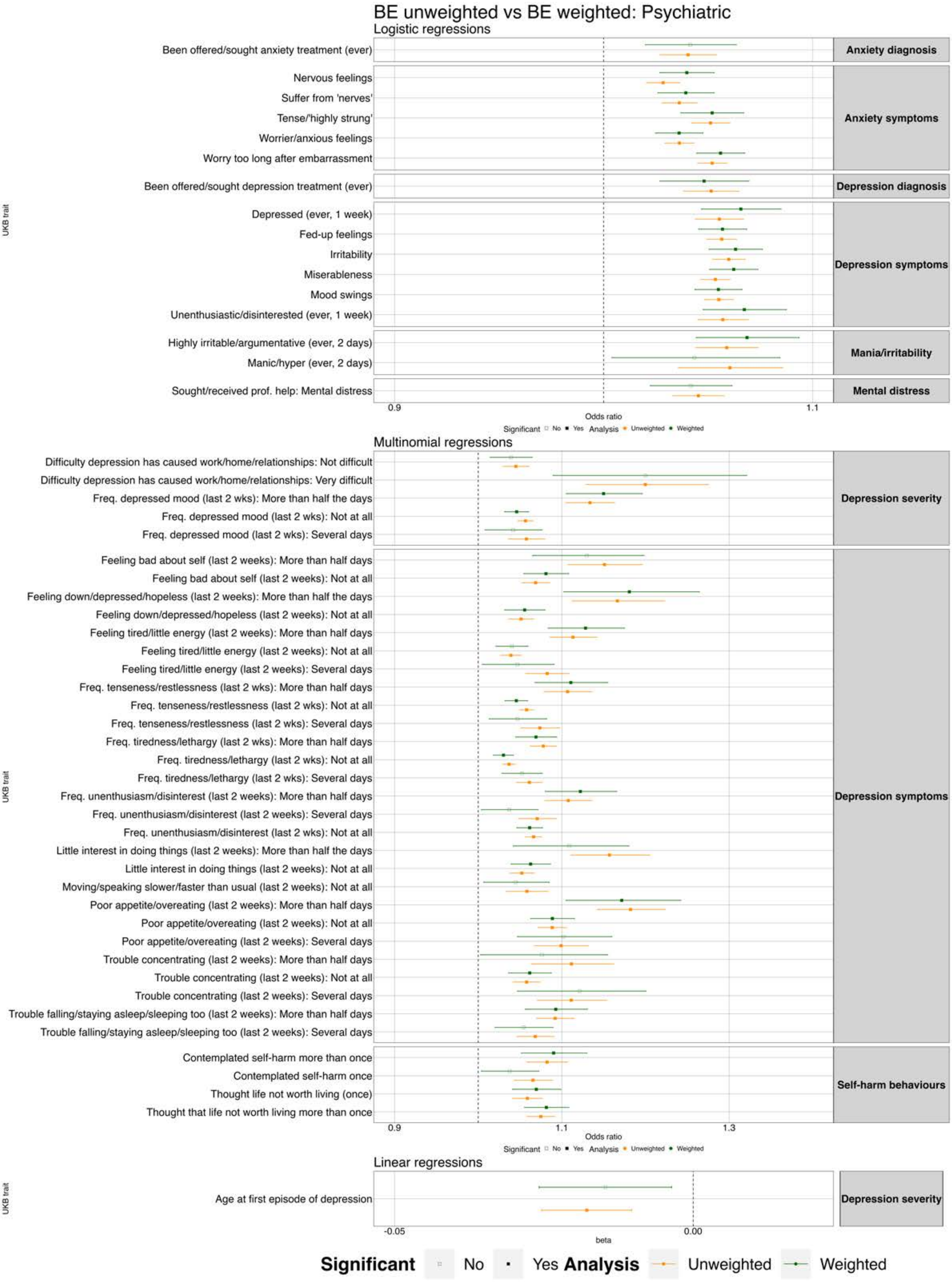


**Figure S33.** Phenome-wide association results between the binge eating PGS and UK Biobank psychiatric traits from the main analysis (orange) and after application of sampling weights (green). *Note. Filled squares=significant; Empty squares= non-significant. We have displayed all results that were significant in at least one of the pheWASs.*

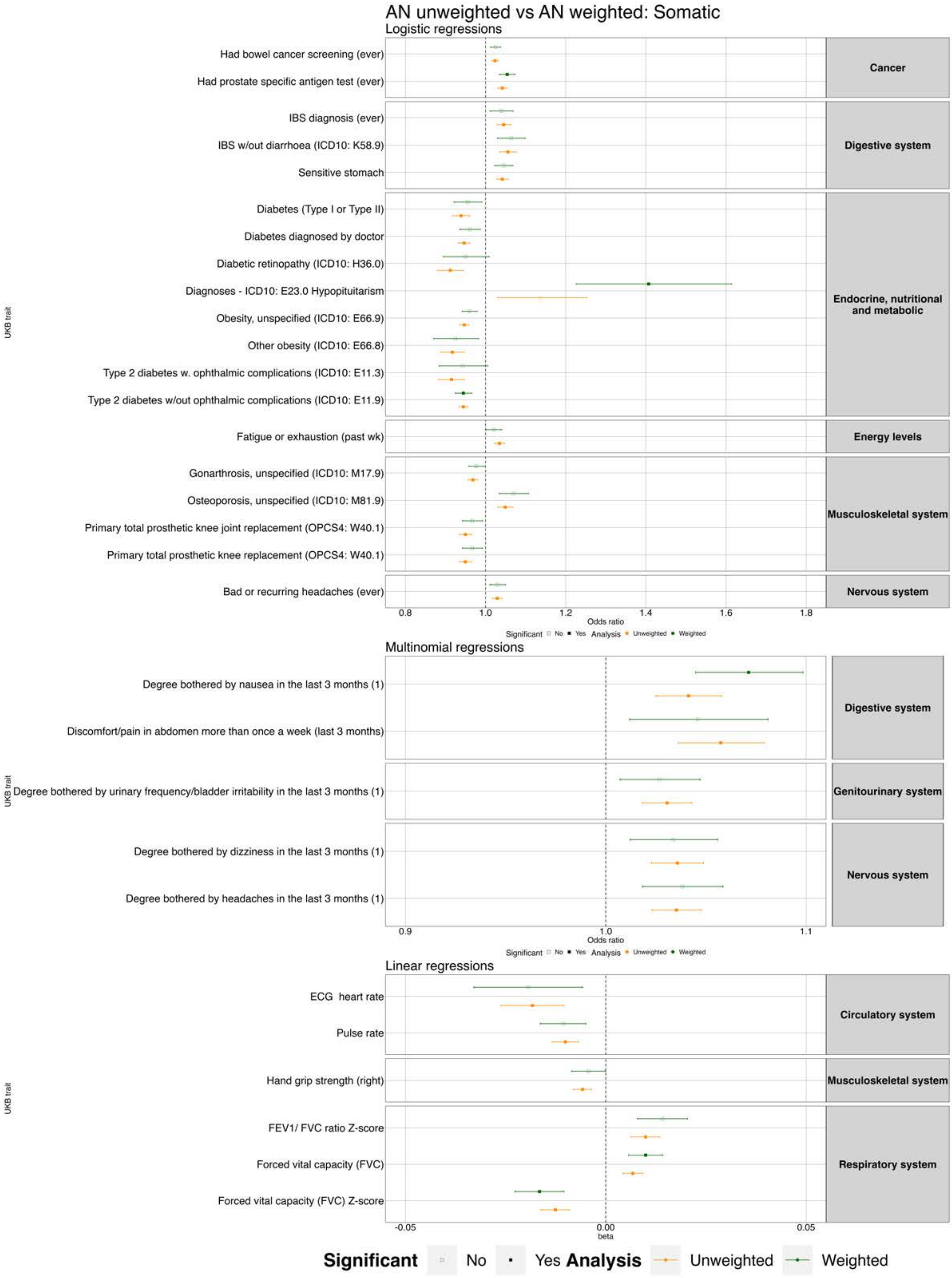


**Figure S34.** Phenome-wide association results between the anorexia nervosa PGS and UK Biobank somatic traits from the main analysis (orange) and after application of sampling weights (green). *Note. Filled squares=significant; Empty squares= non-significant. We have displayed all results that were significant in at least one of the pheWASs.*

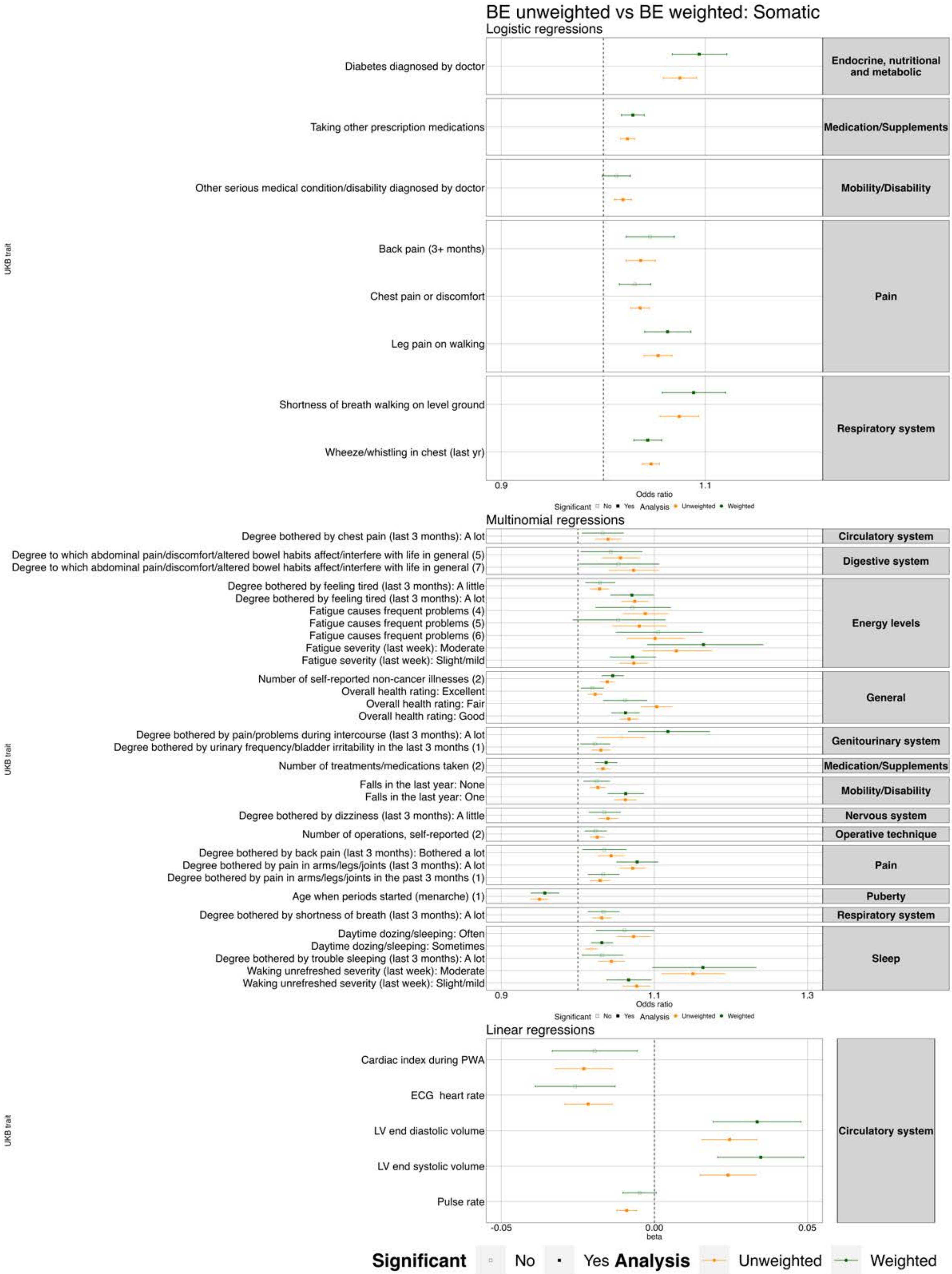


**Figure S35.** Phenome-wide association results between the binge eating PGS and UK Biobank somatic traits from the main analysis (orange) and after application of sampling weights (green). *Note. Filled squares=significant; Empty squares= non-significant. We have displayed all results that were significant in at least one of the pheWASs.*

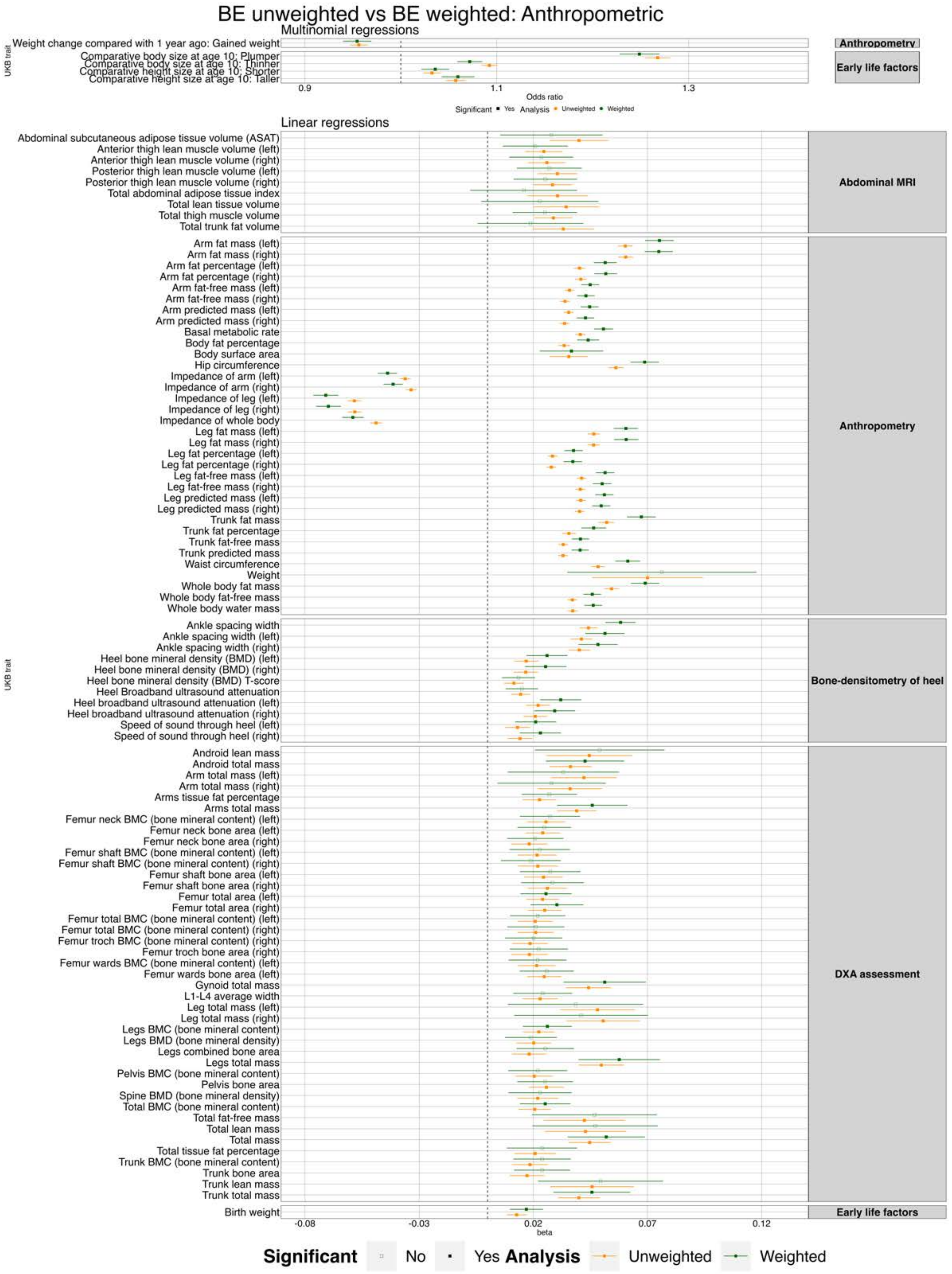


**Figure S36.** Phenome-wide association results between the binge eating PGS and UK Biobank anthropometric traits from the main analysis (orange) and after application of sampling weights (green). *Note. Filled squares=significant; Empty squares= non-significant. We have displayed all results that were significant in at least one of the pheWASs.*

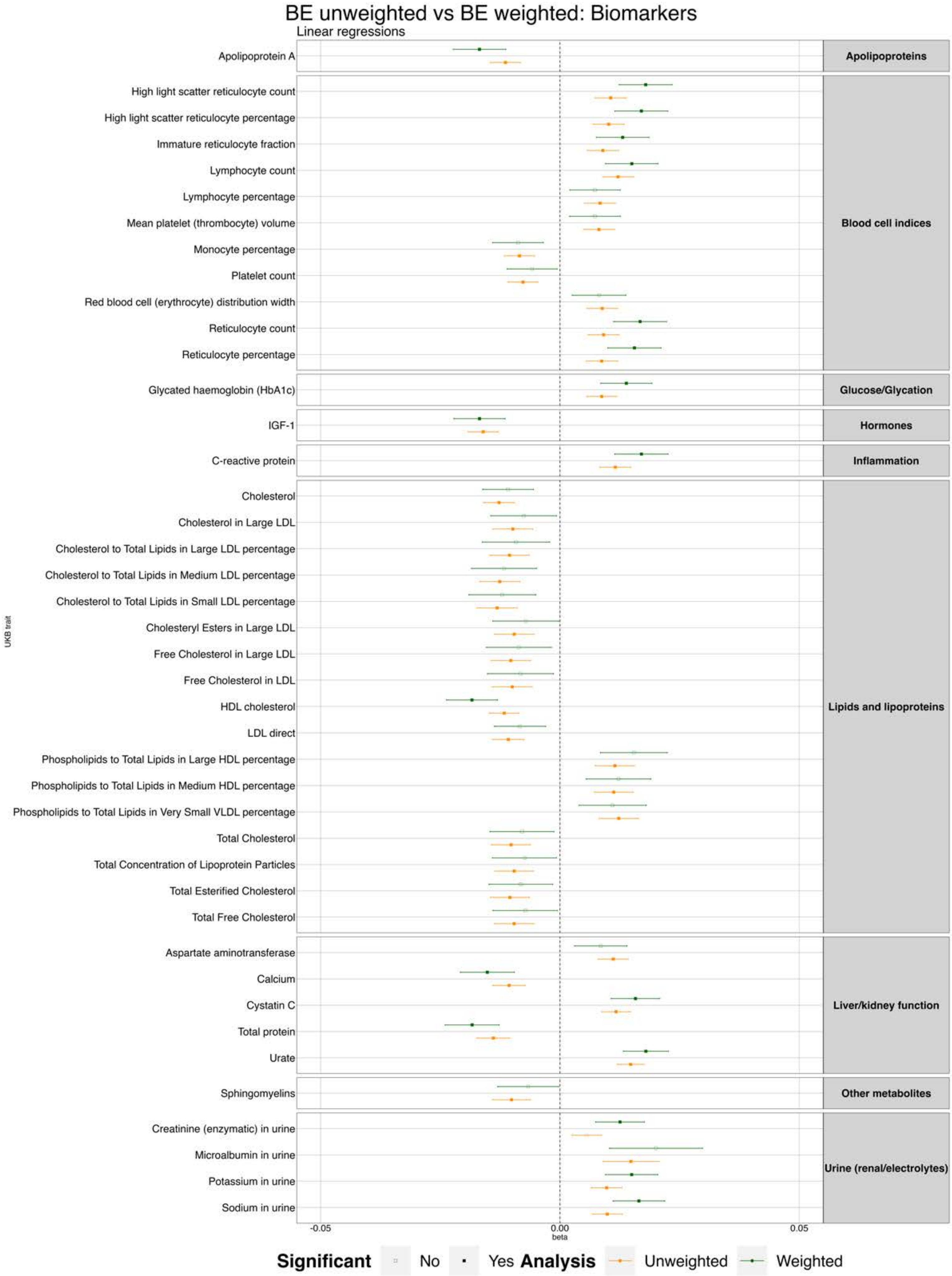


**Figure S37.** Phenome-wide association results between the binge eating PGS and UK Biobank biomarkers from the main analysis (orange) and after application of sampling weights (green). *Note. Filled squares=significant; Empty squares= non-significant. We have displayed all results that were significant in at least one of the pheWASs.*

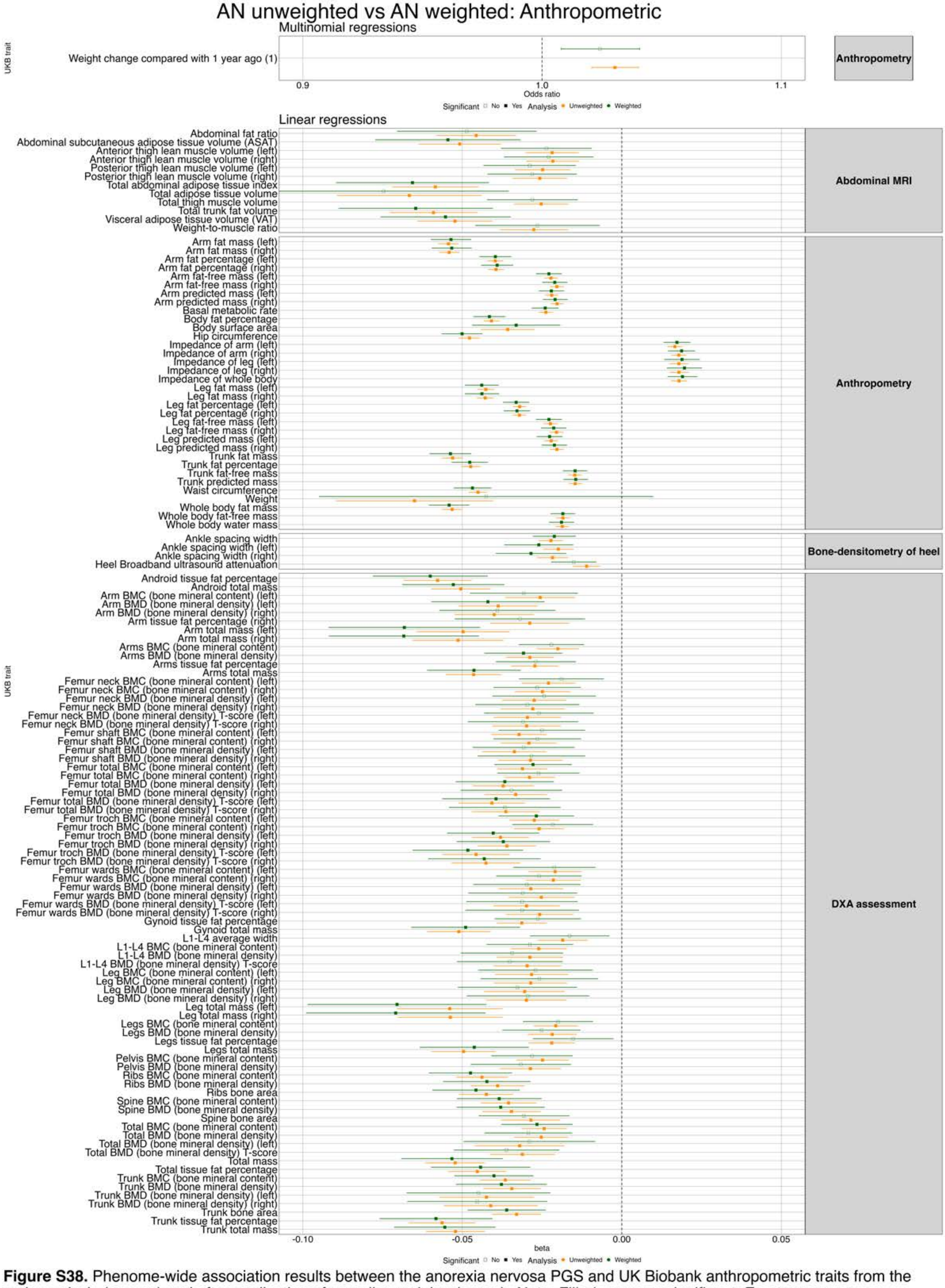

**Figure S38.** Phenome-wide association results between the anorexia nervosa PGS and UK Biobank anthropometric traits from the main analysis (orange) and after application of sampling weights (green). *Note. Filled squares=significant; Empty squares= non-significant. We have displayed all results that were significant in at least one of the pheWASs.*

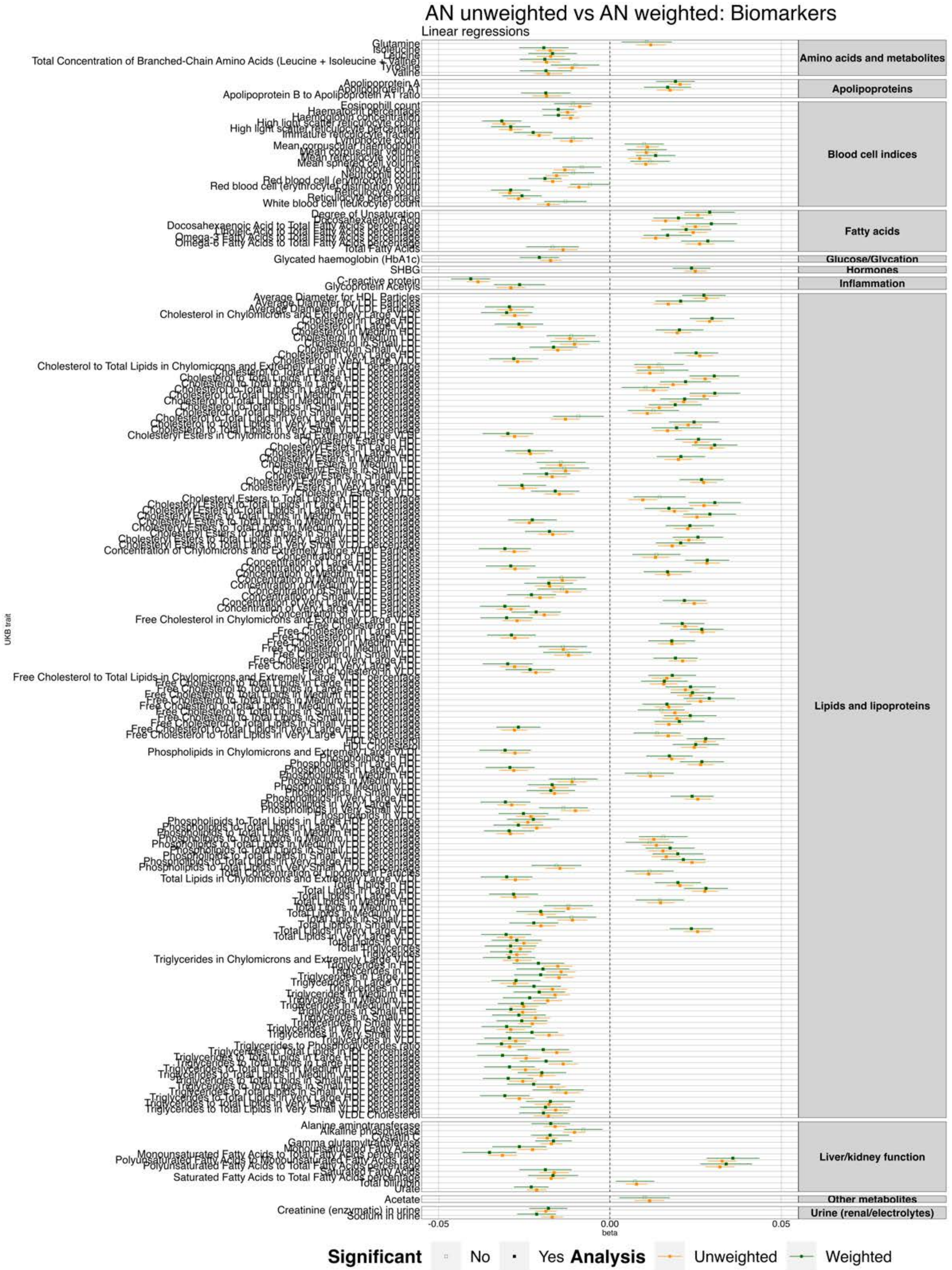


**Figure S39.** Phenome-wide association results between the anorexia nervosa PGS and UK Biobank biomarkers from the main analysis (orange) and after application of sampling weights (green). *Note. Filled squares=significant; Empty squares= non-significant. We have displayed all results that were significant in at least one of the pheWASs.*

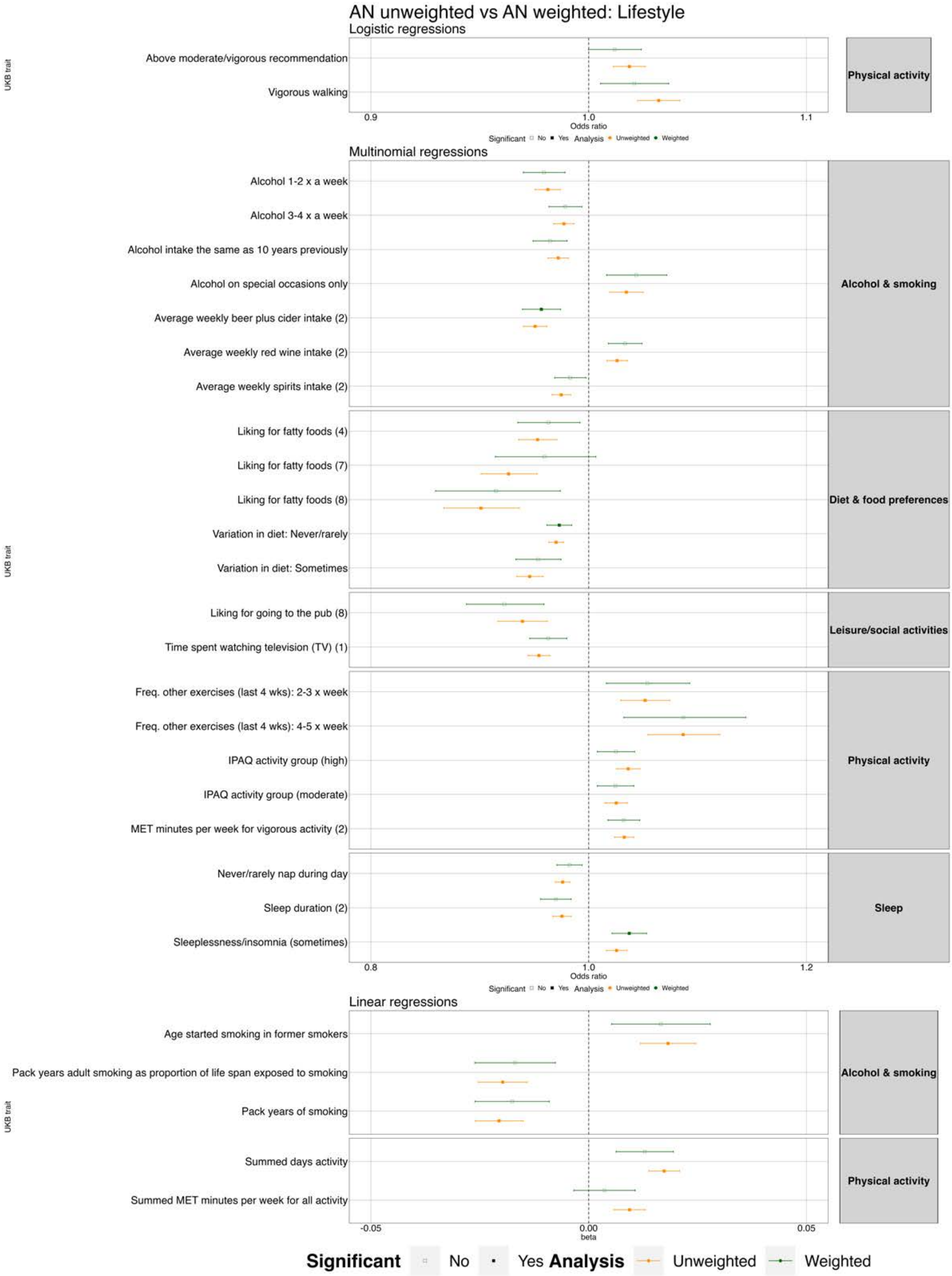


**Figure S40.** Phenome-wide association results between the anorexia nervosa PGS and UK Biobank lifestyle traits from the main analysis (orange) and after application of sampling weights (green). *Note. Filled squares=significant; Empty squares= non-significant. We have displayed all results that were significant in at least one of the pheWASs.*

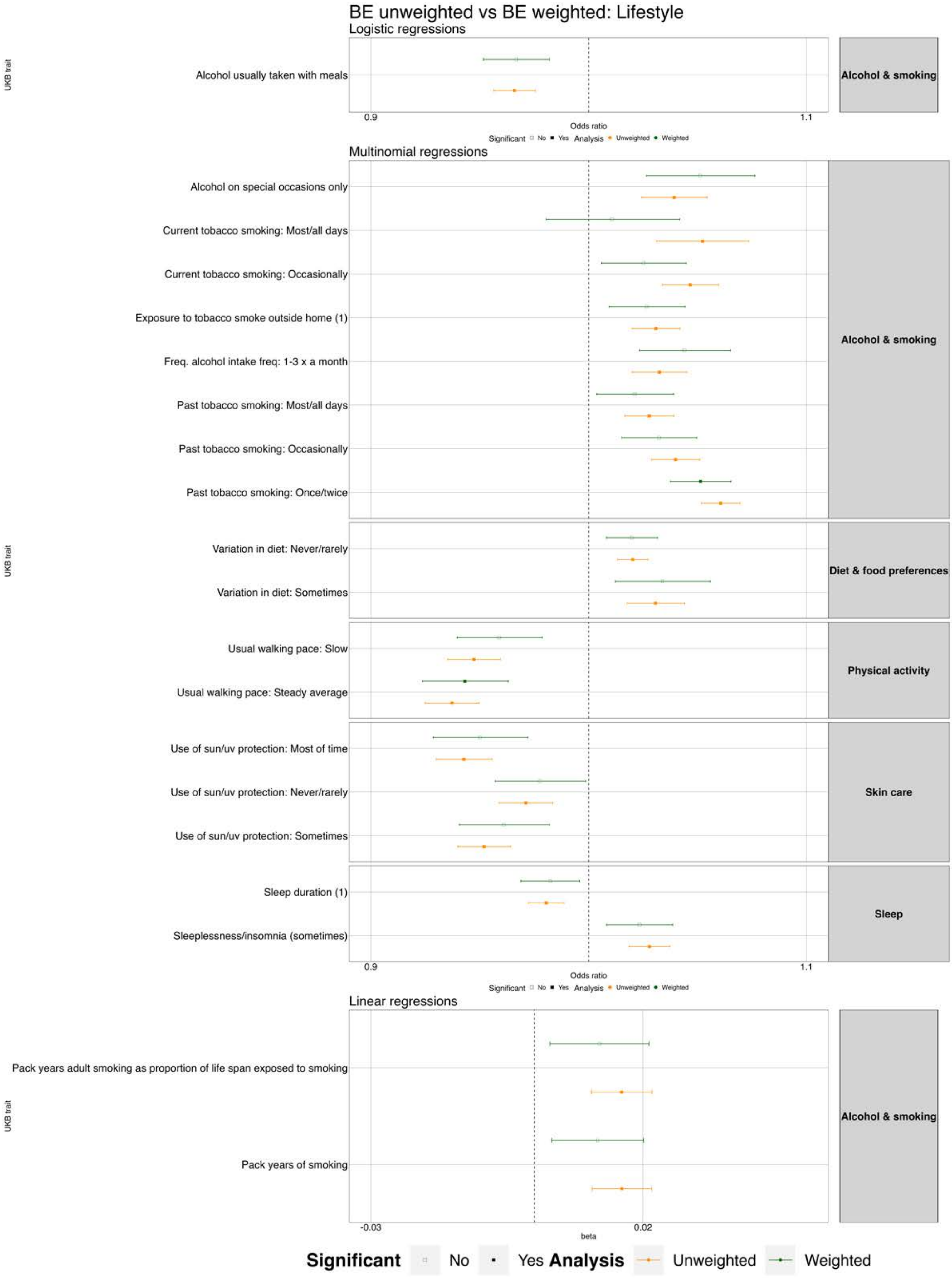


**Figure S41.** Phenome-wide association results between the binge eating PGS and UK Biobank lifestyle traits from the main analysis (orange) and after application of sampling weights (green). *Note. Filled squares=significant; Empty squares= non-significant. We have displayed all results that were significant in at least one of the pheWASs.*

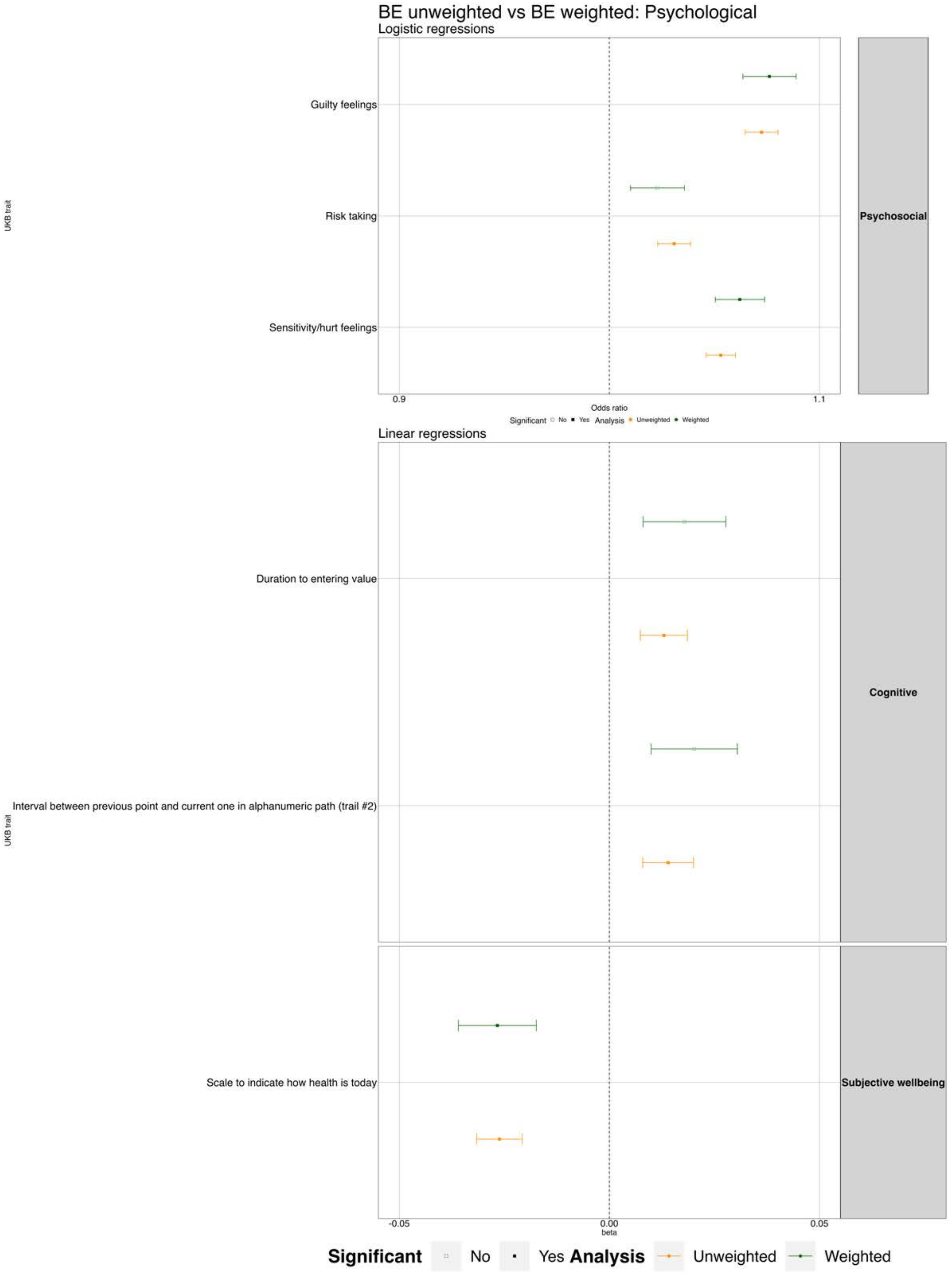


**Figure S42.** Phenome-wide association results between the binge eating PGS and UK Biobank psychological traits from the main analysis (orange) and after application of sampling weights (green). *Note. Filled squares=significant; Empty squares= non-significant. We have displayed all results that were significant in at least one of the pheWASs.*

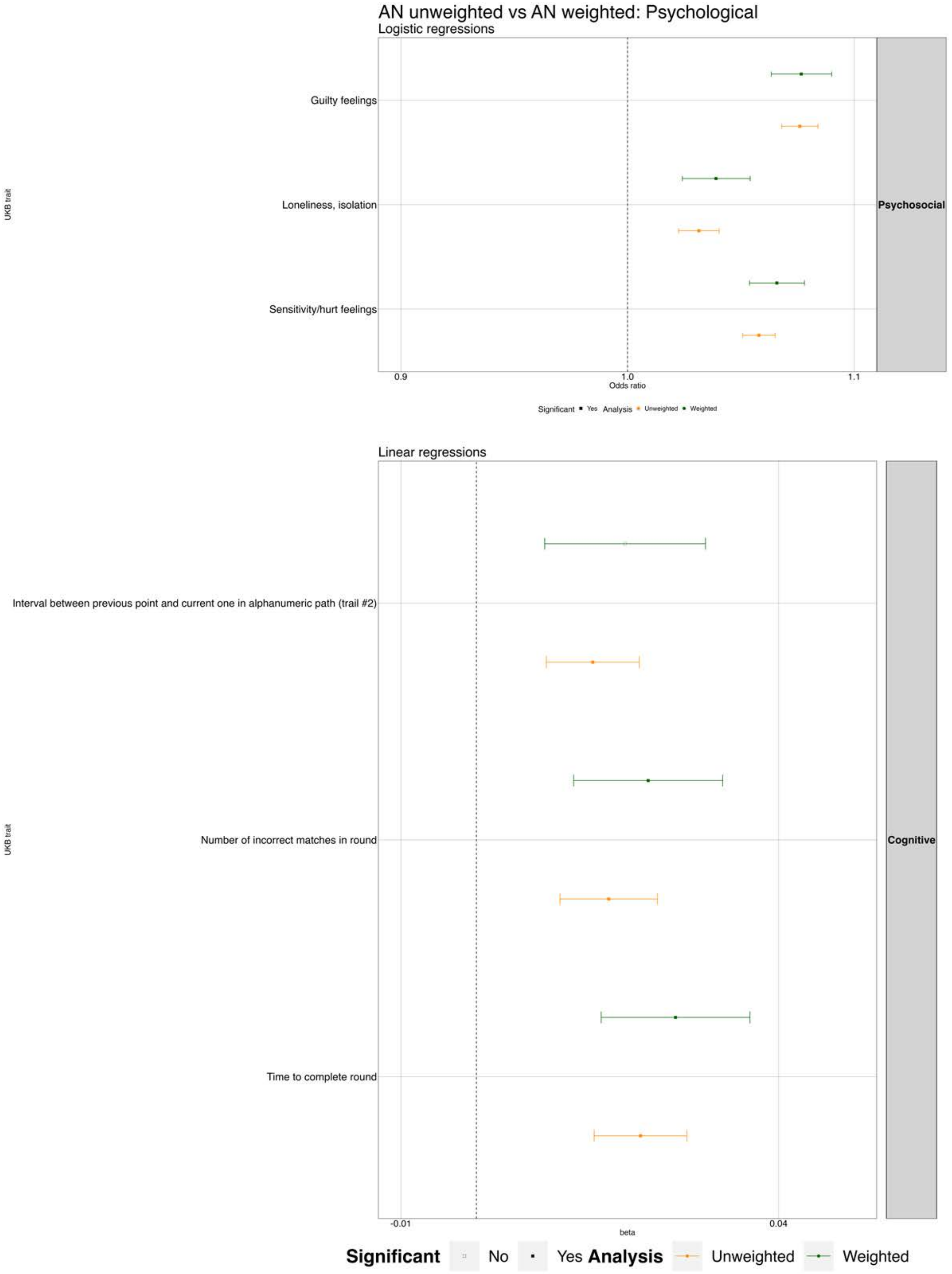


**Figure S43.** Phenome-wide association results between the anorexia nervosa PGS and UK Biobank psychological traits from the main analysis (orange) and after application of sampling weights (green). *Note. Filled squares=significant; Empty squares= non-significant. We have displayed all results that were significant in at least one of the pheWASs.*
